# Development and application of the DePtH framework for categorising the agentic demands of population health interventions

**DOI:** 10.1101/2023.10.18.23297198

**Authors:** Kate Garrott, David Ogilvie, Jenna Panter, Mark Petticrew, Amanda Sowden, Catrin P. Jones, Campbell Foubister, Emma Lawlor, Erika Ikeda, Richard Patterson, Dolly Van Tulleken, Roxanne Armstrong-Moore, Gokulan Vethanayakam, Lorna Bo, Martin White, Jean Adams

**Affiliations:** MRC Epidemiology Unit, University of Cambridge, Cambridge, UK; Department of Public Health, Environments and Society, London School of Hygiene and Tropical Medicine, London, UK; Centre for Reviews and Dissemination, University of York, York, UK; School of Clinical Medicine, University of Cambridge, Cambridge, UK

## Abstract

The ‘agentic demand’ of population health interventions may influence intervention effectiveness and equity, yet the absence of an adequate framework to classify agentic demands limits the fields’ advancement. We systematically developed the DEmands for PopulaTion Health Interventions (DePtH) framework identifying three constructs influencing agentic demand - exposure (initial contact with intervention), mechanism of action (how the intervention enables or discourages behaviour), and engagement (recipient response), combined into twenty classifications. We conducted expert qualitative feedback and reliability testing, revised the framework and applied it in a proof-of-concept review, combining it with data on overall effectiveness and equity of dietary and physical activity interventions. Intervention components were concentrated in a small number of classifications; DePtH classification appeared to be related to intervention equity but not effectiveness. This framework holds potential for future research, policy and practice, facilitating the design, selection, evaluation and synthesis of evidence.

## Introduction

Despite numerous policies attempting to address unhealthy diets and physical inactivity, these practices remain stubbornly common and differentially distributed across the population, contributing to health inequalities.[1] Population health interventions (PHIs) target whole populations or population groups with an aim to reduce disease risk by a small amount across a large population. These are have been described as more appropriate, effective and equitable for primary prevention than interventions targeted at those known to be at risk of disease (so-called ‘high-risk’ interventions).[2] However, PHIs can take a number of different forms and the abundance of evidence available on them can be overwhelming for policymakers to make sense of.[3] Understanding how PHIs work, in what context and for whom[4] and the effect of different interventions on population subgroups is important to drive effective and equitable change.[5] One aspect of PHIs that has been proposed to influence intervention effectiveness and equity is the degree of personal agency individuals have to apply in order to benefit from an intervention.[6] Personal agency includes capacity, resources and freedom to act and achieve an intended outcome.[7]

We use the term ‘agentic demand’ to describe the actions required of individual and organisational actors to enable PHIs to achieve their intended effects. Agentic demand likely exists on a continuum.[6] Interventions with high agentic demands often target individuals’ knowledge and behaviours and rely on individuals’ capacity to act in accordance with intervention aims[8] and to make use of their personal resources, for example time, cognitive or financial resource to benefit.[9] To illustrate, England’s Change4Life campaign provided prompts to recipients to increase walking by getting off the bus one stop earlier than their destination. To realise the health benefit from this, individuals must have and make use of sufficient cognitive resource to understand the prompts, determine how to act on them, remember to act on them, and make use of their temporal resource to implement the strategy over the long term. In contrast, interventions with low agentic demands alter the context within which behaviours are produced and reproduced,[10] focusing on environmental conditions, social institutions and norms that shape individual behaviour.[8] These require little or no personal resources from individuals to realise the intervention aim. For example, when a food manufacturer reformulates packaged snacks to reduce the salt content, individuals will benefit as long as they continue eating the snacks as before.

Over the last 30 years, there have been almost 700 proposed policies for obesity prevention in England. The majority of these placed high agentic demands on individual recipients with only 19% placing low agentic demands on recipients.[1] The ability to meet the agentic demands of interventions is likely to be influenced by a range of social and economic factors. Given personal resources are distributed unequally across the socioeconomic gradient, so too is the ability to respond and benefit from PHIs with high agentic demands likely to be unequally distributed. These interventions may, therefore, contribute to widening health inequalities.[11]

While acknowledged as an important concept,[12] much of the literature exploring agentic demands of interventions applies a simple dichotomy of high vs low agency.[13, 14] An existing framework identified a third intermediate category for interventions which focus on creating supportive environmental conditions but still place an agentic demand on individuals,[11] for example, placing healthy food within a canteen setting creates a conducive environment, yet requires individuals to choose the food. Furthermore, agentic demands are often conflated with other intervention dimensions including high-risk (high agency) vs population (low agency) or intervention mechanism.[9]

While these may be related, they are not synonymous. For example, interventions operating via financial mechanisms are often uniformly categorised as interventions with low agentic demand, yet not all necessarily are. For example, the Healthy Start scheme issues vouchers to low income families in the UK which can be exchanged for fresh fruit, vegetables and milk. To receive vouchers, families must register for the scheme with a health professional’s signature.[15] After using the vouchers to purchase subsidised food they then must have the equipment and knowledge required to prepare the food, placing agentic demands on the individual. In contrast, when visiting a workplace cafeteria with discounted prices on healthy meals, the recipient simply selects the subsidised food, requiring no more agency than any other food selection.[16] These examples illustrate the potential value of a more nuanced and standardised method to classify the agentic demands of PHIs.

To date, the literature has also failed to account for the agentic demands placed on other actors involved in PHIs. For example, a change in school vending machine policy to increase availability of healthier foods may place relatively low agentic demands on users of the machine for them to benefit but requires agreement from the school leadership team and implementation by the vending machine contractor – both activities with high agentic demands. Conditions and strategies to enhance compliance of such actors have been discussed extensively in the literature,[17] yet an understanding ‘what’ these actors are required to do in order to implement interventions, resulting in a ‘layering’ effect of agentic demands in PHIs has not been systematically explored.[18]

The current ad-hoc approach to classifying agentic demands of PHIs is inadequate for capturing their nuance and diversity. A framework to achieve this has potential to improve evidence synthesis by providing a consistent and comprehensive approach to classifying agentic demand. Such a framework may also inform intervention design and prioritisation for use by researchers, public health practitioners and policymakers for understanding how interventions influence inequalities. Here we describe the development of such a framework – the DEmands for PopulaTion Health Interventions (DePtH) framework - and demonstrate its application in a proof-of-concept evidence synthesis to explore associations with intervention effectiveness and equity.

## Results

### The DePtH framework

Here we summarise the final version of the DePtH framework. Supplementary material 2 provides a full description and application guidance.

The framework is applicable to single intervention components, for example, a cycling strategy may include two components: cycling proficiency training and lighting installation on existing cycle lanes. The demands of each component may also vary in different recipient groups, e.g. lighting improvements on existing cycle lanes places different demands on new cycle lane users for their health to benefit compared to existing users. Users of the framework should identify each possible intervention component and recipient combination. The concepts presented herein apply to single component-recipient combinations.

We identified three constructs influencing the agentic demand of PHIs: exposure to the intervention component (two levels), mechanism of action of the intervention component (five levels) and engagement with the mechanism of action (two levels) (Table 1). When combined, these constructs form a matrix of twenty possible classifications (Table 2). We have not sought to order, score or name these categories. Rather we hypothesise that intervention types with similar agentic demands will be grouped within the same framework classification.

**Table 1.**
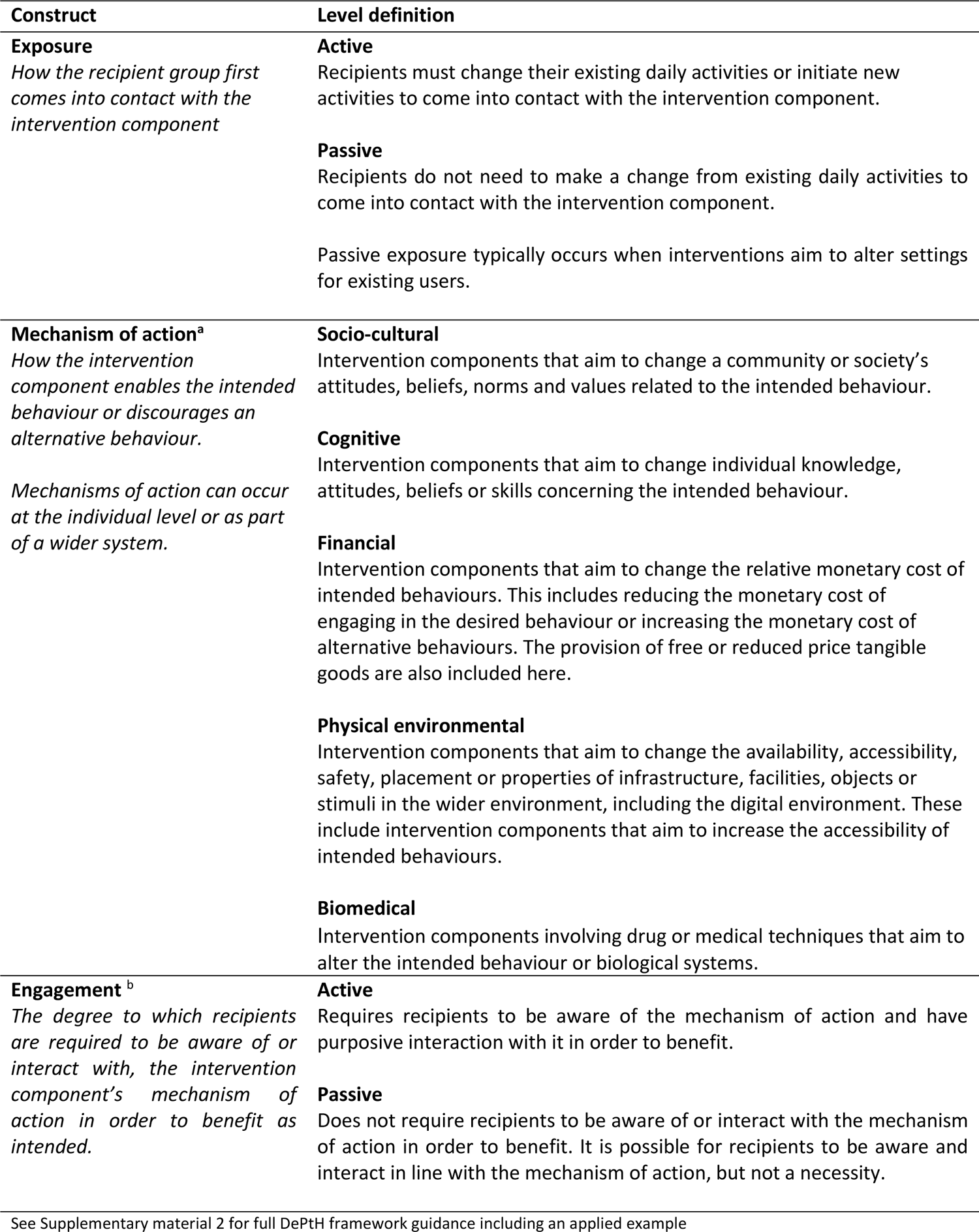
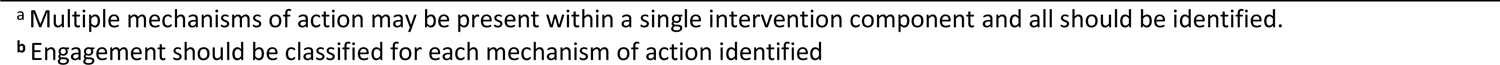
Constructs of the DePtH framework for classifying intervention agentic demand.

| Construct | Level definition |
| --- | --- |
| **Exposure**<br>*How the recipient group first comes into contact with the intervention component* | **Active**<br>Recipients must change their existing daily activities or initiate new activities to come into contact with the intervention component.<br>**Passive**<br>Recipients do not need to make a change from existing daily activities to come into contact with the intervention component.<br>Passive exposure typically occurs when interventions aim to alter settings for existing users. |
| **Mechanism of action<sup>a</sup>**<br>*How the intervention component enables the intended behaviour or discourages an alternative behaviour.*<br>*Mechanisms of action can occur at the individual level or as part of a wider system.* | **Socio-cultural**<br>Intervention components that aim to change a community or society's attitudes, beliefs, norms and values related to the intended behaviour.<br>**Cognitive**<br>Intervention components that aim to change individual knowledge, attitudes, beliefs or skills concerning the intended behaviour.<br>**Financial**<br>Intervention components that aim to change the relative monetary cost of intended behaviours. This includes reducing the monetary cost of engaging in the desired behaviour or increasing the monetary cost of alternative behaviours. The provision of free or reduced price tangible goods are also included here.<br>**Physical environmental**<br>Intervention components that aim to change the availability, accessibility, safety, placement or properties of infrastructure, facilities, objects or stimuli in the wider environment, including the digital environment. These include intervention components that aim to increase the accessibility of intended behaviours.<br>**Biomedical**<br>Intervention components involving drug or medical techniques that aim to alter the intended behaviour or biological systems. |
| **Engagement<sup>b</sup>**<br>*The degree to which recipients are required to be aware of or interact with, the intervention component's mechanism of action in order to benefit as intended.* | **Active**<br>Requires recipients to be aware of the mechanism of action and have purposive interaction with it in order to benefit.<br>**Passive**<br>Does not require recipients to be aware of or interact with the mechanism of action in order to benefit. It is possible for recipients to be aware and interact in line with the mechanism of action, but not a necessity. |

See Supplementary material 2 for full DePtH framework guidance including an applied example
<sup>a</sup> Multiple mechanisms of action may be present within a single intervention component and all should be identified.
<sup>b</sup> Engagement should be classified for each mechanism of action identified

**Table 2.** Matrix classification and intervention examples for DePtH framework constructs.

| Exposure | Engagement | Mechanism of action <sup>a</sup> |  |  |  |  |
| --- | --- | --- | --- | --- | --- | --- |
|  |  | Socio-cultural | Cognitive | Financial | Physical environmental | Biomedical |
| Active | Active | <1% (n=1)<br>*Example: Group nutrition education* | 18% (n=21)<br>*Example: Online self-monitoring of fruit and vegetable intake* | 2% (n=2)<br>*Example: Free bus pass for older adults* | <1% (n=1)<br>*Example: Afterschool physical activity provision* | 0% (n=0) |
|  | Passive | 0% (n=0)<br>*Example: Choosing to move to Healthy New Town with strong cycling culture* | 0% (n=0)<br>*Example: Sign up to SMS service for messages aiming to change individual beliefs* | 0% (n=0)<br>*Example: Provision of food vouchers that restrict purchasing of HFSS food* | <1% (n=1)<br>*Example: Installing online HFSS ad blocker* | 0% (n=0)<br>*Example: Semaglutide drug to suppress appetite and reduce food consumption* |
| Passive | Active | 12% (n=14)<br>*Example: Girl Scout Troop Leader joins in with physical activities* | 37% (n=42)<br>*Example: Educational material within existing church bulletin* | 3% (n=3)<br>*Example: Implementation of SSB tax increases monetary cost* | 11% (n=13)<br>*Example: Provision of free fruit and lunch time <sup>b</sup>* | 0% (n=0) |
|  | Passive | 0% (n=0)<br>*Example: Implementation of SSB tax signals governmental disapproval* | 2% (n=2)<br>*Example: Provision of fruit at lunchtime <sup>b</sup>* | 0% (n=0)<br>*Example: Car parking charges in workplace* | 13% (n=15)<br>*Example: Restrict sugar sweetened beverage portion sizes in schools* | 0% (n=0)<br>*Example: Fluoridation of tap water* |

n = number of intervention component-recipient combinations identified in each category in step 3 (framework application). % = percentage of component-recipient combinations identified in each cell compared to total number identified (n=115). Examples = identified from framework application and if not available, from author knowledge. HFSS = High fat, salt and sugar. SSB = Sugar sweetened beverage
<sup>a</sup> Multiple mechanisms of action possible within a single intervention component. Some examples may be classified under multiple mechanisms, e.g. group nutrition education also operates by cognitive mechanisms.
<sup>b</sup> Intervention authors note two mechanisms of action. (1) Increased availability of fruit (physical environmental); (2) Repeated exposure lead to changes in individual preferences (cognitive)

We also identified four types of actors potentially involved in PHIs (Table 3): (1) Macro-environmental; (2) Micro-environmental; (3) Informal gatekeepers; (4) Secondary recipients. The ability of these actors to execute the actions required for the interventions to achieve their intended effects will be variable, and influenced by structural factors. Further development of this concept was limited by poor reporting and we were not able to proceed further than classifying actors.

**Table 3.** Categories of actors potentially involved in PHIs.

| **Actor category** | **Definition** |
| --- | --- |
| Macro-environmental | Actors at organisational level such as industries, services or supporting infrastructure, which operate at international, national or local level, e.g. food manufacturers, local or national governments. It was rarely possible to specify macro-environmental actors, rather it was clear that action was required at this level to initiate or implement interventions. |
| Micro-environmental | Actors at the level of individual spaces or naturally occurring groups of places where people gather for specific purposes, e.g. actors within schools, individual supermarkets, restaurants, parks. These are usually geographically distinct, relatively small and potentially influenced by individuals. |
| Informal gatekeepers | Actors linked to the intended recipient in a non-professional manner, e.g. parents. These informal gatekeepers must change their behaviour in order for the intervention to achieve the desired effect in the intended recipient. |
| Secondary recipients | Secondary recipients are individuals who may benefit from intervention ‘spill over’ effects, e.g. other members of a household who are affected by food purchasing decisions. |

### Applying the DePtH framework

We applied the DePtH framework within a ‘proof-of-concept’ review. We identified three parent systematic reviews exploring the differential socio-economic effects of dietary and physical activity interventions,[19–21] from which we extracted and screened 87 full text articles on PHIs. We included 33 articles reporting 31 interventions (Supplementary material 3). We were unable to identify intervention components for 5 interventions due to insufficient detail. From the remaining 26 interventions, we identified 163 intervention component-recipient combinations (median = 4.5; range = 1-24 per intervention) and classified the three DePtH framework constructs for 115 of the 163 identified component-recipient combinations. It was not possible to classify the remaining 48 component-recipient combinations due to insufficient detail. Where a framework construct was classified, inter-rater reliability for first assessments ranged from moderate (engagement) to substantial (exposure, mechanism of action) (Supplementary material 2).

We classified the exposure of component-recipient combinations as active (n=26) and passive (n=89); mechanism of action as socio-cultural (n=15), cognitive (n=65), financial (n=5), physical environmental (n=30), biomedical (n=0); and engagement as active (n=97) or passive (n=18). The most common agentic classification was passive exposure, cognitive mechanism and active engagement. Nine classifications and one mechanism of action (biomedical) were not represented at all in the review. Within the 163 intervention component-recipient combinations we identified that macro-environmental (n=135) and micro-environmental actors (n=158) were present in the majority of intervention component-recipient combinations, and that the presence of gatekeepers (n=26) and secondary recipients (n=37) was less common.

Harvest plots[22] show the distribution of intervention component-recipient combinations across the DePtH framework disaggregated by overall effectiveness (Figure 1) and differential effectiveness by socioeconomic position (SEP) (Figure 2). Given the absence of intervention components within some classifications and a small number of components within others, it is only possible to draw tentative conclusions. Figure 1 indicates that the overall effectiveness of interventions on dietary outcomes favoured the intervention group within all but two framework classifications (exposure – active; mechanism of action - physical environmental; engagement – passive; and exposure – passive; mechanism of action – financial; engagement - active). In both these cases, there were few (n≤3) observations. Findings related to the overall effectiveness of interventions on PA outcomes were more mixed. Overall, amongst the most commonly used mechanism (cognitive) for dietary and PA outcomes, there was some indication that interventions were more likely to be effective when exposure was passive rather than active.

**Figure 1.**
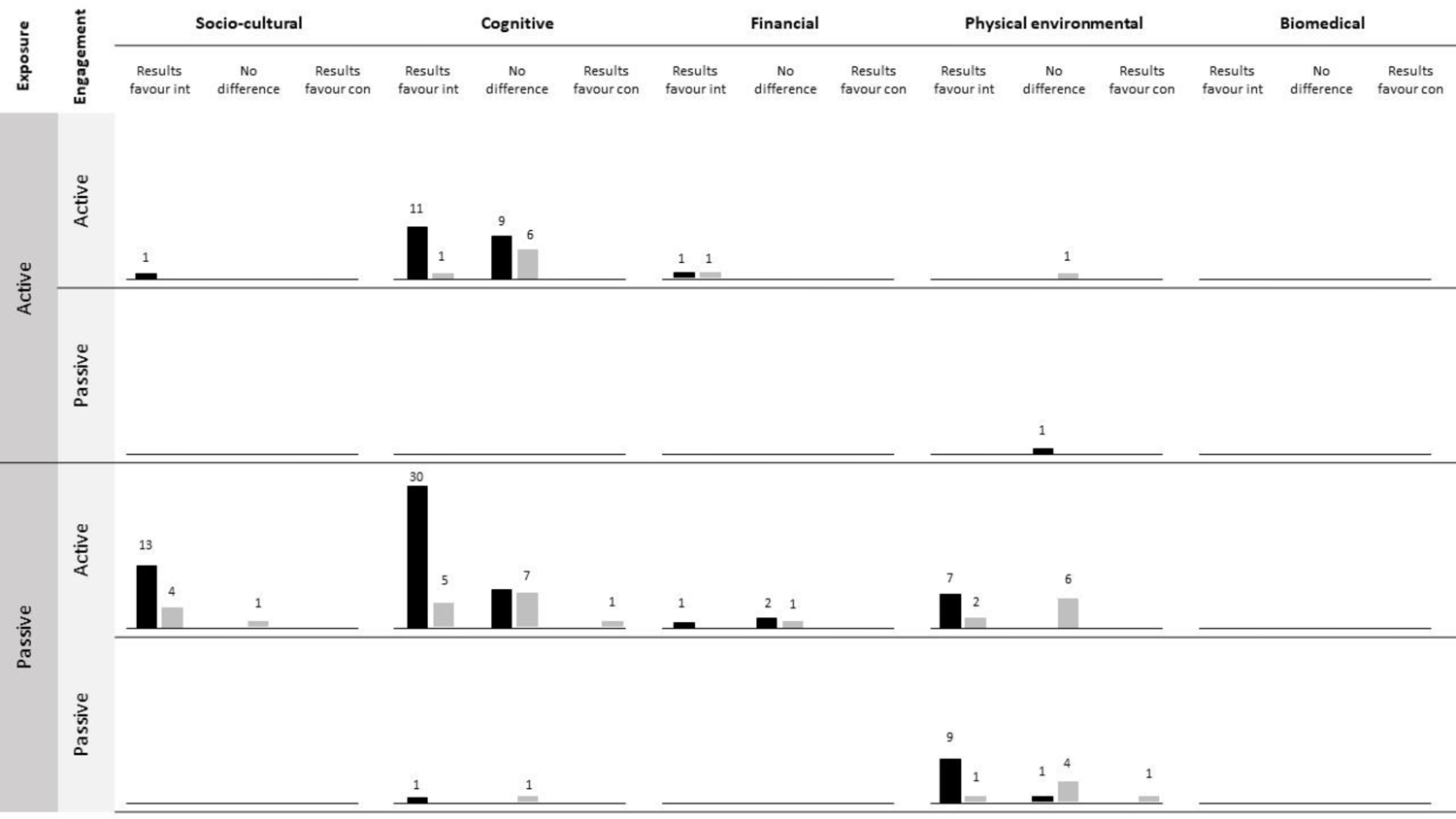
Harvest plot illustrating association between DePtH classification and overall intervention effectiveness. Black bars = dietary outcomes; Grey bars = PA outcomes. Bar height and numbers = number of component-recipient combinations represented in each classification. Int = Intervention group; Con = Control group. DePTh classification is at intervention component-recipient level and effectiveness reported at intervention level.

**Figure 2.**
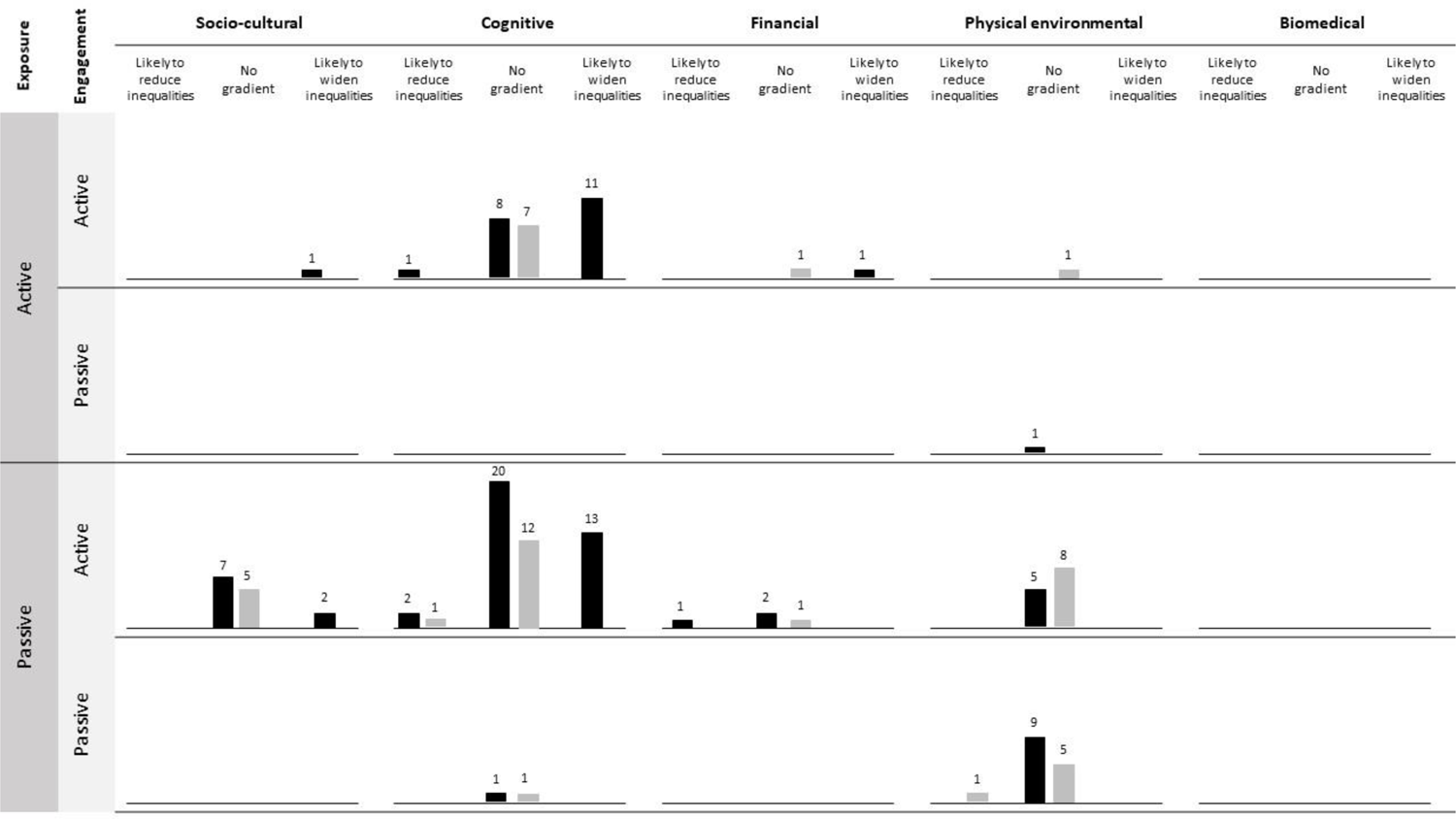
Harvest plot illustrating association between DePtH classification and differential effect by socioeconomic position. Black bars = dietary outcome; Grey bars = PA outcomes. Bar height and numbers = number of component-recipient combinations representing each classification. DePTh classification is at intervention component-recipient level and equity reported at intervention level.

Figure 2 shows that only three intervention components were associated with reductions in socioeconomic inequalities. These included a province-wide physical education policy in Canada,[23] sugar sweetened beverage taxation[24] and a community coalition to promote physical activity.[25] The harvest plots were dominated by data points in the middle column, representing no overall impact on socioeconomic inequalities. There were a considerable number of components targeting cognitive mechanisms that appeared to widen socioeconomic inequalities, although this was less common with passive rather than active exposure. Interventions with socio-cultural and physical environmental mechanisms appeared least likely to have an impact on inequalities.

Despite many interventions containing multiple component-recipient combinations, there was less variation in the number of different DePtH framework categories represented within each intervention (median = 2; range = 1-5 per intervention), indicating that many multi-component interventions include multiple components in the same framework category. Figure 3 provides examples demonstrating a spectrum of clustering of intervention components. Given the sparseness of data in our proof-of-concept review, it is not possible to determine whether clustering of framework classifications is associated with intervention overall or differential effectiveness by SEP.

**Figure 3.**
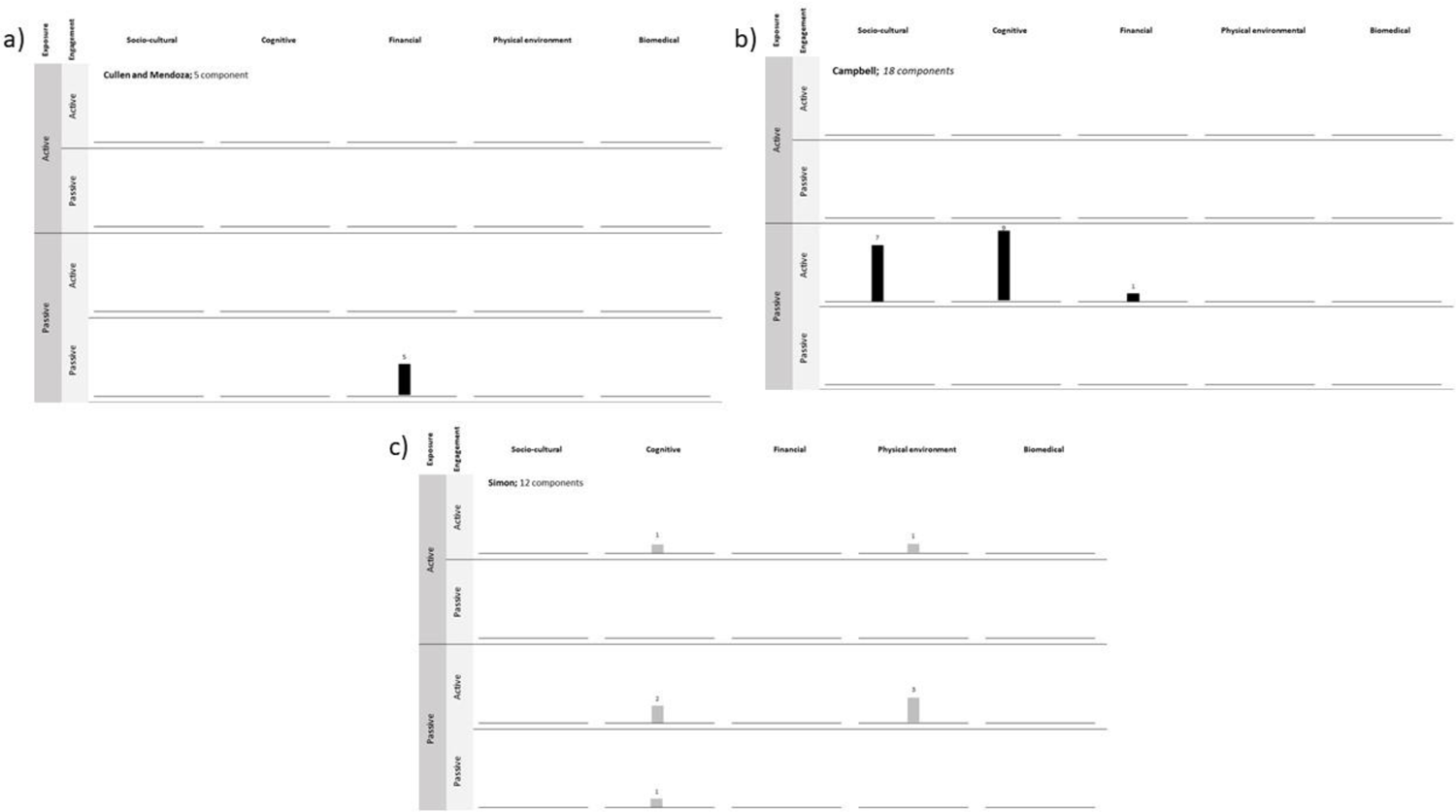
Harvest plots to illustrate the differences in distribution of intervention component-recipient combinations within multi-component interventions. Panel a) intervention with components concentrated within one framework classification. Panel b) intervention with component distributed across multiple mechanisms of action but maintain the same exposure and engagement. Panel c) intervention with components distributed across all framework constructs. DePTh classification is at intervention component-recipient level and effectiveness reported at intervention level.

We provide a detailed account of the application rules we followed for this review in Supplementary material 2. These were guided by our aim to test the framework. Others may wish to apply different rules based on their reasons for using the framework and these should be agreed at the beginning of a project. The application rules we used stem from initial learning from the process presented in box 1 which others may also find useful in developing application-specific rules.

#### Box 1

##### Initial learning for applying the DePtH framework

###### Identify component-recipient combinations a priori

Prior to extracting data on framework constructs, we advise agreeing on the intervention component-recipient combinations. This reduced the number of disagreements at the extraction stage. We identified intervention components to the greatest level of granularity to improve agreement between researchers.

###### Implicit vs explicit information

Not all relevant information is included in intervention reports. For example, some reports may not describe or have explored all mechanisms of action. The degree to which reviewers include only information explicitly included in the report or draw on implicit and wider topic knowledge will be dependent on the review aims and should be agreed a priori.

###### Dealing with insufficient information

Intervention descriptions may not provide sufficient information to classify the framework constructs. In our review, where applicable we chose to classify framework constructs as ‘insufficient information to code’ based on the intervention description. Other approaches may include seeking additional information from a wider range of sources (see below).

###### Consistent application

The framework requires users to apply categorical classifications to constructs that lie on continua. Different users may draw these distinctions in different places. Never-the-less, distinctions should be agreed, reported and applied consistently.

## Discussion

### Statement of principal findings

The DePtH framework is a novel method to standardise the classification of the agentic demands of PHIs based on three constructs: exposure, mechanisms of action and engagement. It also identifies four categories of actors that may be involved in PHIs; (1) Macro-environmental actors; (2) Micro-environmental actors; (3) Informal gatekeepers; and (4) Secondary recipients, yet it was not possible to classify the agentic demand on the actors due to a lack of reporting. The framework was developed through an extensive, iterative process drawing on a systematically assembled pool of interventions and feedback from public health research and policy experts to test usability and reliability.

We have demonstrated that it is possible to apply the DePtH framework within a proof-of-concept review context. Whilst our findings indicate that results favoured the intervention in nearly all DePtH framework classifications, DePtH framework classification may be associated differential effects by SEP. In particular, interventions with passive exposure may then to be more equity-promoting than those with active exposure. The two most frequent framework classification fell within the cognitive mechanism of action with some evidence that this class has the potential to widen health inequalities. Our review did not include any examples in nine framework classifications - particularly those with passive engagement or biomedical mechanisms.

### Strengths and weaknesses of the framework

The DePtH framework makes inroads on unravelling the agentic demands of PHIs on recipients, presenting twenty potential classifications, which we propose as sufficiently discriminatory to group similar intervention types. Existing literature distinguishes the agentic demands of PHIs according to two or three classifications,[6, 11] while these have been important to draw attention to the overarching concept our work demonstrates considerable diversity that may not be addressed by previous classifications. The identification of three constructs within the DePtH framework attempts to explain how agentic demand operates within interventions and contributes to opening the lid on the ‘black box’ of how interventions work to begin dissecting the reasons for intervention successes and failures. Furthermore, the application guidance developed alongside the framework aims to ensure that users can apply the framework consistently.

We acknowledge that the DePtH framework does not adequately address all areas that we set out to explore. Particularly, it was not possible to develop a detailed classification of the agentic demands on the four categories of actors, due to a lack of reporting of the actions required from these actors. Similar limitations were identified when classifying obesity policy in England[1]. We did not consider it appropriate to simply use the same three constructs identified in the DePtH framework for all actors as this could miss important differences involving power, motivations, and population reach of additional actors. As such, the framework cannot address questions relating to intervention implementation and acceptability to actors. However, given that macro-environmental and micro-environmental actors are required in the majority of interventions this feels an important area to explore further. Related literature that was beyond the scope of our methods includes regulatory compliance, exploring the conditions required for such actors to comply with intervention implementation,[17] and may be a starting point to explore this further.

The framework imposes a categorical classifications on constructs that lie on continua, presenting challenges for consistency and reliability. Despite these, we established a process that enabled our team to reach full agreement (Supplementary Material 2). We do not provide this as a definitive instruction manual, rather to be transparent in how we reached the framework classifications in this study. We invite other users to draw inspiration from our approach, but acknowledge that they may require a different application approach to achieve different aims. We encourage users to agree in advance how they will apply the framework for their purpose and report this transparently.

The current DePtH framework identified three constructs, yet some additional intervention features, for example whether the intervention occurs in an environment proximal or distal to the behaviour, may also influence the agentic demands of interventions. At this stage, we deemed that including additional constructs would introduce a level of granularity too great to allow useful evidence synthesis. However, as the framework is used more widely, further important sub-constructs may arise within specific classifications, and we encourage researchers to reflect and report on these.

### Strengths and weaknesses of the methods

A key strength of the methods employed to develop the DePtH framework is the use of standardised methods in each step, yet there pragmatic decisions did have to be made to keep the process manageable. We used systematic methods to search for our initial intervention corpus (Step 1), conducting additional searches to identify all articles and reports to assemble an intervention collection. While our search strategy aimed to identify a breadth of intervention types, our reliance on this corpus of published interventions may have omitted some intervention types leading to missing categories on the framework.

We adopted a proof-of-concept approach to conducting the final review, focusing on a purposive sample of three existing relevant reviews, which limited the number of included studies. Thus, this ‘proof-of-concept’ approach is unlikely to provide a definitive account of all interventions reporting on equity effects of interventions.[26] However, this may also be related to the paucity of studies reporting on the equity effects of PHIs. Additionally, as the source reviews focused on studies reporting differential effects by SEP, they are unlikely to capture on relevant studies of effectiveness. It is therefore likely that our findings are more representative of differential effectiveness than overall effectiveness outcomes. Furthermore, all included studies were based in high-income countries, and it is unclear how the effectiveness and equity effects of interventions differ within other contexts. A strength of the research was involving academic and policy experts in the qualitative assessment and reliability testing of the DePtH framework during step 2. While these audiences represent the main users of the framework, the majority of participants were academics and therefore some user groups may be inadequately represented in the development of the framework. In addition, the core research team comprised only of academics.

The lack of detail provided within intervention reports, particularly those with multiple components was a barrier to establishing the reliability of the framework. It is unclear whether this was due to limited journal space, limited theorisation of interventions, omissions on the part of authors’, or a combination of these factors. As proposed elsewhere, greater use and reporting of intervention theories of change or programme theory may help address this challenge.[4] During our proof of concept review, we applied the framework based on information explicitly provided in reports. While this may have enabled us to reach agreement, the approach led to many cases of ‘insufficient information’. It also limited our classifications to the authors’ interpretation of how an intervention operates and authors may not have identified all possible mechanisms of action. For example, one intervention providing fruit in schools proposed a single mechanism of action of improving availability (physical environmental),[27] yet a similar intervention identified repeated exposure (cognitive) as an additional mechanism of action.[28] To address this issue, other users may choose to draw on their expertise and existing knowledge when applying the DePtH framework, rather than sticking rigidly to information included in intervention reports.

### Unanswered questions and future research

This is unlikely to be the final version of the DePtH framework, and we anticipate that others will suggest modifications and adaptations as they use it. We have developed the DePtH framework for interventions targeting dietary and physical activity interventions, but it is likely to be applicable to other behaviours beyond these, such as tobacco use or alcohol consumption. Our proof-of-concept review identified an absence of interventions within some DePtH classifications, and it was considered premature to remove these before it has been used more widely. Further work could explore whether such interventions are possible and, if so, why they are so infrequently reported. Possible reasons include that: they are uncommonly used, less likely to be reported when used, or used for behaviours beyond diet and physical activity. Either way, these areas may represent particularly fruitful opportunities for innovation. Relying on existing intervention reports to develop a deep understanding of the agentic demand on other intervention actors was not sufficient to address our original aims. In order to address this, further work is required to examine and report intervention implementation in detail. Furthermore, our review focused on exploring the relationship between placement on the DePtH framework and overall and differential effectiveness. Future research could explore associations with other outcomes such as intervention acceptability, safety, empowerment and equity based on measures beyond SEP, such as those included in the PROGRSS Plus criteria.[29]

We have demonstrated it is feasible to use the DePtH framework within a ‘proof-of-concept’ review, suggesting that intervention agentic demand is associated with intervention equity. However, the methods did not enable us to definitively answer this question. Extending our approach to a wider corpus of literature is a next step that is likely to advance the evidence base and our understanding of how different intervention components influence effectiveness and equity. Such a review may need to make use of strategies such as contacting study authors to obtain additional equity data or utilising existing expert topic knowledge to interpret intervention details. Such a review could also explore whether the distribution of components across framework classifications in multi-component interventions is associated with intervention effectiveness and equity. For example, some intervention components that place a lower agentic demand on individuals, such as those with passive exposure and engagement may mitigate the higher demands from other components such as those that require active exposure or engagement. Distribution of intervention components across the framework may diversify the potential for effect of interventions across individuals and contexts. In contrast, concentration of multiple components in the same framework classification may reinforce particular effects.

While we have utilised the DePtH framework within a review to retrospectively assess intervention agentic demand, the framework could also be utilised prospectively by researchers, public health practitioners and policy makers with leverage to design, refine or evaluate interventions. It may also be possible to use the framework as a blueprint to explore less commonly implemented and evaluated interventions.

## Conclusion

The DePtH framework provides a method of classifying intervention agentic demand that advances current approaches by addressing the complexity of PHIs and provides guidance for consistent classification. It provides a description of a concept proposed to influence differential intervention effects by SEP and thus has the potential to play a role in understanding how interventions work for different population groups. We encourage users to build on the current framework, exploring it’s transferability to other behaviours and its association with other relevant outcomes. Future work to understand how the DePtH framework can inform intervention design is critical to ensure that implemented interventions account for agentic demand and do not inadvertently reinforce existing socioeconomic inequalities in health.

## Methods

Below we describe the three steps taken to develop, test and apply the DePtH framework. Firstly, we sought to develop a draft framework by systematically identifying PHIs and synthesising coding of the actors and their actions. Secondly, we tested the framework by seeking expert qualitative feedback and reliability testing and used the results to refine the framework. Thirdly, we assessed the applicability of the framework within an evidence synthesis. Whilst this is presented as a sequence of steps, in reality it was an iterative process (Figure 4). A detailed account of the methods is included in Supplementary material 1 and protocols were pre-registered on Open Science Framework (https://osf.io/nz23j/).

**Figure 4.**
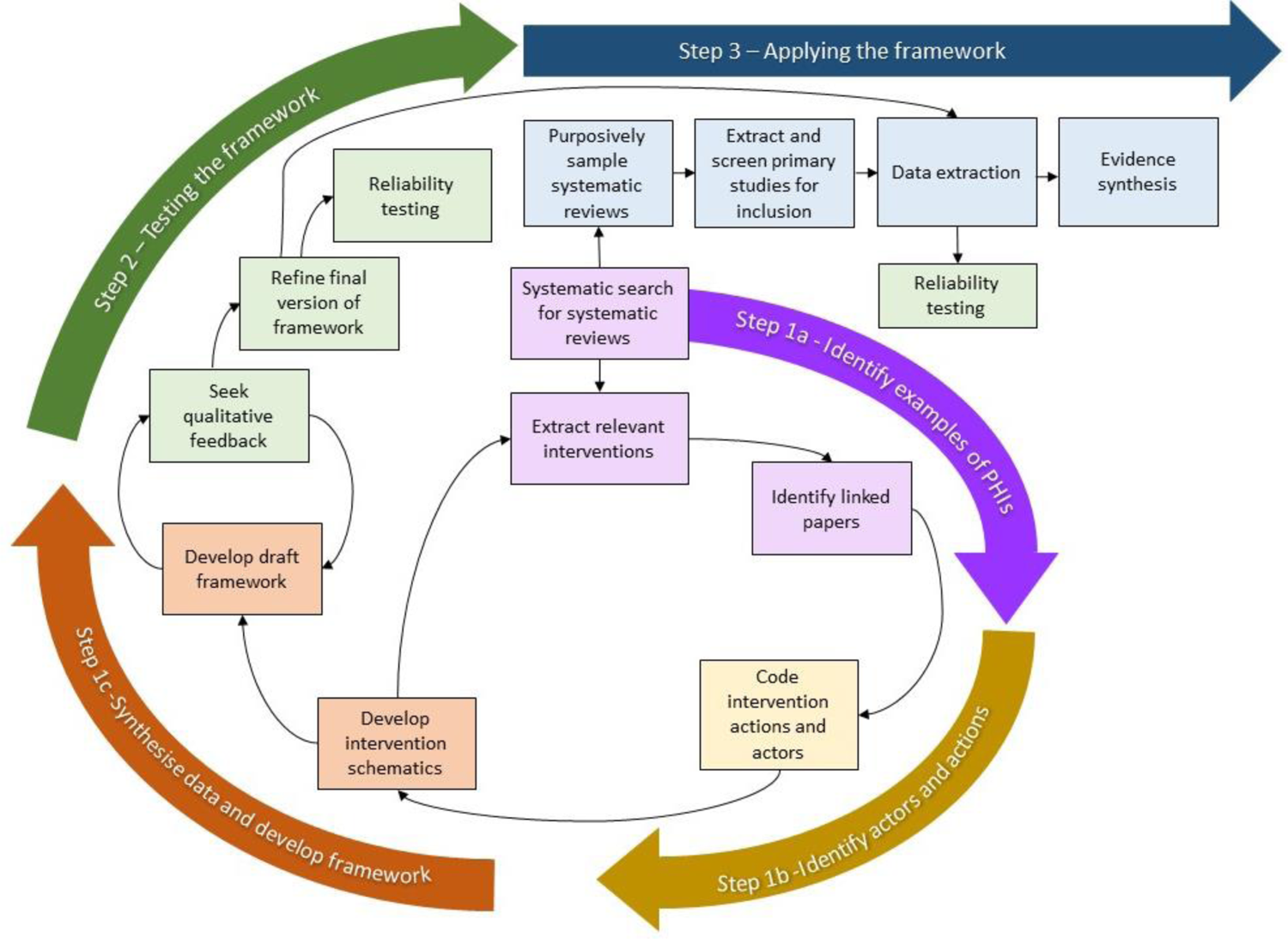
Iterative methods for developing and applying the DePtH framework

### Step 1 – Developing the DePtH framework

#### a ) Identify examples of PHIs

In step 1, we aimed to identify a breadth of PHIs aiming to promote dietary and physical activity (PA) outcomes that could be used to identify a range of actors and their actions from which to develop the DePtH framework. In step 1a we identified systematic reviews likely to include PHIs aiming to promote diet and PA. We defined PHIs as interventions available to whole populations, or population groups defined by non-health indicators in the PROGRESS-PLUS criteria[29]. We excluded high-risk interventions available only to individuals with the presence or absence of a health indicator. Reviews were included if they aimed to impact on dietary or PA outcomes.

We conducted a two stage systematic search using purposive and random sampling of articles where appropriate to maintain a manageable number and breadth of reviews. Firstly, we searched nine databases (MEDLINE, EMBASE, Social Citation Index, CINAHL, Transport Research International Database, Social Science Citation Index, PsychInfo, Applied Social Science Index and Abstracts and International Bibliography for the Social Sciences) to identify systematic reviews that would be likely to include dietary or PA PHIs. We removed duplicates and two reviewers (KG and CPJ/CF/EL/EI/RP/DT/RAM) independently screened the titles and abstracts of 25% randomly selected records (n=8077) followed by the full texts of those included following this screen (n=749). After examining full texts, 408 reviews met the inclusion criteria, from which we purposively selected 9 likely to provide a breadth of intervention types. We identified potentially relevant interventions from their tables of study characteristics. We retrieved full text articles describing these potentially relevant interventions (n=375) and two reviewers independently screened them to identify PHIs aiming to impact on diet and PA outcomes (n=74). To retrieve as much information as possible for each included intervention, we searched Google Scholar, PubMed and funders’ websites to identify linked articles, for example, protocol papers, funder reports or process evaluations. The final collection included 74 interventions, described in 314 articles.

#### b ) Identifying actors and their actions

In step 1b, we used the data in the 74 interventions identified in step 1a to identify all intervention actors and their actions. *Actors* referred to people required to conduct an action for the intervention to have its intended effect on diet or PA. *Actions* were defined as what the actor was required to do in order for the intervention to have its intended effect. We coded all actors and actions explicitly described in the 314 articles from the stage of intervention implementation. Many interventions contained multiple components. We coded actors and actions separately for single intervention components defined as a single pathway or chain of action within an intervention with an intended outcome of dietary or PA change. These are singular aspects of interventions that recipients might ‘see’, for example cycle lanes or point of decision prompts.

#### c ) Synthesising data and developing the DePtH framework

In step 1c, we combined actor and action codes for similar intervention types to develop schematic flow chart diagrams explaining ‘who had to do what’ for single intervention components to be implemented and have their intended effects. We developed the diagrams iteratively, merging similar interventions to refine each diagram. This process was repeated to produce a final set of diagrams (n=8), used to identify concepts, which we organised into a draft conceptual framework (Supplementary material 1, page 69). Data clinics were conducted with the core research team to test and refine the organisation of the draft framework.

### Step 2 – Testing the DePtH framework

In step 2 we iteratively developed the draft framework (and associated user instructions) based on qualitative feedback from relevant experts and reliability testing. The University of Cambridge School of Humanities and Social Sciences Research Ethics Committee granted ethical approval [21.284] for step 2.

#### a ) Seeking expert qualitative feedback

In step 2a we conducted four online workshops with academic and policy experts (n=20) with experience of developing, implementing, evaluating or synthesising evidence of PHIs to promote diet and PA. The disciplinary backgrounds of participants were public health (80%), health economics (5%), health psychology (10%) and health services research (5%). We circulated a copy of the draft DePtH framework ahead of the workshops. During the workshops, participants applied the draft DePtH framework to six intervention examples and used this experience to contribute to the structured discussion. The discussion aimed to explore the content validity and practical utility of the DePtH framework and was facilitated by a member of the research team. We audio recorded the workshops and two researchers wrote detailed field notes. Following workshop feedback, we extensively refined the DePtH framework in three key areas; (1) terminology and categorisation; (2) framework structure; (3) user instructions. We sought verbal feedback on the revised DePtH framework from a purposively selected subsample of workshop participants and produced a final version.

#### b ) Reliability assessment

In step 2b, we conducted an online survey to assess the inter-rater reliability of the final version of the DePtH framework. We recruited a new sample of academic experts (n=22) with similar experience as in step 2a to code 53 intervention examples randomly selected from those identified in step 1a. We used the KappaSize R package[30] to estimate an approximate sample size for the number of interventions to assess, as previously.[31] We estimated the sample size based on the following parameters: alpha value of 0.05; power of 0.8, probability of 0.7, a null hypothesis of a kappa of 0.4 and an expected kappa of 0.7. This suggested that two independent reviewers applying the final DePtH framework to 53 interventions would be required to test if κ>0.4. The online survey involved reading a description of the final DePtH framework and user instructions (Supplementary material 2), and applying the framework to each intervention example. We encouraged participants to provide free text responses to justify or explain each decision. Each intervention example was independently coded by two participants. We asked each participant to code up to five intervention examples based on the time they had available.

We calculated Cohen’s Kappa to assess inter-rater reliability of each categorical item in the survey. Open-ended text answers were coded and compared by one researcher. Kappa values were interpreted as follows: ≤0 no agreement; 0.01-0.2 none to slight; 0.21-0.40 fair; 0.41-0.60 moderate; 0.61-0.8 substantial and 0.81-1.00 as almost perfect agreement. Of 3 constructs and 4 actors included in the DePtH framework, Cohen’s Kappa assessment classed two each as fair, moderate and no agreement and one as none-slight agreement. Because of these low levels of agreement, we explored inter-rater reliability further in step 3.

### Step 3 – Applying the DePtH framework

In step 3 we demonstrated the application of the DePtH framework in a review. We explored the association between intervention agentic demand, as categorised by the DePtH framework, and reported overall and differential effectiveness by SEP. To identify studies that reported effects by SEP, we first searched systematic reviews identified in step 1 (n=32,306) for those that included a term related to equity in their title based on a validated filter for ethnic and socioeconomic inequalities (n=24).[32] From amongst these, we purposefully selected three systematic reviews that provided a breadth of intervention types across dietary and PA behaviours and presented a differential effect by SEP.[19–21] From these three systematic reviews, we extracted included studies (n=87), removed duplicates (n=9) and screened full text articles (n=78) according to the inclusion criteria. We included PHIs aiming to promote diet and PA according to the inclusion criteria in step 1, and which reported both a measure of overall effectiveness and measures of effectiveness in subgroups differentiated by at least one measure of SEP. Measures of SEP included, income, occupation, education at household, parental or area level. We excluded simulation and modelling studies. One reviewer screened all full text articles and a second reviewer independently screened 50% of the articles for inclusion.

We then extracted data from primary included studies (n=31) on study characteristics, outcome measures and coded interventions according to the DePtH framework. We followed application rules developed specifically for this step (Supplementary material 2). Two reviewers (KG and GV or LB) independently extracted data according to the final DePtH framework and a third reviewer (JA) resolved disagreements. We calculated inter-rater reliability of this process as described in Step 2b. Overall effectiveness data was classified into one of three categories; (1) Results favour intervention–any changes in dietary or PA outcomes associated with the intervention are in a direction that supports public health; (2) No difference – no change in relevant outcomes associated with the intervention; (3) Results favour control – any changes in relevant outcomes associated with the intervention are in a direction that doesn’t support public health. If a primary outcome was stated, we categorised intervention effects for this. If a primary outcome was not stated, we classified intervention effectiveness for each relevant outcome and selected the most common effectiveness category across all outcomes.

We extracted data on equity effects across levels of socioeconomic position. We categorised equity effects into one of three categories; (1) Likely to reduce inequalities – the intervention preferentially improves outcomes in people of lower socioeconomic position; (2) No preferential impact by socioeconomic position, including those where there was an overall effect but no differential effect by socio-economic subgroups; (3) Likely to widen inequalities – the intervention preferentially improved outcomes in people of higher socioeconomic position.

We generated harvest plots to aid evidence synthesis by data visualisation,[22] plotting each component-recipient combinations according to its DePtH classification and intervention-level categorisation for effectiveness and equity.

## Data Availability

All data produced in the present work are contained in the manuscript

## Author contributions

KG and JA were involved in the conception, design, data collection, analysis and interpretation of the study. KG wrote the abstract with input from JA. MP, DO, JP, MW, AS were involved in the conception, design and interpretation of the study. CPJ, RP, RAM, DT, EL, CF, EI were involved in study screening during systematic identification of interventions. GV and LB were involved in study screening and data extraction in the systematic review. All authors have seen and approved the final version of the abstract for publication.

