## Supplementary material 1 for "Development and application of the DePtH framework for categorising the agentic demands of population health interventions"

### Supplementary material 1 – detailed account of methods and results for applying and testing the DePtH framework

#### Step 1: Developing the DePtH framework

##### Step 1a. Identify a selection of population health interventions

*Systematic scoping search to identify population health interventions for diet and physical activity*


###### Summary

*Aim:* To identify a diverse collection of population health interventions (PHIs) with the potential to promote healthier dietary intake or physical activity (PA).

*Methods:* We conducted a two stage systematic scoping review to identify relevant articles, firstly searching nine databases to identify systematic reviews that include PHIs to promote healthier diets or PA. Duplicates were removed and two screeners reviewed the titles and abstracts of records followed by full text articles. From the included articles, we identified potentially relevant interventions from the information included in the systematic review, such as the table of study characteristics. Full texts of potentially relevant interventions were retrieved and screened. Throughout this process, we used purposive and random sampling of articles where appropriate to ensure the number of articles was manageable while maintaining the breadth of the intervention collection. For each included article, we identified linked articles by conducting further searches using google scholar, PubMed and the funders website using first and last author and if available, intervention name.

*Results:* We identified 32,306 records following duplicate removal. We randomly selected and screened 25% of these articles (n=8077), resulting in 408 eligible systematic reviews. We purposively selected 9 systematic reviews representing a broad range of interventions and from these 74 included interventions, described in 314 articles.


###### Methods

**Identify systematic reviews**

In the first step of this process, we identified systematic reviews that include at least one PHI to promote healthier diets and PA. The eligibility criteria for systematic reviews is included in Supplementary Appendix 1.1 and outlined below.

*Study design*

We included systematic reviews published in a peer reviewed journal. Any reviews not published in a peer-reviewed journal, for example, in the grey literature or conference abstracts were excluded. For the purpose of this step, we defined a systematic review as a review that reported a specified or recognised method and described a search strategy and inclusion criteria. The location of the full search strategy must had to be accessible to the reader, for example, appendices, supplementary or additional material or in a registered protocol openly available. Where search strategies were available upon request to the corresponding author we did not consider this accessible to the reader. Systematic review protocols and reviews of reviews were excluded, but were used for pearl growing. For relevant protocols we searched for the subsequent paper. For relevant reviews of reviews we hand searched the reference lists for systematic reviews that met our inclusion criteria.

*Population*
Systematic reviews were included if they include a PHI. For the purpose of this review, we define a PHI as an intervention available to whole populations or population groups defined by non-health indicators. We used the PROGRESS-PLUS[1] criteria to define non-health criteria which may have been used to identify population groups. Thus, we included interventions targeting populations defined by the following criteria; place of residence; race, ethnicity, culture or language; occupation; gender or sex; religion; education; socioeconomic status; social capital; features of relationships (eg, excluded from school, inactive parents); time dependent relationships (where a person may be at a temporary disadvantage); or personal characteristics associated with discrimination. We based the characteristics associated with discrimination on the UK protected characteristics[2] of age, gender reassignment, marriage and civil partnership, pregnancy and maternity, race, religion or belief, sex or sexual orientation. The UK protected characteristic of disability, defined as 'a physical or mental impairment which has a substantial and long-term adverse effect on that person's ability to carry out normal day-to-day activities'[2], was excluded because it includes populations with long-term conditions, which are therefore groups defined on the basis of health indicators.

We excluded systematic reviews of interventions for high-risk populations, defined as those only available to individuals with the presence or absence of a health indicator, identified by screening or self-assessment. For the purpose of this review, this included screening on weight, disease risk, confirmed disease diagnosis or health behaviours, eg, dietary patterns, smoking, PA levels. For example, an intervention excluding individuals who meet the PA guidelines was not included.

*Interventions*
We included systematic reviews that included at least one PHI. As our intention was to identify a diverse collection of relevant intervention examples (rather than all possible examples of all possible interventions), we included systematic reviews that focused on interventions to achieve particular dietary or PA outcomes. We also included systematic reviews of specific types of intervention where these were PHIs, eg, menu labelling or cycle lanes.

*Outcomes*
Systematic reviews that included interventions with the potential to improve dietary and PA behavioural outcomes, where these outcomes have been measured were included. For dietary outcomes, systematic reviews that measured food selection, purchasing or consumption or combinations of these, were included. We included reviews that captured these outcomes across the total diet as well as during individual selection, eating and purchasing occasions. Reviews of interventions promoting breastfeeding were included. For PA, systematic reviews measuring total PA domain specific PA, activity specific PA or any combination of these were included. Systematic reviews that only reported physical fitness outcomes, eg, $VO_2$ max or peak oxygen uptake were excluded. Reviews with sedentary behaviour outcomes were excluded because this is a behaviour independent from PA. As PHIs to improve diet and PA are often framed as obesity prevention interventions and measured by weight related outcomes, we also included systematic reviews with weight related outcomes, including but not limited to body mass index, weight, waist circumference, body fat or waist-to-hip ratio.

We considered including systematic reviews of interventions to prevent development of diseases and their risk factors known to be associated with diet or PA, such as prevention of cardiovascular disease or diabetes. The scoping searches identified a large amount of clinical evidence relating to disease prevention interventions and we decided to remove these from the review. Therefore, systematic reviews measuring the effect of interventions on health outcomes, disease incidence or mortality rates were excluded.

*Language*
We included systematic reviews published in any language, subject to locally available translation capacity.

*Date*
For pragmatic reasons, we included systematic reviews published since 2010.

***Information sources and search strategy***

*Electronic searches*
To identify relevant systematic reviews, we searched the following electronic databases of peer-reviewed literature covering biomedicine (MEDLINE, EMBASE, Science Citation Index) nursing and allied health professionals (CINAHL), transport (Transport Research International Database) and the social sciences (Social Science Citation Index, PsycInfo, ASSIA (Applied Social Science Index and Abstracts) and IBSS (International Bibliography for the Social Sciences)). We did not conduct grey literature searches as it is unlikely that many systematic reviews are available only in the grey literature.

We developed detailed search strategies with the assistance of a specialist medical librarian. The review team identified additional terms omitted from the strategy. The search strategy was based on the concepts of (1) dietary and PA outcomes AND (2) systematic reviews AND (3) health interventions. We based the search terms for diet or PA on current UK guidance[3, 4]. For diet, these are to increase consumption of fruit and vegetables, fibre and wholegrains and oily fish; choose low fat dairy over high fat and lean cuts of meat; reduce consumption of red and processed meat, foods high in fat, salt and sugar (such as cakes, biscuits, confectionary and salty snacks) and total energy and; promote breastfeeding. For PA, these are to increase PA of moderate, vigorous, very vigorous intensity, or any intensity combination. We also included search terms for walking, cycling and running, the three most common types of PA in the UK according to the 2019 Active Lives Survey.[5] The SIGN filter for systematic reviews[6] was applied to search for systematic reviews in the medical databases and adapted as necessary for the social science and transport databases. The searches were limited to articles published since 2010. The MEDLINE search strategy is included in Supplementary Appendix 1.2 and was modified for each database to account for database specific terms.

***Study selection***

We conducted title and abstract screening followed by full text screening to identify systematic reviews that could include at least one population intervention with potential to promote healthier dietary and PA behaviours. Results from the electronic database searches were imported into Covidence software and duplicates removed. We conducted an initial pilot between all reviewers to refine the inclusion criteria. All reviewers screened title and abstracts of the same 400 articles against the eligibility criteria and met to discuss discrepancies and to refine the inclusion/exclusion criteria. At this stage, we refined the inclusion criteria to provide clarity of relevant definitions of systematic review and eligible populations. Following this, two reviewers screened the titles and abstracts to identify and exclude publications that definitely did not meet the inclusion criteria.

We retrieved the full text articles of remaining records, which were screened independently by two researchers and reasons for exclusion noted. Disagreements were resolved by discussion between reviewers, or with the wider review team if necessary. We planned to identify additional relevant systematic reviews by searching reference lists of included studies and conduct forward citation searches. However due to the large number of identified reviews, this step was not deemed necessary.

***Identify interventions***
In order to identify a breadth of interventions, we purposively sampled the included systematic reviews to select a collection that were likely to provide a breadth of intervention types. This was defined as reviews that were focused on outcomes, eg, salt intake, active travel as opposed to intervention specific, eg, digital interventions, point-of-decision prompts. Interventions within these reviews were eligible if they met the criteria specified in table 1.1. During this stage, we used the reported information in the systematic review, eg, table of study characteristics to make decisions. We extracted the publication details of potentially eligible articles and retrieved the full text articles, which were also screened. Two members of the research team (KE and JA) piloted this process on 10% of the selected systematic reviews, demonstrating good agreement. Therefore, screening was completed by one researcher (KE).

**Table 1.1 -** *Eligibility criteria for selecting intervention examples*

| | **Inclusion** | **Exclusion** |
|---|---|---|
| **Population** | Intervention is accessible to whole populations or population group defined by a non-health indicator;<br>• Place of residence<br>• Race, ethnicity, culture or language<br>• Occupation<br>• Gender or sex<br>• Religion<br>• Education<br>• Socioeconomic status<br>• Social capital<br>• Features of relationships (eg, excluded from school)<br>• Time dependent relationships (person at a temporary disadvantage)<br>• Personal characteristics associated with discrimination (age, gender reassignment, marriage and civil partnership, pregnancy or maternity, sexual orientation) | Interventions only available to individuals with the presence or absence of a health indicator identified through screening or self-assessment, eg,<br>• weight status<br>• confirmed disease risk<br>• confirmed disease, eg, cancer diagnosis, stroke<br>• health behaviour, eg, PA levels or dietary pattern<br>Interventions in disabled populations |
| **Intervention** | Interventions with the potential to improve dietary quality or increase the frequency or intensity of PA<br><br>Interventions targeting individual behaviours or a combination of behaviours. | |
| **Outcomes** | Include a measure of dietary quality<br>or<br>Include a measure of frequency, duration, intensity or total volume of PA.<br>or | Measures of health that are not specific to diet, PA or obesity, eg, quality of life. |

| | | |
|---|---|---|
| | Include a measure of weight or anthropometric measure related to weight, eg, waist circumference or body fat. | Measures of physical fitness only or participants attitudes, perceptions or motivations<br><br>Sedentary behaviour outcome only |
| **Study design** | Experimental studies whose design allows a comparison measure, eg, comparison to a control group or pre-post measures.<br><br>Published in peer reviewed journal. | |
| **Year of publication** | Any year | N/A |
| **Country** | Any country | N/A |
| **Language** | Any language, subject to local translation capacity | N/A |

***Identify linked papers***

For each selected intervention, we conducted further searches to identify linked sources of information. We searched in Google Scholar, PubMed and funder websites by the name of the first and last authors and, if available, intervention name. We used four criteria to confirm whether an information source was linked to the selected intervention. To be linked, additional sources of information must have included; (1) a description of the intervention that closely matched the index paper, ie, the one identified in our original search; or (2) a description of the intervention setting; or (3) a description of the causal pathway; or (4) similar author group. This method is used in systematic searching to identify linked information sources.[7] There was no limit on the type of publication at this stage and we included qualitative or quantitative studies, policy documents, grey literature, conference abstracts or reports.

#### Results

***Identifying systematic reviews***

A PRISMA diagram detailing the search process is included in Figure 1.1. 32,306 records were identified from 9 databases. To maintain a manageable number of records and within the scope of the review, we randomly selected 25% of records to screen (n=8077). 749 full text articles were screened and 408 eligible systematic reviews were identified.

Of the 408 systematic reviews, we randomly selected 200 and extracted data on target population; socioecological model description of included interventions; behavioural focus, number of included studies and the review focus (outcome, e.g. salt reduction vs intervention, e.g. digital interventions) in order to aid purposive selection of a collection of systematic reviews that could include a broad range of interventions. Of the 60 broad reviews, we purposively selected 9 systematic reviews that included a broad range of interventions (systematic review references provided in Supplementary Appendix 1.3).

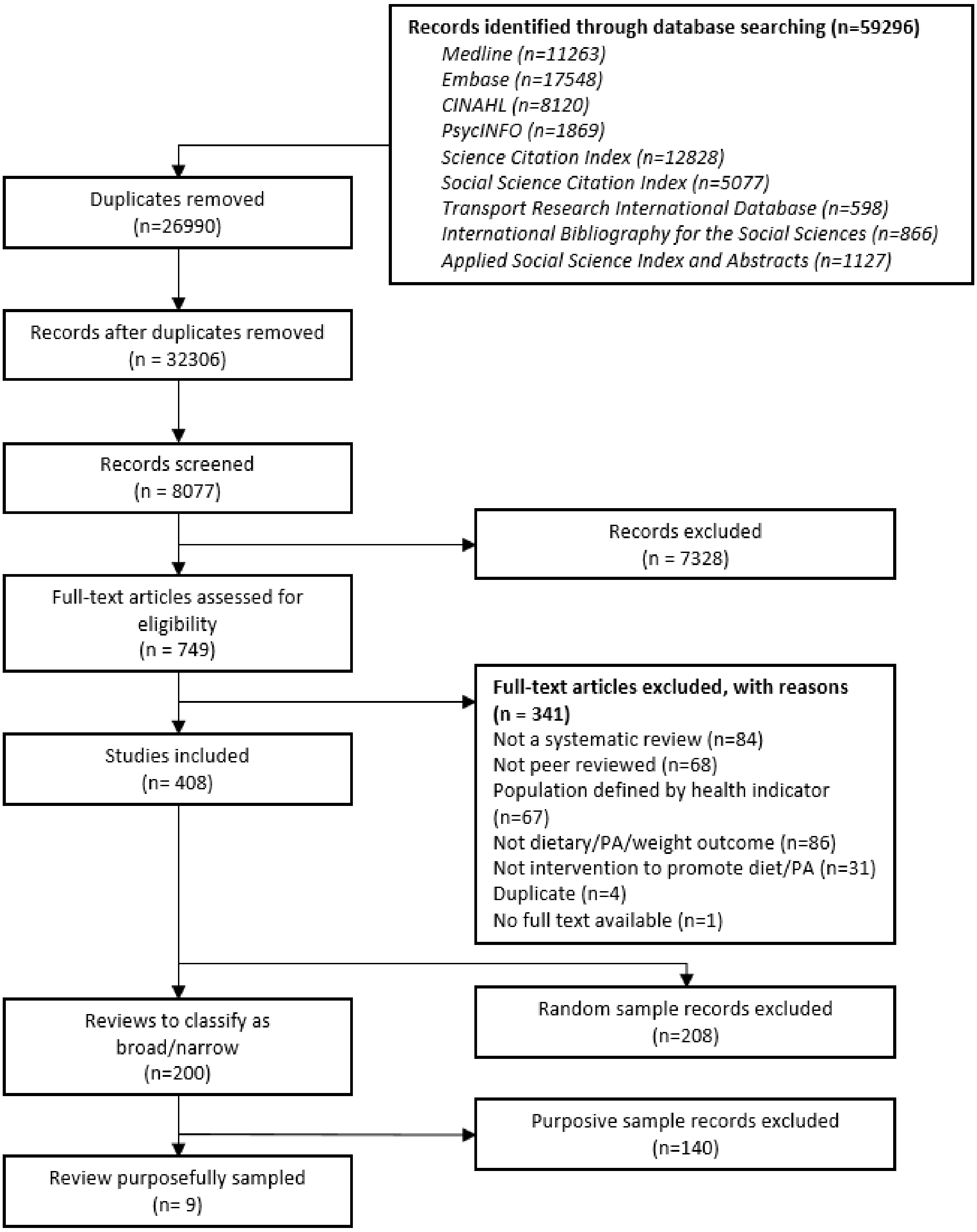


**Figure 1.1 –** *Selection of systematic reviews PRISMA diagram*

***Identify interventions***

A PRISMA diagram for identifying interventions is provided in figure 1.2. The 9 systematic reviews included 409 articles. We reviewed each study according to the information included in the source review's table of characteristics and excluded 22 interventions and identified 12 duplicates. We screened 375 full text articles, of which 214 were excluded and 161 articles describing 141 studies met the inclusion criteria. The 141 studies yielded a large number of school based interventions, which were very similar and did not satisfy our objective to collect a broad range of intervention types. Therefore, we randomly selected 10% of articles on school-based interventions and excluded the remainder. Of the 141 eligible studies, we included 74 studies for the remainder of step 1. We conducted this stage of screening iteratively.

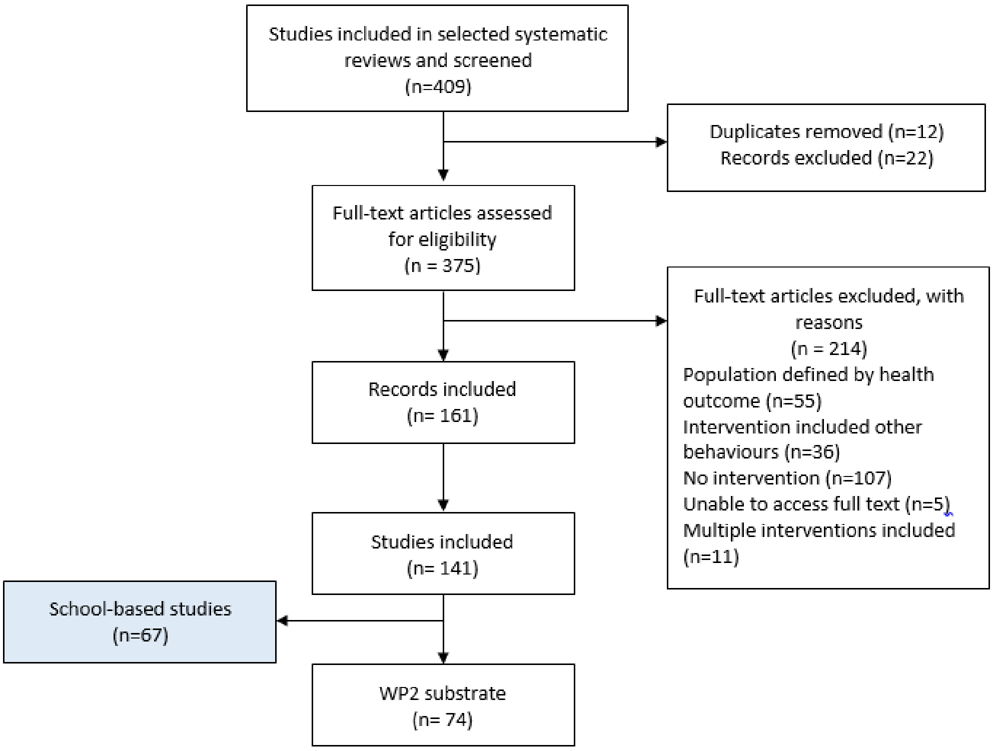


**Figure 1.2** – *Intervention selection PRISMA diagram*

##### Step 3: Identify linked papers

From the 74 included studies, we identified a total of 331 linked papers, of which we retrieved full text articles for 314. All articles are included in Supplementary Appendix 1.4. The included studies and the linked papers formed an intervention collection, which was used in step 1b.

#### Step 1b. Identify actors and actions

*Code actions involved in PHIs and the actions required of them*

##### Summary

*Aim:* To code all included interventions to identify the actors involved in PHIs and the action required of them for the intervention to have its intended effect.

*Methods:* We coded intervention descriptions according to the actors involved and their actions. Actors refer to 'who' has to conduct an action for the intervention to have an effect on diet or PA and may be an individual, group or organisation. Actions were defined as what the actor was required to do in order for the intervention to have its intended effect. The interventions were coded from the stage a policy or intervention was confirmed and we did not include details of policy development.

##### Methods

###### Extract descriptive information

Where available, we used the intervention name as the intervention identifier throughout this step. Where this was not available, the identifier was the first author and year of the index paper. For each intervention, we extracted descriptive information into an excel spreadsheet (citation for all linked papers; target behaviour; country; setting; study population). We read and reread each intervention collection for familiarisation with the content prior to the coding and analysis.

**Coding actors and actions involved in each intervention**

We coded all papers and reports included in the intervention collection. We analysed whole papers or reports as opposed to verbatim intervention descriptions, as relevant information was often included in other sections. We coded all actors and actions, where the *actor* refers to 'who' has to conduct an action for the intervention to have an effect on diet or PA. They could be an individual, group, or organisation required to act for an intervention to exert its intended effect. The *action* was defined as what the actor was required do to the achieve the intended intervention effect. We coded all actions that occurred from the point at which it was confirmed a policy or intervention would happen as we were interested in the implementation process as opposed to the policy process. Only actions and actors explicitly described in the intervention collections were coded. Two reviewers coded an initial 10 interventions and met to discuss the coding and create an initial coding framework. One researcher completed the coding for the remaining intervention collections, using the existing codes where they are applicable. Where required, new codes were created or existing codes refined.

#### Results

All 74 intervention collections were coded. An example of the coding is included in table 1.2.

**Table 1.2** – *Example of intervention coding for 'Get Up and Do Something'[8-10]*

| Actor | Action | Supporting text [a] |
|---|---|---|
| State of Delaware Division of Public Health | Allocate tobacco funds to fund campaign costs | The State of Delaware allocated tobacco settlement funds to address this problem through the development and implementation of a social marketing campaign to promote PA among adolescents aged 12 to 17 years.[9] |
| Research team | Develop and implement TV billboards and bus wrap adverts | The State of Delaware earmarked tobacco settlement funds to address this problem through the development and implementation of a social marketing campaign to promote PA.[8] |
| | Expose target individuals to campaign by selecting correct TV channels and high visibility billboard areas | The television ad placement was limited to the cable network channels E!, MTV, BET, FX, ESPN2, and Comedy Channel. These channels cater to the 18 to 30 year old target market and were exclusive to Delaware residents via the cable network system.[8] |
| | Organise for advert to be placed on BusWrap | A bus wrap was painted on two Delaware Area Rapid Transit buses that were limited to New Castle County—the most populated of the three counties in Delaware (64% of total state population). The campaign ran for a 13-week period starting the latter part of September through December of 2001.[8] |
| | Run media campaign for 6 weeks of TV and billboards | The Get Up and Do Something media campaign ran for 6 weeks during the months February and March in the calendar year 2004.[9] |

| | | |
|---|---|---|
| Adolescents | See campaign adverts | 24.9% of those surveyed had seen either the billboard or the bus wrap.[10] |
| | Recall seeing the bus advert | The ad campaign was considered to have the ability to maintain recollection.[8] |
| | Interpret campaign messages correctly | The campaign message was correctly interpreted.[8] |
| | Change perceptions of PA | The emphasis of the ad was to affect the target market's perception that PA was fun and a good way to socialize—that it did not need to be done in a gym or involve pain.[9] |
| | Be motivated to increase PA | The emphasis of the campaign was to positively affect the target market's perception of PA and to motivate them to get up and do something physically active.[9] |
| | Consider being more active | The goal was to get the audience to consider being more active.[9] |
| | Set intentions to become more active | 31.2% indicated that as a result of seeing the specific Get Up and Do Something television ad they intended to be more active.[9] |
| | Engage in PA | 44.1% reporting becoming more active.[9] |
| | Talk to family member, co-worker or friend about the advert | 25.5% had talked to someone such as a family member, co-worker or friend about the ad.[8] |

[a] Single supporting quote provided to illustrate coding examples.

#### Step 1c. Synthesise data and develop framework

*Synthesise coding from step 1b and develop a draft framework*

##### Summary

*Aim:* To synthesise coding from step 1b of actors and actions into a draft framework.

*Methods:* The intervention coding contributed to the development of schematic flow charts relevant to single intervention components, describing the common sequence of actors and their actions required for the intervention component to exert its effect. We created the flow charts using one intervention and iteratively refined them by integrating similar intervention types into a schematic. The refined flow chart was checked against the previous interventions that contributed to its development. We repeated this process until a final intervention flow chart was produced. Where similarities between schematic flow charts were identified, they were combined to form overview schematics. We used the overview schematic flow charts to identify conceptually similar categories of actors and actions. We conducted iterative data clinics with the full research team to test and refine emerging categories and organisation.

*Results:* The resulting framework was a 3x5 matrix, identifying actors on the y axis (macro-environmental actors, micro-environmental actors and informal gatekeepers) and 5 categories representing the agentic demand on the intervention recipient on the x-axis. The placement of the intervention on the x-axis was determined by how recipients were exposed to the intervention, how they were required to engage with the intervention and what the aim of the intervention was. The category was determined by completing two decision trees that determined the location of the intervention component within the matrix.

#### Methods

##### Developing schematic descriptions

Using the codes assigned for each intervention collection, we created an intervention schematic flow chart of the actors and their actions required for the intervention to exert its intended effect. This included implicit actions that are not described within the text, but may be inferred or known by the review team based on their knowledge of the evidence base. One researcher who conducted the coding developed intervention schematic flow charts, which were discussed and refined with a second reviewer and the wider review team. We developed intervention schematic flow charts for single intervention components, and thus multi-component interventions could contribute to multiple schematics. Multiple interventions that included similar components contributed to refine the schematic, eg, multiple menu labelling interventions contributed to one schematic flow chart description. Refined intervention flow charts were checked against previously coded interventions to ensure they remained accurate. We ceased coding interventions when new coding was no longer contributing to flow chart refinement. In an iterative process, intervention schematics were merged into overview schematic flow charts for similar intervention types and checked against previously coded interventions.

##### Conceptual framework development

We compared the schematics to identify key constructs relating to actors and their actions and, attempted to organise and order them. This process was initiated by the researcher who created the schematic flow charts, and data clinics with the research team were used to test and refine emerging categories and organisation. This process was conducted iteratively throughout the coding and schematic development process.

#### Results

The interventions initially contributed to 55 intervention schematics (example provided in figure 1.3), which were combined to form 8 overview schematics (Information training/provision; Improve healthy food availability; Provision of PA infrastructure; financial assistance; Legislation; Food reformulation; Intervention design; Self-regulatory interventions). An example of how initial intervention schematics were merged to contribute to overview schematic flow chart, in this case the provision of education, training or information is provided in figure 1.4, and the final schematic is included in figure 1.5.

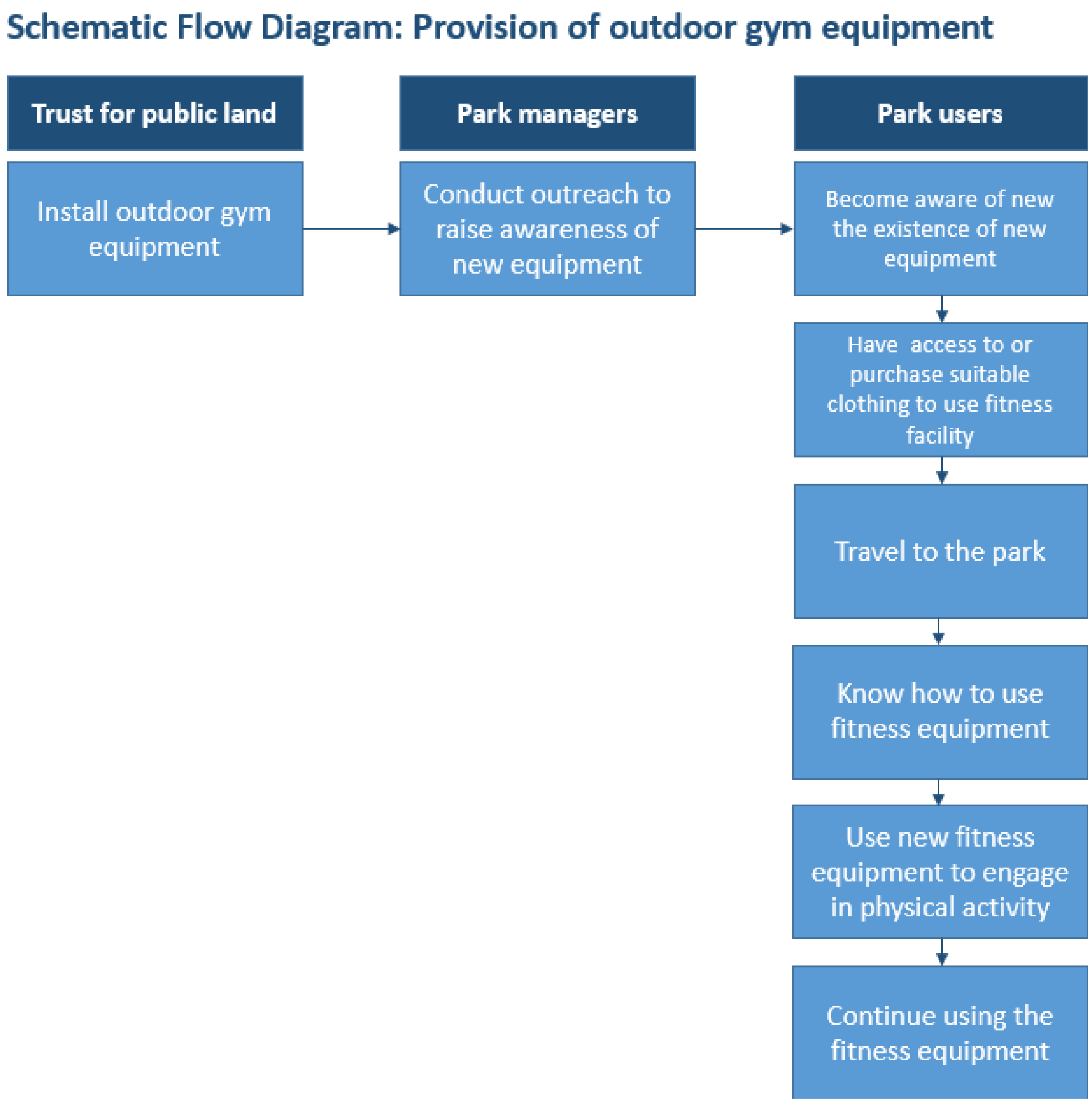


**Figure 1.3** – *Example of initial intervention schematic 'Provision of outdoor gym equipment'*

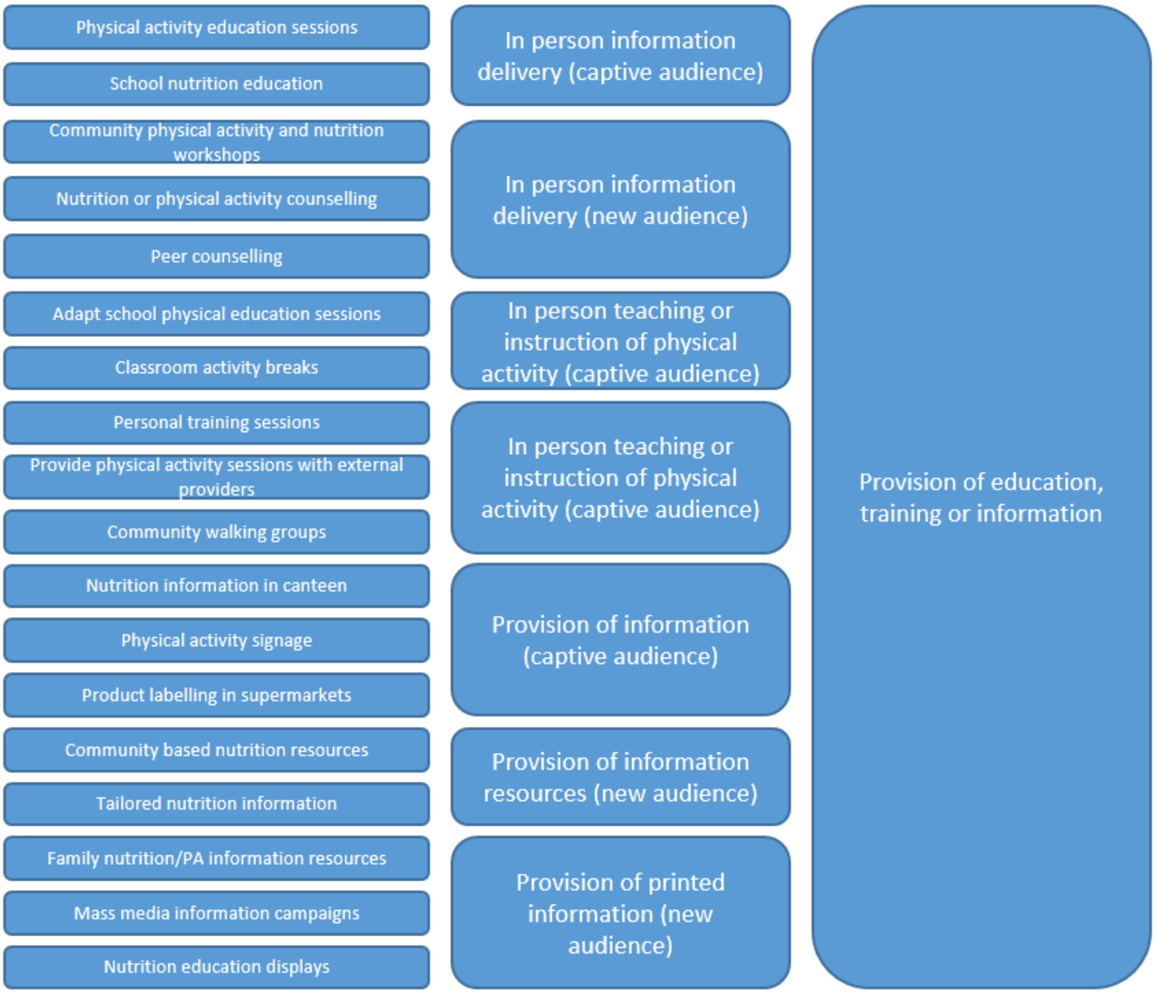


**Figure 1.4** – *Iterative development of intervention schematics*

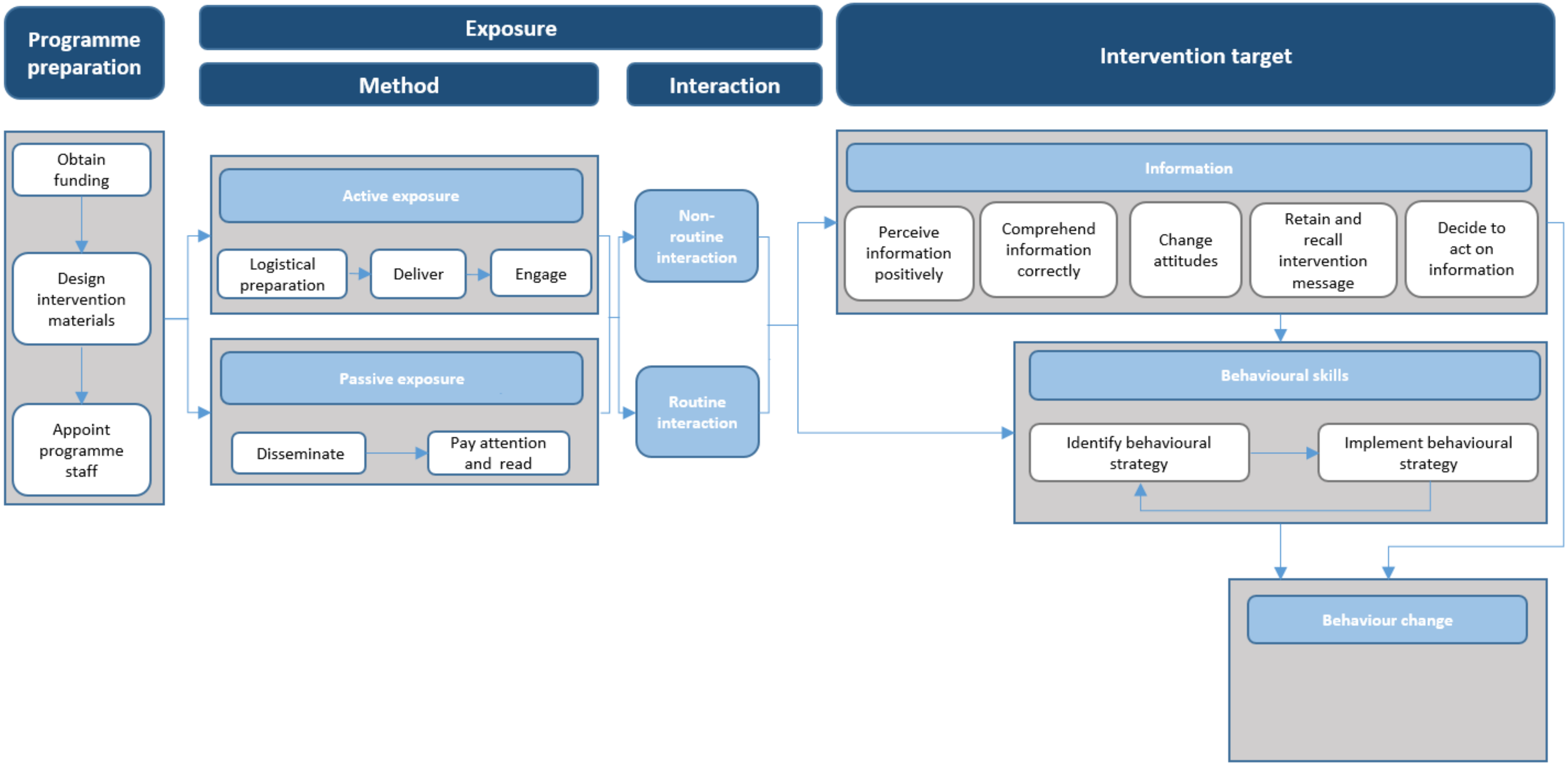


**Figure 1.5** – *Final intervention schematic for provision of training, education or information*

**Draft framework**

When we compared the eight overview schematic flow charts, we identified constructs that differentiated intervention agency demand. We developed a conceptual diagram underpinning these constructs and a framework and application tool categorising interventions according to these constructs. The briefing document, summarising the diagram, framework and application tool is included in Supplementary Appendix 1.5. We do not describe this in detail here because we revised the framework during future steps to achieve a final version. Rather, below is a brief description of the framework structure and the key constructs identified that remain a feature of the final framework.

The resulting framework was a 3x5 matrix (Figure 1.6), identifying 3 categories of actors on the y-axis. These are:

1) Macro-environmental actors – an individual or group of individuals representing an industry, service or supporting infrastructure that influence dietary or PA behaviour.
2) Micro-environmental actors – actors within micro-environmental settings with authority to decide whether, or direct how, an intervention is implemented or an actor with responsibility to implement the intervention. Micro-environmental settings are those that are places or naturally occurring groups of places where people gather for specific purposes.
3) Informal gatekeepers – an individual required to change their behaviour to influence dietary or PA practice of the primary recipient.

The 5 categories on the x-axis represent the agentic demand on the intervention recipient. The placement of the intervention on the x-axis is underpinned by three constructs (1) Exposure; (2) Engagement; and (3) Intervention Target. While these constructs underpin the x-axis of the matrix, this is not transparent in the draft framework, however, they remain a feature of the final framework and become clearer in future iterations.

Collective agency

| Environmental agency demand | | | | | | | |
|---|---|---|---|---|---|---|---|
| | **Higher resource demand** Interventions that require individual level actors (gatekeepers) | **A** | Policy that requires parents to provide healthy snacks at school **A1** | Low emission zones **A2** | **A3** | Financial assistance for healthy food purchasing **A4** | After school nutrition counselling for parents. **A5** |
| | **Medium resource demand** Interventions that require micro-environmental actors | **B** | Modify PE lessons to increase MVPA time **B1** | Point of purchasing labelling in supermarkets **B2** | Community based PA classes **B3** | Workplace information and motivational intervention **B4** | Community-based physical activity counselling **B5** |
| | **Lower resource demand** Interventions that require macro-environmental actors | **C** | Food reformulation **C1** | Changes to whole city transport networks **C2** | Public bicycle hire schemes **C3** | Mass media information campaigns **C4** | Recipe guides on health agency websites **C5** |
| | | | **1** | **2** | **3** | **4** | **5** |
| | | | Directly targets behaviour change; or targets intermediate steps via routine interactions and non-conscious processes | Targets intermediate steps via routine interactions and conscious processes | Targets intermediate steps or direct behaviour change via non-routine interactions | Targets individual resource via routine interactions | Targets individual resource via non-routine interactions demand |
| | | | **Distribution of agency demand** | | | | |

**Collective agency**

**Figure 1.6** – *Draft framework demonstrating intervention actors (Y axis) and agency demand on individuals (x-axis)*

### Step 2: Testing the DePtH framework

#### Step 2a: Seek qualitative feedback

*Recruit academic and policy experts to assess the validity and utility of the draft framework*


##### Summary

*Aim:* To explore the content validity and practical utility of the draft framework.

*Methods:* Four workshops were conducted with invited experts (n=20) with experience of developing, implementing, evaluating or synthesising evidence of PHIs to promote diet and PA. Participants predominantly worked in the academic sector (85%). The disciplinary background of the majority of participants was public health (80%). We circulated the draft framework in advance. During the workshop, participants applied the framework to six intervention examples and took part in a structured discussion. Workshops were audio-recorded and detailed field notes were taken.

*Results:* Feedback from the workshop was grouped into three main categories: inconsistent and poorly defined terminology; Unintuitive framework structure that was not clearly linked to theory; Unclear user instructions.

*Subsequent actions:* Based on the workshop findings, we refined the framework. We sought further feedback from selected workshop participants via email and virtual meetings and made further minor changes.


##### Methods

###### Participants

We invited experts with experience of developing, evaluating or synthesising evidence of PHIs to promote diet and PA to one of four online workshops with an aim to recruit up to 30 experts in groups of 5-15 participants per workshop. We sought to include a range of perspectives, including by country of location, career stage, disciplinary background, sector, and whether experience is primarily in development, evaluation or synthesising evidence of PHIs.

Inclusion criteria for online workshop participants were working within the academic, policy or third sector; experience in the development, implementation evaluation or synthesis on population diet and/or PA interventions; and willing and able to take part in the research. We initially developed a longlist of potential participants to reflect the range of perspectives we were seeking, identified by (1) project team networks; (2) authors of interventions included in Step 1a; and (3) Internet searches. We populated a spreadsheet of potential participants and, where known, indicated their research focus (diet/PA), country of location, career stage, disciplinary background, sector, and whether experience was primarily in development, evaluation or synthesising evidence on PHIs. We prioritised potential participants using the coding classification ‘must ask’, ‘should ask’, ‘could ask’, ensuring the highest priority group included a breadth of perspectives. We first invited participants in the highest priority group.

###### Recruitment

Figure 1.7 presents the recruitment process. Participants were invited to take part in the workshop via an email invitation that included a written participant information sheet and opportunity to ask questions via email or phone before making a decision to attend. Reminder emails were sent if no response was received within 1-2 weeks, or in line with an automated response indicating respondent unavailability. Potential participants were considered non-responders if no response was received within 3 days of the reminder email.

We sent e-consent links to participants who indicated they were interested in participating to complete ahead of the workshop. A reminder to complete the e-consent was sent three days prior to the workshop. The hyperlink to the zoom call was circulated to participants upon receipt of a completed e-consent form.

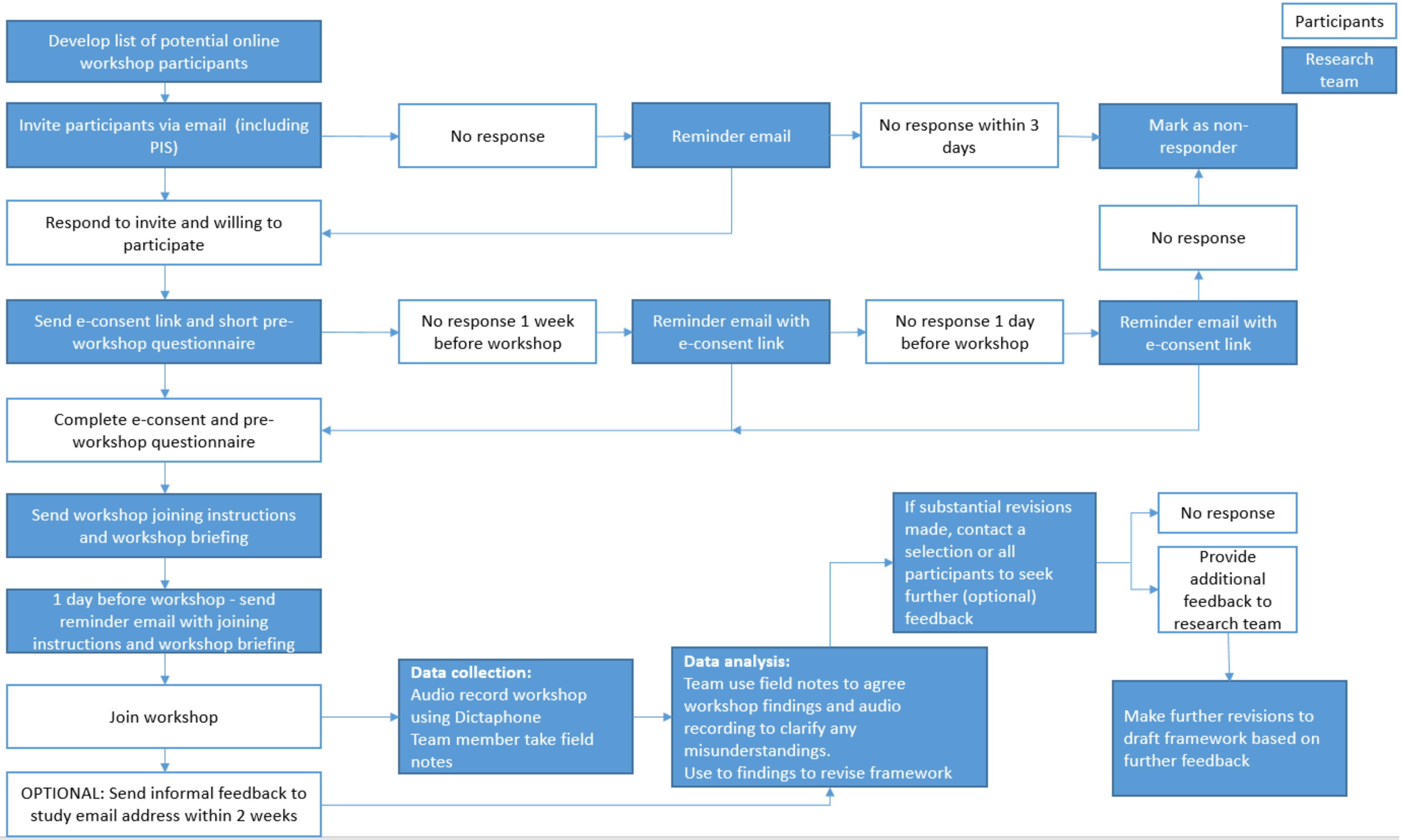


**Figure 1.7 –** *Recruitment and data collection process for step 2a*

##### Data collection

###### *Preparation*

Prior to the workshop, participants completed an online survey about their professional background (country of location, career stage, disciplinary background, sector, and whether experience is primarily in development or evaluation of PHIs, and primarily in diet or PA). Participants received a pre-workshop briefing document that included an explanation of the framework development process, the framework and pre-selected intervention descriptions (Appendix 1.5).

###### *Online workshop*

Three members of the project team and 5-10 participants attended each online workshop. One member of the project team facilitated the workshop, which included introducing the framework and facilitating the discussion. Two members of the team wrote field notes, summarising the main discussion points in the workshop. We ran online workshops until we deemed that we were no longer receiving new feedback from participants. In order to maintain a manageable number of participants within each workshop and provide flexibility for participants we ran four workshops, lasting a maximum of 2 hours.

*Introduction:* During the introduction, we (1) introduced the research team; (2) recapped the study participation information, including possibility of withdrawal, participant confidentiality; (3) requested that participants abide by simple ground rules including placing mobile phones on silent, using the mute function when others are speaking; (4) recapped the purpose of the workshop and shared an agenda for the workshop, which involved three further parts.

*Part 1:* A practical session where participants applied the framework to a pre-selected set of interventions. A member of the research team briefly introduced and provided instructions for the practical task. Participants were given the opportunity to ask any clarification questions ahead of the practical exercise. We selected the interventions [8, 11-15] from those identified in Step 1a representing a variety of intervention types and verbatim intervention descriptions were selected from the journal articles and reports collected in step 1a. Participants were given 20 minutes to apply the framework to the pre-selected intervention examples. The task aimed to provide experience to the participants on the practical application of the framework.

*Part 2*: involved a guided discussion with all workshop participants, facilitated by one member of the research team. The facilitator guided the discussion through a series of questions that aimed to explore the content validity and practical utility of the framework (Table 1.3) and the facilitator asked clarifying and follow up questions to participants. Participants were asked to think about the previous exercise and their experiences of delivering, implementing, evaluating or synthesising evidence on PHIs when responding to the questions. The facilitator sought other participants' opinions on topics raised throughout the discussion. Part 2 was curtailed when useful discussion dried up, or when the whole workshop lasted 2 hours – whichever occurred first. Participants were given an informal opportunity to provide further feedback or reflections on the framework and its application after the workshop by emailing these to the study email address.

**Table 1.3 –** *Questions to guide Part 2*

| | |
|---|---|
| ***Content validity*** | To what extent do you feel the framework captures the underlying phenomena of the agentic demands of PHIs? |

| | |
|---|---|
| | Are there any concepts relating to the topic that are not captured, or under-represented in the framework?<br><br>How much overlap were there between concepts and how did this affect the application of the framework?<br><br>Does the framework place too much emphasis on any particular concept? |
| ***Practical utility*** | What were your experiences of applying the framework to the intervention examples?<br><br>How do you see the framework being utilised?<br><br>Are there any changes that could be made to improve the applicability of the framework? |

The workshops took place in October-November 2021. To minimise the potential for pandemic related disruption and be sensitive to participants' desire to socially distance, workshops took place via zoom. This also maximised value for money, minimised time and travel demands on participants. We used Microsoft PowerPoint to introduce the workshops and provide instructions to participants. Two members of the research team took field notes. The data from the workshops consisted of an audio recording of each workshop, field notes and informal written feedback provided by participants after the workshop. Audio recordings were not be transcribed and only referred to if clarification during revisions to the framework was required.

###### *Framework refinement*

We used workshop feedback to revise the framework and discussed the changes in a project meeting. Given we made substantial changes to the draft framework, we re-contacted purposively selected workshop participants and asked them to provide additional further verbal feedback in informal interviews.

##### Results

We invited 61 potential participants, of which 20 participated (Figure 1.8). We conducted 4 workshops in October and November 2021 (Workshop 1 (n=4); Workshop 2 (n=5); Workshop 3 (n=5); Workshop 4 (n=6)). Participant characteristics are described in table 1.4; they predominantly worked in the academic sector (85%) and the majority described their background as public health (80%). While our aim was to recruit 30 participants, we found that after four workshops, no new findings were emerging and we curtailed data collection.

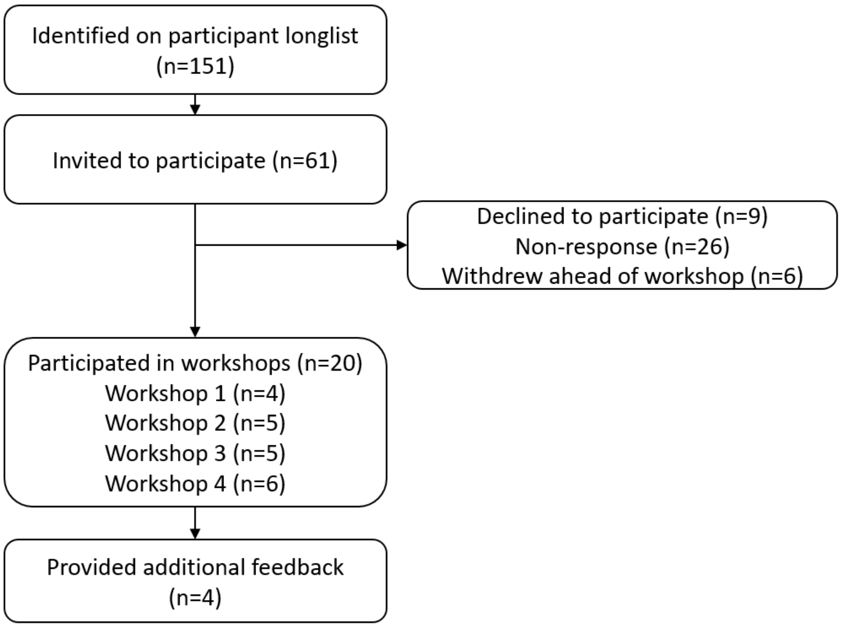


**Figure 1.8 –** *Participant recruitment to online workshops*

**Table 1.4** – *Participant characteristics*

| | **n (%)** |
|---|---|
| **Country where research occurs** | |
| Australia | 2 (10) |
| United Kingdom | 17 (85) |
| Canada | 1 (5) |
| **Professional sector** | |
| Academic | 17 (85) |
| Third Sector | 1 (5) |
| Policy | 2 (10) |
| **Disciplinary background** | |
| Public health | 16 (80) |
| Health economics | 1 (5) |
| Psychology | 2 (10) |
| Health services research | 1 (5) |
| **Focus of research** | |
| Dietary | 10 (50) |
| PA | 2 (10) |
| Both | 8 (40) |
| **Years of professional experience** | |
| Less than 5 years | 3 (15) |
| 6-10 years | 8 (40) |
| 11-20 years | 5 (25) |
| More than 20 years | 4 (20) |
| **PHI experience** | |
| Developing PHI's | 10 (50) |

| | |
|---|---|
| Implementing PHI's | 5 (25) |
| Evaluating PHI's | 15 (75) |
| Synthesising PHI's | 16 (80) |

**Workshop feedback and subsequent action**

We grouped participant feedback into three categories, presented below alongside details of the research teams' response and subsequent changes to the framework. The three categories of feedback were: (1) Terminology and categorisation; (2) Framework structure; and (3) User instructions.

*(1) Terminology and categorisation*

Participants highlighted the importance of ensuring that terminology was appropriate across disciplines, noting that in some cases, the framework used terminology that held specific definitions within specific disciplines, which were not being used appropriately. For example, the terms macro-environment and micro-environment have very specific meanings within health economics and conscious and non-conscious and routine have very specific meanings in psychology. As a result, the terms conscious and non-conscious and non-routine and routine were replaced with active and passive to describe exposure and engagement with the intervention. We made no change to the use of the terms macro- and micro-environmental actors because we based these terms and their definitions on an existing framework accepted and widely cited in the public health literature.[16]

Some participants discussed the use of the term 'agency' in the workshops. While there was agreement it was being used in the right way, for accessibility some participants recommended the use of an alternative term. This is because agency has a specified meaning particularly within Sociology that may not align with how we use it here. Furthermore, to people who are not familiar with the concept of agency as we use it here, it is not necessarily an easily accessible concept. We were aware of this and had previously sought views from colleagues on alternative terminology. Unfortunately, no other options seemed appropriate, and given its use as a term across public health literature, we deemed it the least worst option to continue use 'agency'.[17]

Within the draft framework, we proposed that the agentic demand of interventions varied by what we termed the intervention target - that is how an intervention aims to change behaviour. One category proposed as an intervention target was interventions that aim to change the resource available to recipients in order to facilitate behaviour change. Within this, resources included financial, cognitive, social or material resource. Feedback from workshop participants indicated that the agentic demand of intervention types included within the 'resource' category were too heterogeneous. For example, interventions that aim to change financial resource, eg, food vouchers and interventions that aim to change cognitive resource, eg, information about how to use fresh produce to cook a healthy meal are not equivalent in their agentic demand. While they may still require substantial action from recipients to implement the intervention, an intervention to improve financial resource aims to enhance the individuals' agentic capacity in a way that is not equivalent to increasing knowledge. This led to a substantial change to the framework. Rather than the 'intervention target', we refocused on mechanisms of action. These was categorised into socio-cultural, cognitive, financial, physical environmental and biomedical.[16]

One participant also noted that the draft framework was not able to address interventions that operate via multiple mechanisms of action (which may have difference agentic demands on recipients). For example, calorie-labelling interventions known to operate via two mechanisms (1) to inform consumers of calorie content and to prompt a change in purchasing behaviour, and (2) incentivise food manufacturers to reformulate their products. As a result, we changed the framework to allow for multiple mechanisms of action by which interventions operate.

Participants noted difficulty categorising the constructs, which inherently lie on continua, into binary categorisations, yet participants also recognised the necessity of this for a classification system. Thus, rather than make further changes to the framework, we recognised this challenge in the guidance.

*(2) Framework structure*

The draft framework and its accompanying application tool aimed to streamline the process of identifying an intervention's location on the matrix. However, feedback indicated that this process was not transparent, particularly in relation to actors. In particular, the draft framework skipped some questions, for expediency, and participants did not think this was intuitive. Participants also found it confusing to code actors and individual agency on the same matrix. In response to both of these comments, we ensured that all questions were asked of all interventions during coding.

While the framework instructed participants to apply the framework to single intervention components, participants noted that this would be better as a stand-alone step. Therefore, we added an early step of identifying all intervention components.

*(3) User instructions*

Participants suggested some additional user instructions that could be included to make the application process simpler. As well as identifying all intervention components we also added an early step to identify the recipient group.

**Further feedback**

We sought feedback on the revised framework from eight workshop participants, selected due to their contributions during the initial workshops. Four workshop participants agreed to provide verbal feedback in an online meeting with two members of the project team. The revised framework was circulated to these participants ahead of the feedback session, which included an explanation of the revised version, a worked example and a blank example for the participant to either complete ahead of, or during, the feedback session. During the feedback session, participants were asked to complete a think-aloud exercise while applying the framework to an intervention example. Following the feedback sessions, minor changes were made to application instructions and terminology ahead of step 2b. The changes made to the framework were:

- Separation between socio-cultural and cognitive mechanisms of action as participants noted that these categories overlapped. This change placed the focus of the cognitive category on individual changes, eg, to attitudes , beliefs etc, because it was not possible to separate individual beliefs (previously categorised as socio-cultural) and knowledge (previously categorised as cognitive). The socio-cultural category thus focused on changes to society and community including changes to wider social structures.

All changes described in this stage led to the development of the final framework (Supplementary material 2). While some of the application guidance was changed in subsequent steps, the core framework remained the same.

#### Step 2b. Reliability assessment

##### Summary

*Aim:* To assess consistency of intervention coding using the final DePtH framework via an online survey.

*Methods:* We recruited a new sample of academics working in the area of PHIs (n=22) to code 53 intervention examples according to the final DePtH framework, using a bespoke online survey. Two participants independently coded each intervention example. We calculated Cohen's Kappa for each categorical framework construct to assess inter-rater reliability.

*Results:* When identifying intervention components, participants agreed or marginally agreed on 75% of intervention components and disagreed on 25%. Of the categorical framework classifications, Cohen's Kappa classed two constructs as moderate agreement (Mechanism of action; macro-environmental actors), two constructs as fair agreement (exposure; engagement), one as none to slight agreement (informal gatekeepers) and two as no agreement (micro-environmental actors; secondary recipients). Due to the poor agreement, we explored inter-rater reliability further in step 3.

##### Methods

###### Participants

We recruited a new sample of academic experts with experience of developing, implementing, evaluating or synthesising PHIs for diet and/or PA.

###### *Sample size*

We used the KappaSize R Package[18] to calculate the number of intervention descriptions required in the survey, as has been previously used to calculate sample size for inter-rater reliability.[19] We estimated a sample size using the following parameters; alpha value of 0.05; power of 0.80, using 2 coders, a null hypothesis of a kappa of 0.4 and expected kappa of 0.7. This calculation suggested a sample size of 53 intervention descriptions would be required to test whether the kappa value exceeds 0.4, the lower bound of substantial agreement when interpreting the kappa value. The 53 intervention descriptions were purposively sampled from the interventions identified in WP1 and used in WP2 to develop the framework. We selected interventions that represented a balance across the framework classifications ensuring that all levels of each classification were present in the sample.

Given we were seeking to assess inter-rater reliability between two participants for each intervention example, 106 assessments were required – i.e. two of each description. Assuming that each participant would complete 5 intervention examples, we aimed to recruit 22 participants to assess 53 intervention examples.

###### *Recruitment*

Given the relatively small number of participants required, we circulated information about the survey in the first instance to relevant research groups within the MRC Epidemiology Unit. The research groups had no previous knowledge of the draft and final framework. The chosen groups include researchers with expertise in PHIs for diet and PA across a range of disciplinary backgrounds and career stages. To recruit further participants, we circulated information via the International Society for Behavioural Nutrition and PA and the Public Health Policy Research Unit.

We emailed potential participants asking them to respond within 14 days if they were interested in participating, and a reminder email was sent after 10 days. We sent interested participants the survey link via email, which directed them to complete a consent form. Once participants completed the consent form, they were automatically directed to the online survey. The email also included a PDF document containing five intervention examples to use in the survey.

**Data collection**

The online questionnaire asked participants to complete questions about their professional background, including: country, sector, discipline, and experience. Participants were asked to download a briefing document, which included a short introduction to the project and a description of the final DePtH framework. We recommended that participants read the document ahead of completing the survey and referred back to the document while completing the survey.

For each of the intervention examples allocated to each participant, the survey required them to complete questions based on steps 1-3 of the framework (Figure 1.9). Application Stage 1 asked participants to identify all intervention components, followed by all intervention recipients. The survey then automatically cross-tabulated answers to generate a list of intervention component-recipient combinations. For each combination, participants were asked to complete Application Stage 2 to identify the agentic demand of each combination by categorising exposure to the intervention, the mechanism of action and the required engagement. The final Application Stage 3 required participants to identify additional classes of actors involved in the intervention. At each stage, we encouraged participants to provide supporting evidence to explain their decision and describe difficulties encountered when applying the framework.

The survey was be hosted on a secure web form developed by the Data Management team at the MRC Epidemiology Unit.

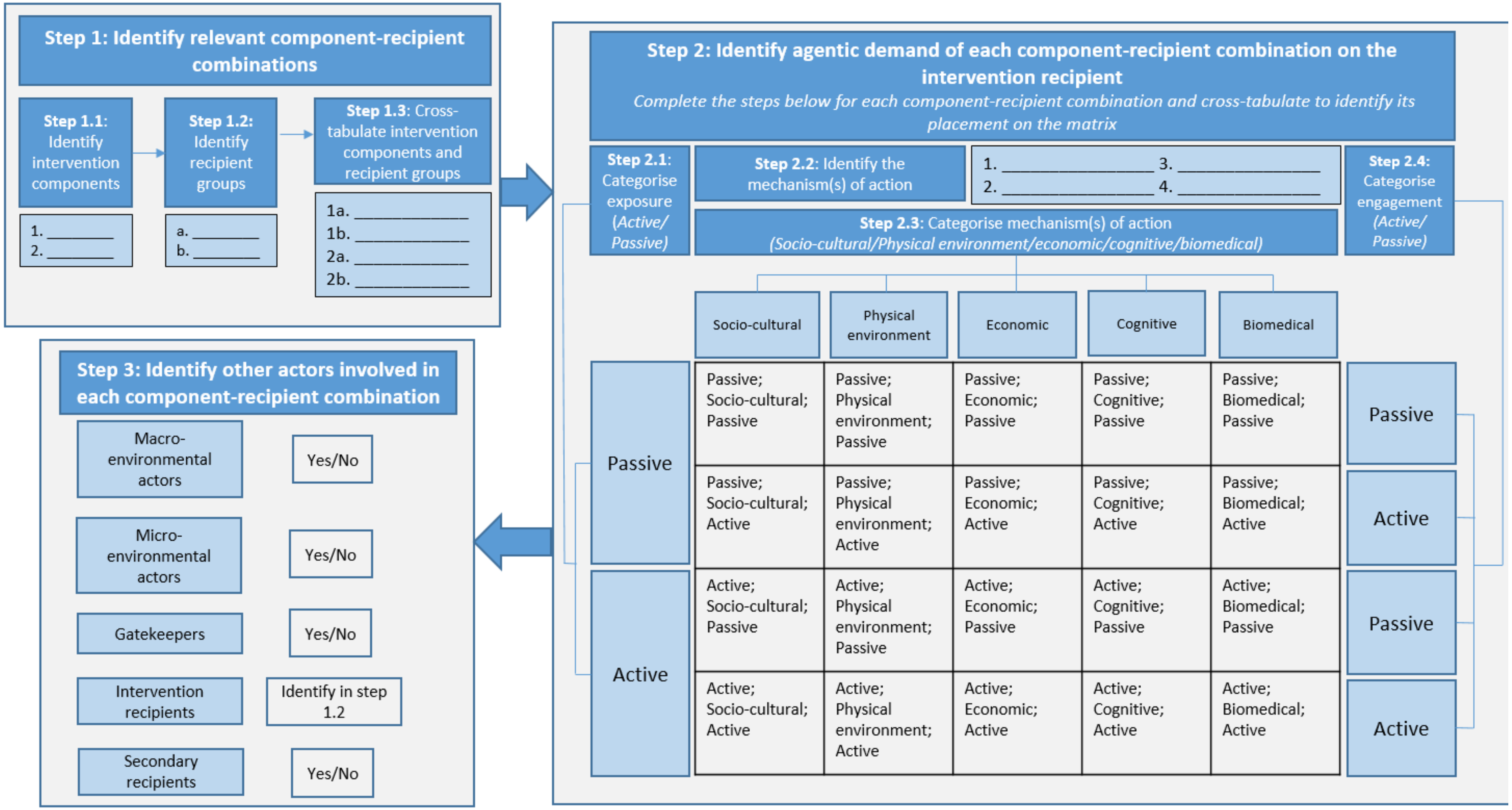


**Figure 1.9 –** *Overview of the framework application process*

**Data analysis**
We firstly compared participants' identification of intervention components. For each intervention, we assessed participants free text responses and justifications for identifying single intervention components and matched these between participants. We classified their agreement according to three categories:

1. *Agree* – Two participants' descriptions and justification refer to the same intervention component. For example, participant A – Exercise opportunities at community centre; Participant B – Exercise Opportunities.
2. *Marginal agreement* – Participant A and B identify similar intervention components but they have categorised them so that it is not possible to match them due to differences in level of detail. We speculate that the two participants would have agreed if they had had the opportunity to discuss. For example participant A identifies '*Education intervention on improving knowledge, skills and food choices'* as a single component. Participant B identifies '*Improve knowledge'* and '*Improving skills'* as two intervention components.
3. *Disagree* – One participant identified an intervention component that was not possible to match with components identified by second participant. For example, participant A identified '*Improve quality of cycle paths',* but this was not identified by participant B.

We analysed inter-rater reliability of the DePtH framework for intervention components with agreement between participant a and participant b. It was not possible to calculate inter-rater reliability for components with only marginal agreement or disagreement because participants did not make the classification based on the same component.

For each intervention component that was agreed by both participants, we calculated Cohen's Kappa for each framework item classification (exposure, mechanism of action, engagement, macro-environmental actor, micro-environmental actor, gatekeeper, secondary recipients). We interpreted the kappa values according to standardised categories ≤0 no agreement; 0.01-0.2 none to slight; 0.21-0.40 fair; 0.41-0.60 moderate; 0.61-0.8 substantial and 0.81-1.00 as almost perfect agreement.[20]

#### Results

##### *Intervention components*

From the 53 intervention examples, participants identified 157 intervention components. Of these, there was agreement on 58% (n=92), marginal agreement on 13% (n=20) and disagreement on 29% (n=45).

##### *Framework categories*

Table 1.5 presents Cohen's Kappa statistics for each DePtH framework item. The total number of observations is lower than the number of intervention components as it relates to component-recipient group combinations deemed applicable by participants.

**Table 1.5 –** *Statistical inter-rater agreement for individual framework items.*

| | | Analysis | | Sensitivity analysis* | |
|---|---|---|---|---|---|
| | Observations (n) | Kappa value | Interpretation | Kappa value | Interpretation |
| **Exposure** | 85 | 0.29 | Fair | 0.38 | Fair |
| **Mechanism of action category [a]** | 100 | 0.57 | Moderate | 0.63 | Substantial |
| **Engagement [a]** | 100 | 0.23 | Fair | 0.27 | Fair |

| | | | | | |
|---|---|---|---|---|---|
| **Macro-environmental actors** | 85 | 0.49 | Moderate | 0.21 | Moderate |
| **Micro-environmental actors** | 85 | -0.05 | No agreement | -0.10 | No agreement |
| **Informal gatekeepers** | 85 | 0.20 | None-slight agreement | 0.24 | None-slight |
| **Secondary recipients** | 85 | -0.04 | No agreement | -0.01 | No agreement |

[a] Number of observations greater than other items because participants can identify multiple mechanisms of action within intervention components.
*After removal of two participants from analysis. One who self-identified as completing in a rush and one whose answers indicated they had not understood the framework.

We *a priori* identified an acceptable kappa value of 0.6 for this project and this was reached for only one framework item – mechanism of action. The remaining framework items had kappa values within ranges of moderate (Macro-environmental actors), fair (Exposure and engagement), none to slight (Informal gatekeepers) and no agreement (Micro-environmental actors and secondary recipients). Using free text responses we found some key areas responsible for poor agreement are outlined in table 1.6.

**Table 1.6 –** *Explanation for key discrepancies between participants*

| **Framework construct** | **Explanation for poor inter-rater agreement** |
|---|---|
| Exposure | Some discrepancies occur from one participant misunderstanding the difference between exposure and engagement.<br>Other participants did not sufficiently distinguish different recipient groups, assuming the same exposure rating for all recipient groups.<br>Other discrepancies relate to lack of clarity in intervention reporting where users made different best estimates of the exposure. |
| Mechanism of action | The key discrepancy was distinguishing between cognitive and socio-cultural mechanisms.<br>Further, some participants did not make use of the facility to identify multiple mechanisms of action. |
| Engagement | Participants did not provide sufficient justification to be able to discern reasons for disagreements. |
| Actors *(Macro-environmental; micro-environmental; Informal gatekeepers; secondary recipients)* | Participants often identified the same actors within interventions, yet classified them differently, particularly noted discrepancies between macro-environmental and micro-environmental actors, and between micro-environmental actors and informal gatekeepers. For example, schools, companies, community groups classified as informal gatekeepers not as micro-environmental actors. |

#### Implications

We found that inter-rater reliability was low. However, we note that the study design did not allow an opportunity for piloting or discussion of discrepancies which are processes that would normally take place in a systematic review. Therefore, we explored inter-rater reliability further in the step 3, taking account of the key reasons for disagreement as presented in table 1.6.

#### Step 3: Applying the DePtH framework

*Apply the final DePtH framework within a 'proof-of-concept' systematic review exploring the association of intervention agentic demand with intervention effectiveness and equity*

##### Summary

*Aims:* Apply the final DePtH framework within a 'proof-of-concept' systematic review to explore the relationship between intervention agency demand and intervention effectiveness and equity.

*Methods:* We adopted a 'proof-of-concept' approach to assess the application of the final DePtH framework within a systematic review. We identified three systematic reviews assessing the equity effects of dietary and PA PHIs and extracted and screened interventions included in them. We included PHIs for diet and PA that presented results overall and differentiated by a measure of socioeconomic position (SEP). We coded DePtH classification for eligible interventions and extracted data on overall and differential effectiveness on diet and PA outcomes. We present results in a harvest plot.

*Results:* 31 interventions met the inclusion criteria. There was enough information to apply the DePtH framework to 26 of these, including 163 intervention component-recipient combinations. We identified all three framework constructs for 115 intervention components. The most frequent framework classification was passive exposure – cognitive mechanism – active engagement and in contrast we found no intervention examples within nine framework classifications. It was only possible to draw tentative conclusions due to the small number of components in some framework classifications. Three intervention component-recipient combinations appeared to be associated with reduced socioeconomic inequalities. Most intervention components did not appear to have an influence on socio-economic inequalities, but there were a considerable number of component-recipient combinations that targeted cognitive mechanisms of action that appeared likely to widen socioeconomic inequalities.

##### Methods

We originally conceived this stage as a systematic review to explore the associations between intervention agentic demand and intervention effectiveness, (socio-economic) equity and acceptability. Our intention was to conduct a systematic search for articles evaluating intervention effectiveness, identify linked papers (see step 1) and use these linked papers to identify data related to equity and acceptability. However, during step 1, we found that few linked papers included equity and acceptability data. Such data may not exist or may only be accessible via author requests.[21]

As an alternative, we applied the DePtH framework to studies included in existing systematic reviews exploring differential socio-economic effects of PHIs with dietary and PA outcomes. As well as allowing us to efficiently identify data on differential effects by markers of SEP (equity), we assumed that included papers in these reviews would also report data on overall effectiveness. Hence, we are able to demonstrate the feasibility of applying the DePtH framework within a systematic review and examine the relationships between intervention agentic demands and both intervention effectiveness and equity.

###### Study selection

We used a two stage approach to identify studies for step 3. Firstly, we identified systematic reviews with a focus on differential effects by SEP and from these, we extracted included studies and screened them against our inclusion criteria.

###### *Identifying relevant systematic reviews*

We purposively selected existing systematic reviews that included a collection of primary intervention studies that:

- *include a breadth of intervention types across the framework:* reviews that include a narrow range of intervention types or interventions in a specified population group were not selected because they are likely to occupy a narrow range of the framework limiting our ability to explore patterns of association with effectiveness and equity.
- *present results by a measure of SEP:*
- *include interventions targeting both dietary and PA behaviours.* We preferentially selected reviews that focused on obesity outcomes assuming that studies included in them were likely to include measures of diet or PA.

We purposively selected reviews from the initial systematic search conducted in step 1a. This identified systematic reviews with the potential to include PHIs to promote diet and PA (n=32,306). We searched the titles of these for terms related to equity, based on a validated filter for ethnic and socioeconomic inequalities.[22] Given our aim to demonstrate feasibility in a proof-of-concept study, we did not update the search used in step 1a (conducted in August 2020). We retrieved full texts of reviews identified and purposively sampled a selection that met our criteria.

***Identifying relevant primary studies from included reviews***

From selected equity focused systematic reviews,[23-25] we extracted references for all primary studies, removed duplicates and retrieved full texts. Given primary studies had already been screened for inclusion in the systematic reviews, one reviewer screened all full texts for inclusion and a second reviewer independently screened 50% of the articles. The inclusion criteria are listed below according to the PICO (Population; Intervention; Criteria; Outcome) framework.

*Population*

Interventions that had been evaluated in a (sample of) whole population or population groups as identified by a non-health indicator were included. As in step 1a, non-health indicators are based on the PROGRESS-PLUS criteria;[1] place of residence; race, ethnicity, culture or language; occupation; gender or sex; religion; education; socioeconomic status; social capital; features of relationships; time dependent relationships or personal characteristics associated with discrimination (age, gender reassignment, marriage and civil partnership, pregnancy or maternity, sexual orientation). Interventions evaluated in individuals identified by the presence or absence of a health indicator identified through screening or self-assessment, e.g weight status, confirmed disease risk, confirmed disease or health behaviour were excluded.

*Interventions*

Interventions that met our definition of a PHI (defined above) with the potential to improve dietary quality or increase the frequency or intensity of PA were included. Interventions could target individual behaviours or a combination of behaviours. This includes behaviours beyond diet and PA, eg, smoking cessation. Studies of interventions implemented experimentally or that evaluate local, national or international policy were included.

*Control*

Studies were included if they include a comparison to usual or normal 'treatment' or no intervention.

*Outcome*

Studies were included that had a primary outcome measure of either dietary quality or a measure of frequency, duration, intensity or total volume of PA. Interventions that included only anthropometric measurement or measurements of health not specific to diet or PA were excluded. Studies that include only measures of physical fitness, participants' attitudes, perceptions or motivations, or food purchasing data were excluded. Studies were only included if they also reported differential effects of the intervention by at least one measure of SEP and provide a measure of overall intervention effectiveness in all socio-economic groups combined either in the index paper or in a paper cited in the index paper.

*Study design*
We included randomised controlled trials, quasi-randomised controlled trials, before and after studies, and natural experimental evaluations of the effect of an intervention or policy. Modelling or simulation studies were excluded.

**Data extraction**
Where available, data was extracted from source systematic reviews. Where this was not available, we extracted data from primary studies. One reviewer extracted the study characteristics and outcomes and two reviewers independently extracted intervention characteristics data, including data on the DePtH classification (KG and GV or LB). Disagreements were resolved by a third reviewer (JA).

***Study characteristics***
We extracted publication details, study design, country, setting and the study aims.

***Intervention characteristics***
We used all sections of included papers as the data source for extraction according to the DePtH framework. For each intervention, we identified intervention components and recipient groups and coded each intervention component-recipient combination according to the DePtH framework constructs (exposure; mechanism of action, engagement, macro-environmental actors, micro-environmental actors, informal gatekeepers, secondary recipients). If intervention descriptions did not provide sufficient information to allow reviewers to confidently classify the DePtH framework construct, we recorded 'insufficient information to decide'. Alongside the classifications, reviewers extracted verbatim intervention descriptions and made notes to justify their classification decisions.

All reviewers extracting using the DePtH framework attended an initial meeting to review examples of common disagreements identified in step 2b and agree application rules to address these. The full DePtH guidance is provided in supplementary material 3. Inter-rater reliability was calculated and interpreted as described in step 2b.

***Outcomes***
Outcome data was extracted for dietary and PA outcomes as available. One outcome measure was included from each study for each of diet and PA. Where multiple dietary or PA outcomes were presented, the single outcome that best represented overall or total diet or PA was selected, for example healthy eating score, or total PA was preferred to SSB intake or leisure-time PA. Objective behavioural measures were preferred to self-reported data. When multiple outcome measures, but no measure of overall or total diet or PA behaviour, was presented, the proportion of associations that favoured the intervention, control or no difference was calculated to derive an overall impact for each study, based on the approach taken in a previous review.[25] For example, a study exploring the effect of a free bus pass included three outcomes (Use of active transport, use of buses and walking 3 times per week), all three of which showed a statistically significant effect.[26] If 2/3 (66%) were showed a statistically significant effect, this would have been classified as 'favours intervention group' as the proportion was greater than 50%. However, if only 1/3 (33%) were statistically significant, this would have been classified as 'No difference).

*Intervention Effectiveness:* We used the measures of effectiveness (and statistical significance) comparing selected outcome measure(s) in intervention and control conditions provided by authors to assess intervention effectiveness. We classified intervention effectiveness into one of three categories:

- Results favour intervention; i.e. results indicate that any statistically significant changes in relevant outcomes associated with the intervention in a direction that supports public health are greater in the intervention vs control condition.
- No difference; i.e. results indicate that there are no statistically significant differences in any changes in relevant outcomes between intervention vs control condition.
- Results favour control; i.e. results indicate that any statistically significant changes in relevant outcomes associated with the intervention in a direction that supports public health are greater in control vs intervention condition

*Intervention Equity:* We extracted data on differential effects by a measure of SEP from the primary studies. Where multiple measures of SEP were provided we selected only one. We prioritised measures in the following order (1) income; (2) occupation; (3) education and prioritised the level at which measures were assessed in the following order: (1) individual; (2) household; (3) area-based. We then classified differential effects by SEP into one of three categories:

- likely to reduce inequalities: the intervention led to statistically significant improvement in outcomes in people of lower SEP more than people of higher SEP
- no differential impact by SEP: there is no statistical significant difference in effects (or non-effects) by SEP
- likely to widen inequalities: the intervention improved outcomes in people of higher SEP more than people of lower SEP

##### *Quality assessment*

Quality assessment was conducted in the source reviews and readers are directed there for further information. As such, we did not repeat quality assessment here.

##### Data analysis

Statistical analysis such as a meta-analysis was not possible due to the heterogeneity of included studies. We narratively summarised the included studies, describing patterns of association between DePtH framework category and intervention overall and differential effectiveness categories. We used harvest plots[27] to graphically represent the data and generated separate harvest plots for overall and differential effectiveness by SEP.

#### Results

##### *Search results*

A PRISMA diagram summarising searching, screening and selection is provided in figure 1.10. From the 32,306 records identified in step 1a, 25 included a term based on equity in the title. From these, we purposively selected three systematic reviews[23-25] that included 87 primary studies (McGill (n=37); Olstad (n=36); Beauchamp (n=14)). We removed 9 duplicates, assessed 78 full text articles and included 31 interventions.

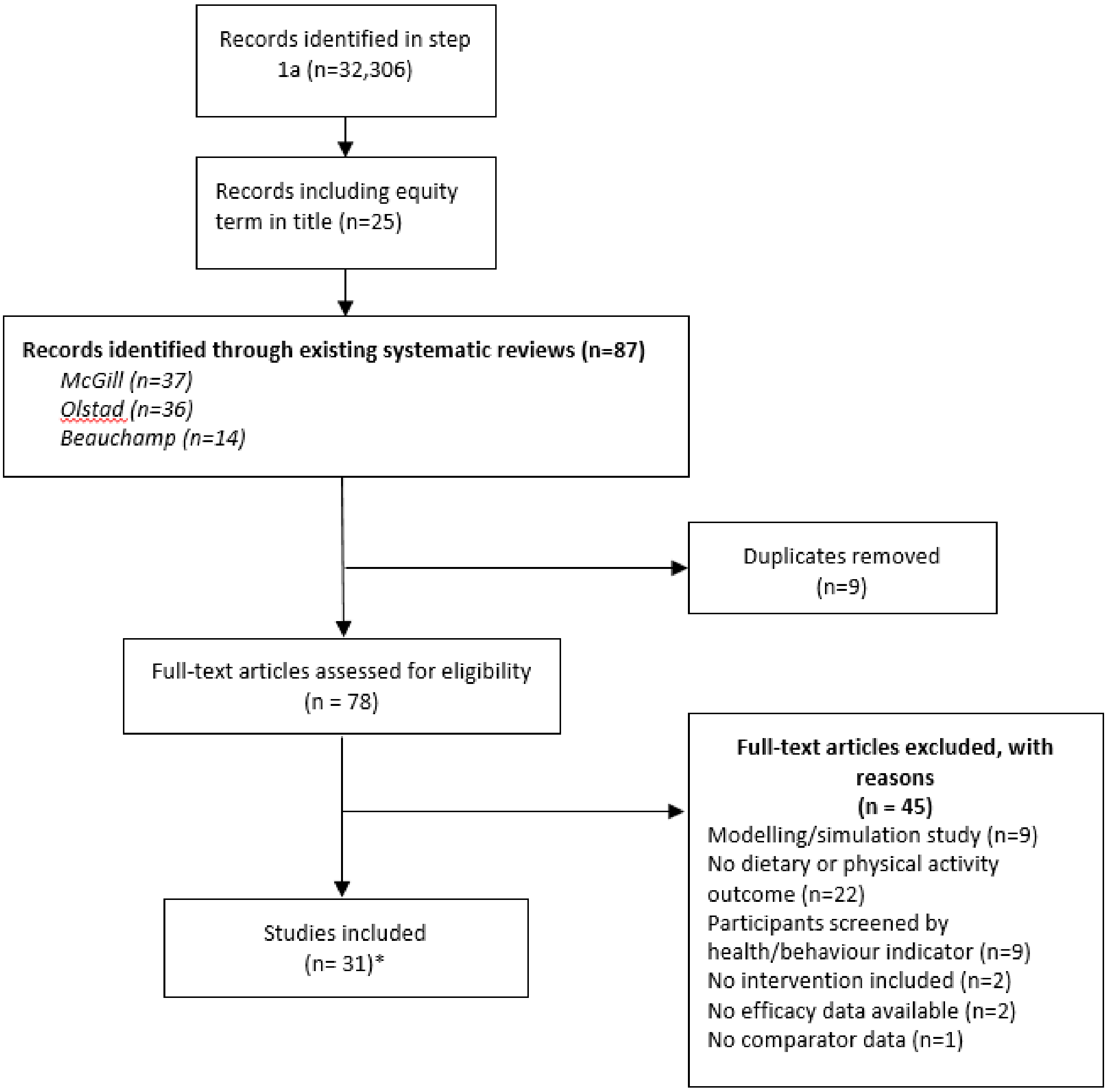


**Figure 1.10** – *PRISMA diagram for step 3*
* 31 interventions described in 33 articles.

***Overview of included studies***

A summary of included interventions is provided in supplementary material 3. Of the 31 interventions, it was not possible to code five interventions using the DePtH framework due to insufficient information on intended intervention recipients.[28-32] Herein, we refer to the 26 interventions included in the analysis.

Included interventions were conducted in the USA (n=11), UK (n=5), Norway (n=2), Netherlands (n=2) and one each in Ireland, Belgium, Canada, France, Australia and Denmark. Study designs included repeat cross sectional (n=10), cluster RCTs (n=7), RCTs (n=5), longitudinal follow up studies (n=3) and 1 quasi-RCT. Of the 26 studies, the majority were conducted in children (n=16) with 9 conducted in adults and 1 in older adults. Consequently 13 study settings were schools, with 9 in the community and one each is churches, healthcare settings, workplaces and girl-scout troops. The studies mainly targeted diet (n=18), with only 4 targeting PA and 4 targeting both dietary and PA outcomes. Supplementary material 3 provides details on intervention characteristics.

***DePtH framework application***

Of the 26 studies, we identified 163 intervention components (Median = 4.5 per intervention; Range = 1-24 per intervention). Of the 163 identified components, there was sufficient information to code all three constructs (exposure, mechanism of action engagement) for 115 intervention components (71%). It was not possible to categorise exposure in 32 components, mechanism of action in 25 and engagement in 20. Insufficient information to categorise framework constructs was more common in studies with a high number of intervention components (>10) where descriptions were, perhaps necessarily, more brief. However, even some studies reporting a small number of intervention components did not provide sufficient information to classify all framework constructs.

*Inter-rater reliability*

As in step 2b, we firstly assessed inter-rater reliability of identified intervention components. We independently extracted intervention components in duplicate in two phases with 50% of studies in each phase. In the first phase, we used the guidance in supplementary material 2. There was agreement on 49% of intervention components, marginal agreement on 36% of components and disagreement on 15% of components (Table 1.7). Following discussion, we reached 100% agreement by applying the additional rule that intervention components should be extracted to the greatest degree of granularity. In the second phase, there was agreement on 75%, marginal agreement on 21% and disagreement on 4% of intervention components. Again, following discussion, we resolved 100% of discrepancies.

**Table 1.7** – *Reviewer agreement on intervention components before discussion*

| | **Agree[a]** | **Marginal agreement[a]** | **Disagree[a]** |
|---|---|---|---|
| ***Step 2**** | | | |
| Inter-rater reliability survey | *63%* | *12%* | *25%* |
| ***Step 3*** | | | |
| Phase 1 | 49% | 36% | 15% |
| Phase 2 | 75% | 21% | 4% |
| Overall | 61% | 29% | 10% |

[a] *Agree* – Two raters description or justification of the intervention was clearly referring to the same intervention component; *Marginal agreement* - Both raters have identified similar intervention components however have categorised them so that they cannot be matched; *Disagree* - These are cases where one rater has identified intervention component that has not been identified by the second rater.
* results from step 2b included for comparison purposes.

Inter-rater reliability statistics for assessing the framework constructs are presented in table 1.8. We coded the framework constructs for 163 intervention component-recipient combinations. Analysis 1 assesses agreement between two raters for all framework constructs coded by reviewers. This shows lower than acceptable agreement. We met to discuss the reasons for discrepancies. In most cases one reviewer coded many constructs as '*insufficient information to code'* based explicitly on the text available whilst the second reviewers did attempt coding using their wider knowledge implied by text available. Analysis 2 removed the framework constructs where there was insufficient information to code, improving the agreement to an acceptable level for the three framework constructs. Agreement was still poor for identifying actors within interventions. We developed additional rules to address disagreements, which included using only information available in the text (Supplementary material 2). Once these rules were applied and further discussion, we resolved all disagreements to reach 100% agreement.

**Table 1.8** – *Inter-rater reliability statistics*

| | Analysis 1 | | | Analysis 2 * | | |
|---|---|---|---|---|---|---|
| | **n** | **Kappa value** | **Interpretation** | **n** | **Kappa value** | **Interpretation** |
| Exposure | 128 | 0.42 | Moderate | 95 | 0.65 | Substantial |
| Mechanism of action | 129 | 0.39 | Fair | 85 | 0.69 | Substantial |
| Engagement | 129 | 0.25 | Fair | 83 | 0.57 | Moderate |
| Macro-environmental actors | 128 | 0.14 | None to slight | 127 | 0.15 | None to slight |
| Micro-environmental actors | 128 | 0.16 | None to slight | 124 | 0.08 | None to slight |
| Informal gatekeepers | 128 | 0.35 | Fair | 125 | 0.40 | Fair |
| Secondary recipients | 128 | 0.01 | None to slight | 110 | 0.02 | None to slight |

* Removal of observations where one rater coded '*Insufficient information to code'*

Learning from this stage suggests that a staged approach is key to extracting data with a high degree of inter-rater reliability. Identifying disagreements after each stage and developing additional data extraction rules to guide subsequent data extraction enhances the efficiency of data extraction and reliability. The rules developed for this project are detailed in supplementary material 2, but we do not intend for these to be definitive. Other researchers may wish to adapt them in line with their aims, the research questions and research team expertise.

***Association with effectiveness and equity***

Figures 1.11 and 1.12 are Harvest Plots that visually display the distribution of intervention components according to their framework classification and overall effectiveness (figure 1.11) and differential effectiveness by SEP (figure 1.12). The structure of the Harvest Plot matrix matches the framework classifications and each cell is a unique framework classification. The three columns within each cell represents the three overall or differential effectiveness classifications. Within the matrix, the height and number above the bar represents the number of intervention component-mechanism combinations that fall within that framework classification and overall or differential effectiveness classification. The black shading represents interventions with dietary outcomes and grey shading those with PA outcomes.

The median number of framework classifications present in a single intervention was 2 (range 1-5 per intervention), and across the interventions we found different distribution patterns across the framework classifications (figure 1.13). Panel A displays framework classifications within the Texas School Nutrition Policy[33-35] demonstrating no variation in framework classification across five intervention component-recipient combinations; all five fall within the passive exposure-physical environmental-passive engagement classification. Panel B displays an intervention[36] with a little more variation with 17 codable component-recipient combinations (out of 24) distributed across three framework classifications. However, all of the 17 components fall within the same row on the framework, showing variation in the mechanisms of action, but not in recipients exposure or engagement. Panel C displays an intervention[37] where all five codable components were in more different framework classifications. Supplementary Appendix 1.6 contains similar diagrams for all included interventions. At this stage it is not possible to determine whether clustering of framework classifications is associated with intervention overall or differential effectiveness.

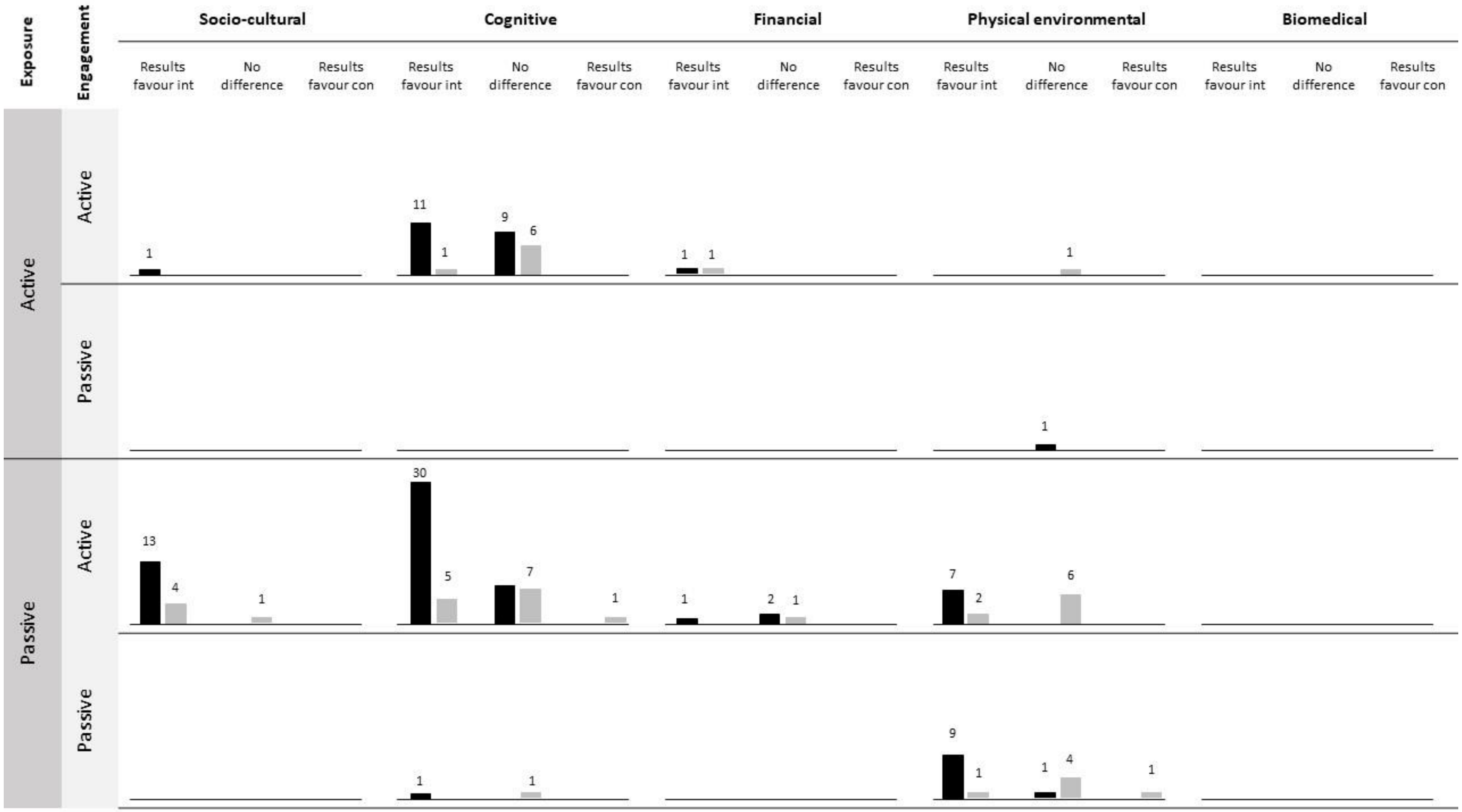


**Figure 1.11** – *Harvest plot illustrating association between DePtH classification and intervention effectiveness.*

Black bars = dietary outcome; Grey bars = PA outcomes. Bar height and numbers = number of component-recipient combinations representing each classification.

Int = Intervention group; Con = Control group.

DePTh classification is at intervention component-recipient level and effectiveness reported at intervention level.

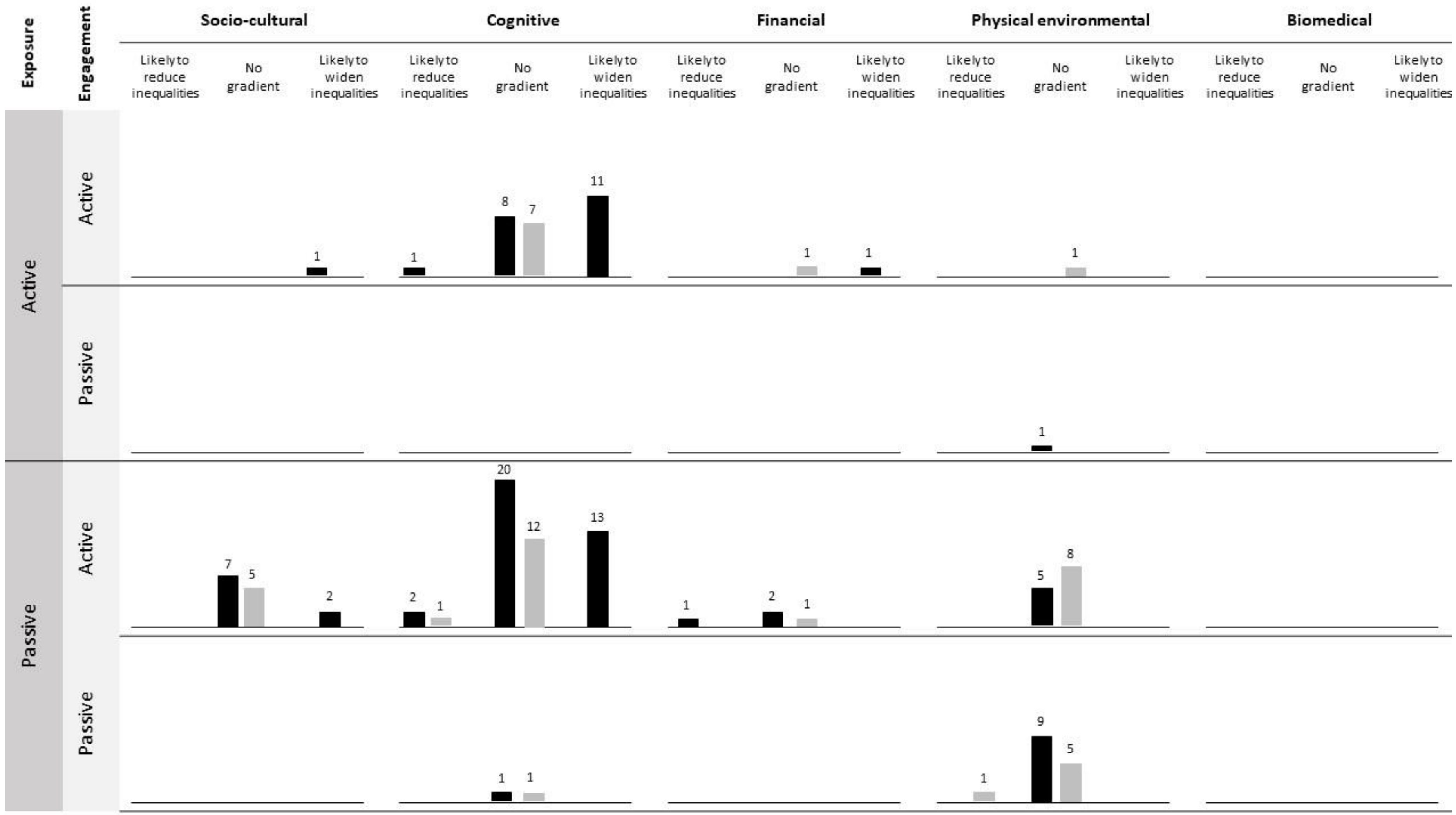


**Figure 1.12** – *Harvest plot illustrating association between DePtH classification and intervention equity.*

Black bars = dietary outcome; Grey bars = PA outcomes. Bar height and numbers = number of component-recipient combinations representing each classification. DePTh classification is at intervention component-recipient level and equity reported at intervention level.

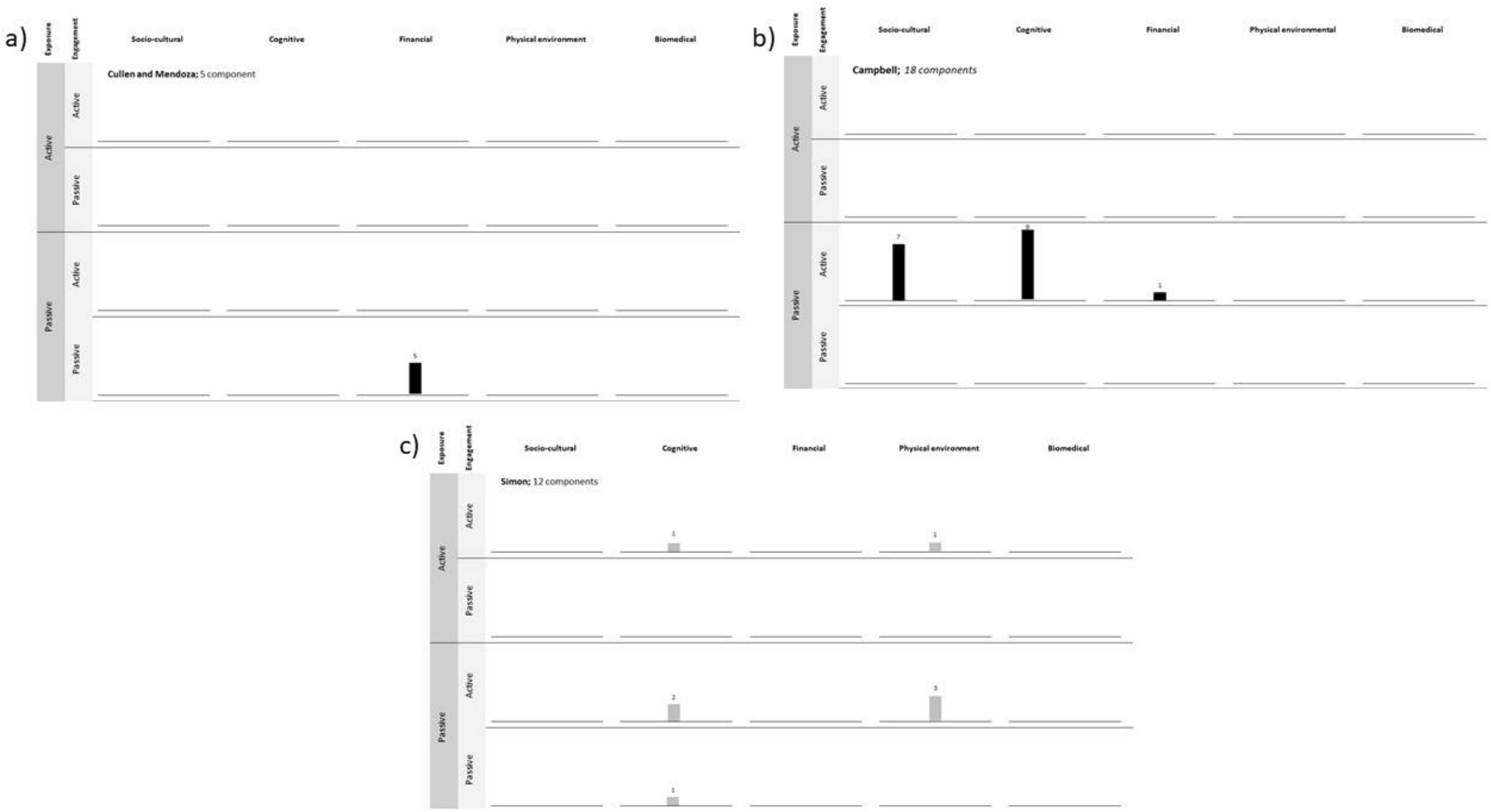


**Figure 1.13** – *Harvest plots to illustrate the differences in distribution of intervention component-recipient combinations within multi-component interventions.* Panel a) intervention with components concentrated within one framework classification. Panel b) intervention with component distributed across multiple mechanisms of action but maintain the same exposure and engagement. Panel c) intervention with components distributed across all framework constructs.

**Supplementary Appendix 1.1** – *Systematic review eligibility criteria*

| | Include | Exclude |
|---|---|---|
| Study design | Systematic reviews, defined where there is a specific inclusion criteria and search strategy (available to readers within the article, eg, supplementary material, in a registered protocol or appendices). | Systematic review protocols<br><br>Reviews of reviews<br><br>Primary studies of interventions, for example RCTs.<br><br>Systematic review where search strategy is available upon request to the authors<br><br>Review that does not have replicable search strategy or defined inclusion criteria. |
| Population | Review includes at least one intervention accessible to whole populations or population group defined by a non-health indicator;<br>• Place of residence<br>• Race, ethnicity, culture or language<br>• Occupation<br>• Gender or sex<br>• Religion<br>• Education<br>• Socioeconomic status<br>• Social capital<br>• Features of relationships (eg, excluded from school)<br>• Time dependent relationships (person at a temporary disadvantage)<br>• Personal characteristics associated with discrimination (age, gender reassignment, marriage and civil partnership, pregnancy or maternity, sexual orientation) | Reviews of interventions for individuals with the presence or absence of a health indicator, eg,<br>• weight status<br>• confirmed disease risk<br>• confirmed disease, eg, cancer diagnosis, stroke<br>• health behaviour, eg, PA levels or dietary pattern<br><br>Studies in disabled populations |
| Outcomes | Review includes a dietary or PA behavioural outcome. These include:<br>• food selection, purchasing or consumption<br>• Breastfeeding<br>• Measures of total PA, domain specific PA or activity specific PA.<br><br>OR | Review that measure the intervention effect on:<br>• sedentary behaviour<br>• Physical fitness outcomes ,eg, peak oxygen uptake, $VO_2$ max, hand grip strength.<br>• Health outcomes, disease incidence or mortality rates. |

| | | |
|---|---|---|
| | Review includes weight-related outcomes for example, body mass index, weight, waist circumference, body fat or waist-to-hip ratio. | |
| Publication type | Published in a peer reviewed journal | Grey literature<br><br>Conference abstracts |
| Date | Published after 2010 | Published before 2010 |

| **Supplementary Appendix 1.2** – *Medline search strategy* | |
|---|---|
| 1 | Meta-Analysis as Topic/ |
| 2 | meta analy$.tw. |
| 3 | metaanaly$.tw. |
| 4 | Meta-Analysis/ |
| 5 | (systematic adj (review$1 or overview$1)).tw. |
| 6 | exp Review Literature as Topic/ |
| 7 | or/1-6 |
| 8 | cochrane.ab. |
| 9 | embase.ab. |
| 10 | (psychlit or psyclit).ab. |
| 11 | (psychinfo or psycinfo).ab. |
| 12 | (cinahl or cinhal).ab. |
| 13 | science citation index.ab. |
| 14 | bids.ab. |
| 15 | cancerlit.ab. |
| 16 | or/8-15 |
| 17 | reference list$.ab. |
| 18 | bibliograph$.ab. |
| 19 | hand-search$.ab. |
| 20 | relevant journals.ab. |
| 21 | manual search$.ab. |
| 22 | or/17-21 |
| 23 | selection criteria.ab. |
| 24 | data extraction.ab. |
| 25 | 23 or 24 |
| 26 | Review/ |
| 27 | 25 and 26 |
| 28 | Comment/ |
| 29 | Letter/ |
| 30 | Editorial/ |
| 31 | animal/ |
| 32 | human/ |
| 33 | 31 not (31 and 32) |
| 34 | or/28-30,33 |
| 35 | 7 or 16 or 22 or 27 |
| 36 | 35 not 34 |
| 37 | Diet, healthy/ |
| 38 | (Diet* adj (interven* or promot* or advice* or health*)).ti,ab. |
| 39 | ((energy or calor* or food) adj (restric* or limit or minim* or low* or reduc*)).ti,ab. |
| 40 | fruit/ |
| 41 | Vegetables/ |

| | |
|---|---|
| 42 | "fruit and vegetable juices"/ |
| 43 | (Fruit* or vegetabl*).ti,ab. |
| 44 | Dietary fiber/ |
| 45 | ((dietary adj (fibre)) or (dietary adj (fiber))).ti,ab. |
| 46 | (Fish oil* or oil* fish or fatty acid or omega?3).ti,ab. |
| 47 | Red meat/ or Diet, atherogenic/ or exp vegetarians/ or diet, vegetarian/ or diet, vegan/ |
| 48 | ((Process* or red) adj (meat* or protein)).ti,ab. |
| 49 | (vegetarian* or vegan*).ti,ab. |
| 50 | Breast feeding/ |
| 51 | ((nursing adj (baby or infant or mother)) or "infant nutri*" or lactating or lactation).ti,ab. |
| 52 | ((Breast or human) adj (milk or feed* or fed)).ti,ab. |
| 53 | (fat adj (low* or reduc* or modif*)).ti,ab. |
| 54 | Exp sodium, dietary/ or sodium chloride/ or diet, sodium-restricted/ |
| 55 | ((Sodium or sodium chloride or salt or dietary sodium) adj (restrict* or curb* or limit* or minim* or low* or reduc* or diet* or free)).ti,ab. |
| 56 | Exp Carbonated beverages/ or energy drinks/ or exp dietary sugars/ or beverages/ or exp agents, sweetening/ |
| 57 | ((sugar or sugar?sweetened or sugarsweetened or sugary or carbonated or fizzy or energy or soft or sport*) adj (drink* or beverage* or soda)).ti,ab. |
| 58 | ((Soda or beverage* or drink*) adj (consumption or intake or sales)).ti,ab. |
| 59 | Exp exercise/ or exp physical fitness/ or muscle strength/ or Walking/ or bicycling/ |
| 60 | (Physical or physically) adj (active or activity).ti,ab. |
| 61 | (MVPA).ti,ab. |
| 62 | ((moderate or vigorous or very vigorous or high) adj (exerci*e or intensity)).ti,ab. |
| 63 | (cycling or cyclist or cycle hire or bicycl* or bike or biking).ti,ab. |
| 64 | (walk or walking or walker or steps).ti,ab. |
| 65 | Sedentary behaviour/ or screen time/ |
| 66 | (sedentary* or sitting or sit or physical inactivity or screen?time or screen?based or television or tv).ti,ab. |
| 67 | or/37-66 |
| 68 | Evaluation study/ |
| 69 | exp Randomized controlled trial/ or evaluation study/ |
| 70 | Randomi*ed control* trial.ti,ab. |
| 71 | Observational study/ |
| 72 | (pilot or feasib* adj (experim* or stud* or trial* or interven* or program* or initiat*)).ti,ab. |
| 73 | (Evaluat* or intervention).ti,ab. |
| 74 | ((Effectiv* or effic*) adj (interven* or stud* or trial* or program* or policy)).ti,ab. |
| 75 | (Natural adj (experim* or evaluat* or stud*)).ti,ab. |
| 76 | (prospective adj (study or studies)).ti,ab. |

| | |
|---|---|
| 77 | (program* evaluation or evaluation research).ti,ab. |
| 78 | (controlled adj (trial or trials or study or studies or experiment*)).ti,ab. |
| 79 | or/68-78 |
| 80 | 36 and 67 and 79 |
| 81 | limit 80 to year="2010-current" |

#### Supplementary Appendix 1.3 – Systematic reviews selected for identifying interventions in step 1


Barberio, Amanda M., et al. "Population-level interventions in government jurisdictions for dietary sodium reduction: a Cochrane Review." *International journal of epidemiology* 46.5 (2017): 1551-1405.

Bleich, Sara N., et al. "Interventions to prevent global childhood overweight and obesity: a systematic review." *The Lancet Diabetes & Endocrinology* 6.4 (2018): 332-346.

Brown, Vicki, et al. "A systematic review of economic analyses of active transport interventions that include PA benefits." *Transport Policy* 45 (2016): 190-208.

Calancie, Larissa, et al. "Nutrition-related policy and environmental strategies to prevent obesity in rural communities: a systematic review of the literature, 2002–2013." (2015).

Ling, Jiying, Lorraine B. Robbins, and Fujun Wen. "Interventions to prevent and manage overweight or obesity in preschool children: A systematic review." *International journal of nursing studies* 53 (2016): 270-289.

McKinnon, Robin A., et al. "Obesity-related policy/environmental interventions: a systematic review of economic analyses." *American journal of preventive medicine* 50.4 (2016): 543-549.

Perez-Ferrer, Carolina, et al. "The food environment in Latin America: A systematic review with a focus on environments relevant to obesity and related chronic diseases." *Public health nutrition* 22.18 (2019): 3447-3464.

Richardson, Andrea S., et al. "Obesity prevention interventions and implications for energy balance in the United States and Mexico: a systematic review of the evidence and meta-analysis." *Obesity* 27.9 (2019): 1390-1403.

Scheepers, C. E., et al. "Shifting from car to active transport: a systematic review of the effectiveness of interventions." *Transportation research part A: policy and practice* 70 (2014): 264-280.

#### Supplementary Appendix 1.4 – Included interventions and papers in step 1

* indicates index paper from which other papers within the intervention collection were identified.

##### 10,000 Steps Ghent

De Cocker, Katrien A., et al. "Effects of "10,000 steps Ghent": a whole-community intervention." *American journal of preventive medicine* 33.6 (2007): 455-463.

De Cocker, Katrien A., et al. "Four-year follow-up of the community intervention '10 000 steps Ghent'." *Health Education Research* 26.2 (2011): 372-380.

De Cocker, Katrien, et al. "Moderators and mediators of pedometer use and step count increase in the" 10,000 Steps Ghent" intervention." *International Journal of Behavioral Nutrition and PA* 6.1 (2009): 1-7.

De Cocker, Katrien A., et al. "The effect of a pedometer-based PA intervention on sitting time." *Preventive Medicine* 47.2 (2008): 179-181.

* De Smedt, Delphine, et al. "A cost-effectiveness study of the community-based intervention '10 000 Steps Ghent'." *Public health nutrition* 15.3 (2012): 442-451.

##### Apache Healthy Stores

* Curran, Sarah, et al. "Process evaluation of a store-based environmental obesity intervention on two American Indian Reservations." *Health Education Research* 20.6 (2005): 719-729.

Ethelbah, B., et al. "The Apache Healthy Stores program: Results of the formative research." *FASEB JOURNAL* 17.4 (2003).

Gittelsohn, Joel, and Megan Rowan. "Preventing diabetes and obesity in American Indian communities: the potential of environmental interventions." *The American journal of clinical nutrition* 93.5 (2011): 1179S-1183S.

Vastine, Amy, et al. "Formative research and stakeholder participation in intervention development." *American journal of health behavior* 29.1 (2005): 57-69.

##### Apple Project

* McAuley, Kirsten A., et al. "Economic evaluation of a community-based obesity prevention program in children: the APPLE project." *Obesity* 18.1 (2010): 131-136.

Taylor, Rachael W., et al. "APPLE Project: 2-y findings of a community-based obesity prevention program in primary school–age children." *The American journal of clinical nutrition* 86.3 (2007): 735-742.

Taylor, Rachael W., et al. "Reducing weight gain in children through enhancing PA and nutrition: the APPLE project." *International Journal of Pediatric Obesity* 1.3 (2006): 146-152.

Taylor, Rachael W., et al. "Two-year follow-up of an obesity prevention initiative in children: the APPLE project." *The American Journal of Clinical Nutrition* 88.5 (2008): 1371-1377.

Williden, Micalla, et al. "The APPLE project: An investigation of the barriers and promoters of healthy eating and PA in New Zealand children aged 5-12 years." *Health Education Journal* 65.2 (2006): 135-148.

##### Arkansas Act 1220

Craig, Rebekah L., et al. "Public health professionals as policy entrepreneurs: Arkansas's childhood obesity policy experience." *American Journal of Public Health* 100.11 (2010): 2047-2052.

Krukowski, Rebecca A., et al. "Development and evaluation of the school cafeteria nutrition assessment measures." *Journal of School Health* 81.8 (2011): 431-436.

Krukowski, Rebecca A., et al. "No change in weight-based teasing when school-based obesity policies are implemented." *Archives of pediatrics & adolescent medicine* 162.10 (2008): 936-942.

Krukowski, Rebecca A., et al. "Overweight children, weight-based teasing and academic performance." *International Journal of Pediatric Obesity* 4.4 (2009): 274-280.

Phillips, Martha M., et al. "Changes in school environments with implementation of Arkansas Act 1220 of 2003." *Obesity* 18.S1 (2010): S54-S61.

Phillips, Martha M., Kevin Ryan, and James M. Raczynski. "Public policy versus individual rights in childhood obesity interventions: perspectives from the Arkansas experience with Act 1220 of 2003." (2011).

* Phillips, Martha M., et al. "The evaluation of Arkansas Act 1220 of 2003 to reduce childhood obesity: conceptualization, design, and special challenges." *American journal of community psychology* 51.1 (2013): 289-298.

Raczynski, James M., et al. "Arkansas Act 1220 of 2003 to reduce childhood obesity: its implementation and impact on child and adolescent body mass index." *Journal of Public Health Policy* 30.1 (2009): S124-S140.

#### Be Active

* Frew, Emma J., et al. "Cost-effectiveness of a community-based PA programme for adults (Be Active) in the UK: an economic analysis within a natural experiment." *British Journal of Sports Medicine* 48.3 (2014): 207-212.

#### Be Fit

Thorndike, Anne N., et al. "Abstract P145: Using an Activity Monitor to Motivate PA among Healthy Employees: A Randomized Controlled Trial." (2012): AP145-AP145.

* Thorndike, Anne N., et al. "Activity monitor intervention to promote PA of physicians-in-training: randomized controlled trial." *PloS one* 9.6 (2014): e100251.

#### Being Active, Eating Well

Bolton, Kristy A., et al. "Expanding a successful community-based obesity prevention approach into new communities: challenges and achievements." *Obesity Research & Clinical Practice* 10.2 (2016): 197-206.

Bolton, Kristy A., et al. "The association between self-reported diet quality and health-related quality of life in rural and urban Australian adolescents." *Australian journal of rural health* 24.5 (2016): 317-325.

* Bolton, K. A., et al. "The outcomes of health-promoting communities: being active eating well initiative—a community-based obesity prevention intervention in Victoria, Australia." *International Journal of Obesity* 41.7 (2017): 1080-1090.

de Silva-Sanigorski, Andrea M., et al. "Scaling up community-based obesity prevention in Australia: background and evaluation design of the health promoting communities: being active eating well initiative." *BMC public health* 10.1 (2010): 1-7.

Kelaher, Margaret, et al. "Final evaluation report for “Go for your life” Health Promoting Communities: Being active and eating well initiative (HPC: BAEW) Wathaurong Aboriginal Co-operative." *University of Melbourne, Melbourne* (2011).

#### Bike Now

O'Fallon, Carolyn. "Bike Now: encouraging cycle commuting in New Zealand." *New Zealand Transport Agency Research Report* 414 (2010).

* O'Fallon, Carolyn. "Bike Now: Exploring methods of building sustained participation in cycle commuting in New Zealand." *Road & Transport Research: A Journal of Australian and New Zealand Research and Practice* 19.2 (2010): 77-89.

#### Bike, Walk, Wheel

Sayers, Stephen P., et al. "Bike, walk, and wheel: a way of life in Columbia, Missouri, revisited." *American journal of preventive medicine* 43.5 (2012): S379-S383.

* Thomas, Ian M., et al. "Bike, walk, and wheel: a way of life in Columbia, Missouri." *American journal of preventive medicine* 37.6 (2009): S322-S328.

#### BIXI

Bélanger-Gravel, Ariane, et al. "Association of implementation of a public bicycle share program with intention and self-efficacy: The moderating role of socioeconomic status." *Journal of health psychology* 21.6 (2016): 944-953.

Bélanger-Gravel, Ariane, et al. "Implementing a public bicycle share program: impact on perceptions and support for public policies for active transportation." *Journal of PA and health* 12.4 (2015): 477-482.

Bernatchez, Annie C., et al. "Knowing about a public bicycle share program in Montreal, Canada: are diffusion of innovation and proximity enough for equitable awareness?." *Journal of Transport & Health* 2.3 (2015): 360-368.

Fuller, Daniel, et al. "Impact evaluation of a public bicycle share program on cycling: a case example of BIXI in Montreal, Quebec." *American journal of public health* 103.3 (2013): e85-e92.

Fuller, Daniel, Lise Gauvin, and Yan Kestens. "Individual-and area-level disparities in access to the road network, subway system and a public bicycle share program on the Island of Montreal, Canada." *Annals of behavioral medicine* 45.suppl_1 (2013): S95-S100.

Fuller, Daniel. *Potential of built environment interventions involving deployment of public bicycles to increase utilitarian cycling: The case of BIXI© in Montreal, Quebec*. Universite de Montreal (Canada), 2012.

Fuller, D., et al. "Sociodemographic correlates of public bicycle share program use: An intercept survey of users in Montreal, Canada." *Journal of Science and Medicine in Sport* 15 (2012): S71.

Fuller, Daniel, et al. "The impact of implementing a public bicycle share program on the likelihood of collisions and near misses in Montreal, Canada." *Preventive medicine* 57.6 (2013): 920-924.

* Fuller, Daniel, et al. "The potential modal shift and health benefits of implementing a public bicycle share program in Montreal, Canada." *International journal of behavioral nutrition and PA* 10.1 (2013): 1-6.

Fuller, Daniel, et al. "Use of a new public bicycle share program in Montreal, Canada." *American journal of preventive medicine* 41.1 (2011): 80-83.

#### Boston’s Downtown Crossing

* Weisbrod, Glen. "Business and travel impacts of Boston's Downtown Crossing automobile-restricted zone." *Transportation Research Record* 882 (1982): 25-32.

Weisbrod, Glen, et al. *Downtown Crossing: Auto Restricted Zone in Boston*. No. UMTA-MA-06-0049-82-3. United States. Department of Transportation. Urban Mass Transportation Administration, 1982.

Weisbrod, Glen, and Henry O. Pollakowski. "Effects of downtown improvement projects on retail activity." *Journal of the American Planning association* 50.2 (1984): 148-161.

#### Burbidge-2009

* Burbidge, Shaunna K., and Konstadinos G. Goulias. "Evaluating the impact of neighborhood trail development on active travel behavior and overall PA of suburban residents." *Transportation research record* 2135.1 (2009): 78-86.

#### Cashing Out

Medina, Hazael. "An Analysis of Local Government Implementation of California's Parking Cash-Out Law." (2020).

Shoup, Donald C. "An opportunity to reduce minimum parking requirements." *Journal of the American planning association* 61.1 (1995): 14-28.

Shoup, D. C. “Cashing out employer-paid parking: An opportunity to reduce minimum parking requirements. Working paper.” No. UCTC No. 204. 1994.

Shoup, Donald C. "Cashing out employer-paid parking: a precedent for congestion pricing?." (1993).

Shoup, Donald C., and Richard W. Willson. "Commuting, congestion and pollution: the employer-paid parking connection." (1992).

Shoup, Donald C. "Congress okays cash out." *ACCESS Magazine* 1.13 (1998): 2-8.

Shoup, Donald C., and Mary Jane Breinholt. "Employer-paid parking: a nationwide survey of employers’ parking subsidy policies." *The full costs and benefits of transportation*. Springer, Berlin, Heidelberg, 1997. 371-385.

Shoup, Donald C., and Richard W. Willson. "Employer-paid parking: the problem and proposed solutions." (1992).

* Shoup, Donald C. "Evaluating the effects of cashing out employer-paid parking: eight case studies." *Transport Policy* 4.4 (1997): 201-216.

Shoup, Donald C. "Parking Cash Out. American Planning Association." *Planning Advisory Service, Chicago, IL, USA* (2005).

#### CATCH

* Brown, Henry Shelton, et al. "The cost-effectiveness of a school-based overweight program." *International Journal of Behavioral Nutrition and PA* 4.1 (2007): 1-12.

Coleman, Karen J., et al. "Prevention of the epidemic increase in child risk of overweight in low-income schools: the El Paso coordinated approach to child health." *Archives of pediatrics & adolescent medicine* 159.3 (2005): 217-224.

Heath, Edward M., and Karen J. Coleman. "Adoption and institutionalization of the child and adolescent trial for cardiovascular health (CATCH) in El Paso, Texas." *Health Promotion Practice* 4.2 (2003): 157-164.

Heath, Edward M., and Karen J. Coleman. "Evaluation of the institutionalization of the coordinated approach to child health (CATCH) in a US/Mexico border community." *Health education & behavior* 29.4 (2002): 444-460.

#### Charlotte Area Transit System

MacDonald, John M., et al. "The effect of light rail transit on body mass index and PA." *American journal of preventive medicine* 39.2 (2010): 105-112.

* Stokes, Robert J., John MacDonald, and Greg Ridgeway. "Estimating the effects of light rail transit on health care costs." *Health & Place* 14.1 (2008): 45-58.

#### Cherokee Choices

* Bachar, Jeffrey J., et al. "Cherokee Choices: a diabetes prevention program for American Indians." *Preventing Chronic Disease* 3.3 (2006).

Bachar, Jeff. "Cherokee Choices Diabetes Prevention Program: The Eastern Band of Cherokee Indians." *Diabetes and Health Disparities* (2009): 275.

Bachar, Jeff. "Sidebar: Cherokee Choices: A Diabetes Prevention Program in Cherokee, North Carolina." *North Carolina Medical Journal* 72.5 (2011): 394-395.

Bachar, Jeffrey J., et al. "VOCES COMUNITARIAS Cherokee Choices (Opciones cherokee): Programa de prevención de la diabetes para indígenas americanos." *VOCES* 3.3 (2006): 05_0221_es.

Bell, Ronny A. "Barriers to diabetes prevention and control among American Indians." *North Carolina medical journal* 72.5 (2011): 393-396.

#### Ciclovia

Meisel, Jose D., et al. "Network analysis of Bogota's Ciclovia Recreativa, a self-organized multisectorial community program to promote PA in a middle-income country." *American journal of health promotion* 28.5 (2014): e127-e136.

Montero, Sergio. "Worlding Bogotá's ciclovía: from urban experiment to international "best practice"." *Latin American Perspectives* 44.2 (2017): 111-131.

* Montes, Felipe, et al. "Do health benefits outweigh the costs of mass recreational programs? An economic analysis of four Ciclovía programs." *Journal of Urban Health* 89.1 (2012): 153-170.

Parra, Diana C., et al. "Geographic distribution of the Ciclovia and Recreovia programs by neighborhood SES in Bogotá: how unequal is the geographic access assessed via distance-based measures?." *Journal of Urban Health* 98.1 (2021): 101-110.

Sarmiento, Olga L., et al. "Promotion of recreational walking: Case study of the Ciclovía-Recreativa of Bogotá." *Walking*. Emerald Publishing Limited, 2017.

Torres, Andrea D. "The Bogota ciclovia-recreativa and cicloruta programs: promising interventions to promote PA, and social capital in the city of Bogota." (2012).

Triana, Camilo A., et al. "Active streets for children: The case of the Bogotá Ciclovía." *PloS one* 14.5 (2019): e0207791.

Sarmiento Dueñas, Olga Lucía, Gloria Niño, and Oscar Alberto Bernal Acevedo. "Promotion of PA among children through a community intervention-the case of the ciclovía of Bogotá."

#### Clean Air Force Campaign

* Alcott, Randi, and Maureen Mageau DeCindis. "Clean Air Force Campaign 1989-1990: programs, attitudes, and commute behavior changes." *Transportation Research Record* 1321 (1991): 34-44.

#### Conrey-2003

* Conrey, Elizabeth J., et al. "Integrated program enhancements increased utilization of farmers' market nutrition program." *The Journal of nutrition* 133.6 (2003): 1841-1844.

Dollahite, Jamie S., et al. "Building community capacity through enhanced collaboration in the farmers market nutrition program." *Agriculture and Human Values* 22.3 (2005): 339-354.

#### CROPP

Schwarte, Liz, et al. "The Central California Regional Obesity Prevention Program: changing nutrition and PA environments in California's heartland." *American journal of public health* 100.11 (2010): 2124-2128.

#### Croteau

* Croteau, Karen A., et al. "Effect of a pedometer-based intervention on daily step counts of community-dwelling older adults." *Research quarterly for exercise and sport* 78.5 (2007): 401-406.

Jones, David B., et al. "Focus groups to explore the perceptions of older adults on a pedometer-based intervention." *Research quarterly for exercise and sport* 80.4 (2009): 710-717.

#### Cycling Demonstration Towns

Cavill, Nick, Andy Cope, and Angela Kennedy. "Valuing increased cycling in the Cycling Demonstration Towns." *Sustrans and Cavill Associates: London, UK* (2009).

* Cope, Andy, et al. "Cycling demonstration towns–an economic evaluation." *Association for European transport and contributors* (2010).

Goodman, Anna, et al. "Effectiveness and equity impacts of town-wide cycling initiatives in England: a longitudinal, controlled natural experimental study." *Social science & medicine* 97 (2013): 228-237.

Goodman, A., et al. "OP22 Effectiveness and Equity Impact of Town-Wide Cycling Investment in England: A Longitudinal, Controlled Natural Experimental Study." *J Epidemiol Community Health* 67.Suppl 1 (2013): A13-A13.

Pooley, Colin, et al. "Shaping the city for walking and cycling: a case study of Lancaster." *Built Environment* 36.4 (2010): 447-460.

Sloman, Lynn, et al. "Summary of outcomes of the Cycling Demonstration Towns and Cycling City and Towns programmes." (2017).

#### EFNEP

* Schuster, Ellen, et al. "Investing in Oregon's Expanded Food and Nutrition Education Program (EFNEP): documenting costs and benefits." *Journal of Nutrition Education and Behavior* 35.4 (2003): 200-206.

#### Elbel

* Elbel, Brian, et al. "Assessment of a government-subsidized supermarket in a high-need area on household food availability and children’s dietary intakes." *Public health nutrition* 18.15 (2015): 2881-2890.

Elbel, Brian, et al. "The introduction of a supermarket via tax-credits in a low-income area: the influence on purchasing and consumption." *American Journal of Health Promotion* 31.1 (2017): 59-66.

Infahsaeng, Tida. *Inner-City Grocery Store Development as Community Economic Development: A Case Study of the New York City Food Retail Expansion to Support Health Program (FRESH)*. Diss. Tufts University, 2014.

Rogus, Stephanie, et al. "Measuring micro-level effects of a new supermarket: do residents within 0.5 mile have improved dietary behaviors?." *Journal of the Academy of Nutrition and Dietetics* 118.6 (2018): 1037-1046.

#### Family Fitness Zones

* Cohen, Deborah A., et al. "Impact and cost-effectiveness of family fitness zones: a natural experiment in urban public parks." *Health & place* 18.1 (2012): 39-45.

#### Fit for Life Boy Scout Badge

Jago, Russell, et al. "Distance to food stores & adolescent male fruit and vegetable consumption: mediation effects." *International Journal of Behavioral Nutrition and PA* 4.1 (2007): 1-10.

* Jago, Russell, et al. "Fit for Life Boy Scout badge: outcome evaluation of a troop and Internet intervention." *Preventive medicine* 42.3 (2006): 181-187.

Latif, Hira, et al. "Effects of goal setting on dietary and PA changes in the Boy Scout badge projects." *Health education & behavior* 38.5 (2011): 521-529.

Lu, A. S., et al. "Five-a-day and fit-for-life badge programs for cancer prevention in Boy Scouts." *Cancer Disparities: Causes, Evidence-based Solutions* (2011): 169-92.

#### Fitter for Walking

Adams, E., M. Goad, and N. Cavill. "Changing the environment to promote transport-related walking: Evaluation of the 'Fitter for Walking'project." *Journal of Science and Medicine in Sport* 15 (2012): S69.

Adams, Emma J., and Lauren B. Sherar. "Community perceptions of the implementation and impact of an intervention to improve the neighbourhood physical environment to promote walking for transport: a qualitative study." *BMC public health* 18.1 (2018): 1-14.

Adams, Emma J., and Nick Cavill. "Engaging communities in changing the environment to promote transport-related walking: Evaluation of route use in the 'Fitter for Walking'project." *Journal of Transport & Health* 2.4 (2015): 580-594.

Adams, Emma, Mary Goad, and Nick Cavill. "Evaluation of Living Streets' Fitter for Walking project." (2012).

Adams, Emma J., Nick Cavill, and Lauren B. Sherar. "Evaluation of the implementation of an intervention to improve the street environment and promote walking for transport in deprived neighbourhoods." *BMC public health* 17.1 (2017): 1-20.

* Sinnett, Danielle, and Jane Powell. "Economic evaluation of Living Streets' fitter for walking project." (2012).

#### Get Up and Do Something

Abraham, A., P. M. Peterson, and D. A. Waterfield. "Efficacy of a statewide social marketing campaign to promote PA." *Medicine & Science in Sports & Exercise* 35.5 (2003): S220.

* Peterson, Michael, Margaret Chandlee, and Avron Abraham. "Cost-effectiveness analysis of a statewide media campaign to promote adolescent PA." *Health Promotion Practice* 9.4 (2008): 426-433.

Peterson, Michael, Avron Abraham, and Allan Waterfield. "Marketing PA: Lessons learned from a statewide media campaign." *Health Promotion Practice* 6.4 (2005): 437-446.

#### Guide to Health

Anderson, Eileen S., et al. "Social-cognitive determinants of PA: the influence of social support, self-efficacy, outcome expectations, and self-regulation among participants in a church-based health promotion study." *Health psychology* 25.4 (2006): 510.

Anderson, Eileen S., et al. "Social cognitive mediators of change in a group randomized nutrition and PA intervention: social support, self-efficacy, outcome expectations and self-regulation in the guide-to-health trial." *Journal of health psychology* 15.1 (2010): 21-32.

Anderson, Eileen S., Richard A. Winett, and Janet R. Wojcik. "Self-regulation, self-efficacy, outcome expectations, and social support: social cognitive theory and nutrition behavior." *Annals of behavioral medicine* 34.3 (2007): 304-312.

* Winett, Richard A., et al. "Guide to health: nutrition and PA outcomes of a group-randomized trial of an Internet-based intervention in churches." *Annals of Behavioral Medicine* 33.3 (2007): 251-261.

#### Harnack

French, Simone A., et al. "Financial incentives and purchase restrictions in a food benefit program affect the types of foods and beverages purchased: results from a randomized trial." *International Journal of Behavioral Nutrition and PA* 14.1 (2017): 1-10.

* Harnack, Lisa, et al. "Effects of subsidies and prohibitions on nutrition in a food benefit program: a randomized clinical trial." *JAMA internal medicine* 176.11 (2016): 1610-1619.

Rydell, Sarah A., et al. "Participant satisfaction with a food benefit program with restrictions and incentives." *Journal of the Academy of Nutrition and Dietetics* 118.2 (2018): 294-300.

Valluri, Sruthi, et al. "Within-and Between-Household Variation in Food Expenditures Among Low-Income Households Using a Novel Simple Annotated Receipt Method." *Frontiers in nutrition* 7 (2020): 582999.

#### Healthy Foods North

Bains, A., et al. "Healthy Foods North improves diet among Inuit and Inuvialuit women of childbearing age in Arctic Canada." *Journal of human nutrition and dietetics* 27 (2014): 175-185.

Gittelsohn, J., et al. "Participatory approaches for a community-based chronic disease prevention program in two Canadian Inuit communities: Development of Healthy Foods North." *International journal of circumpolar health.* Vol. 69. 2-4 Park Square, Milton Park, Abingdon OR14 4RN, Oxon, England: Taylor & Francis Ltd, 2010.

Hopping, B., et al. "Nutrient intake among Inuit in the Canadian arctic: Results from Healthy Foods North." *International journal of circumpolar health.* Vol. 69. 2-4 Park Square, Milton Park, Abingdon OR14 4RN, Oxon, England: Taylor & Francis Ltd, 2010.

Kolahdooz, Fariba, et al. "Impact of the Healthy Foods North nutrition intervention program on Inuit and Inuvialuit food consumption and preparation methods in Canadian Arctic communities." *Nutrition journal* 13.1 (2014): 1-10.

Mead, Erin L., et al. "A community-based, environmental chronic disease prevention intervention to improve healthy eating psychosocial factors and behaviors in indigenous populations in the Canadian Arctic." *Health education & behavior* 40.5 (2013): 592-602.

Mead, E., et al. "Factors influencing diet and the food environment in two Inuit communities in Nunavut: Qualitative formative research results from Healthy Foods North." *International journal of circumpolar health.* Vol. 69. 2-4 Park Square, Milton Park, Abingdon OR14 4RN, Oxon, England: Taylor & Francis Ltd, 2010.

* Mead, Erin, et al. "Impact of the changing food environment on dietary practices of an Inuit population in Arctic Canada." *Journal of Human Nutrition and Dietetics* 23 (2010): 18-26.

Mead, E., et al. "The influence of psychosocial factors on food-related behaviours among Inuit communities in Nunavut: Results from Healthy Foods North." *International journal of circumpolar health.* Vol. 69. 2-4 Park Square, Milton Park, Abingdon OR14 4RN, Oxon, England: Taylor & Francis Ltd, 2010.

Mead, E., et al. "The psychosocial determinants of diet-related behaviours among the Inuvialuit: Results from Healthy Foods North." *International journal of circumpolar health.* Vol. 69. 2-4 Park Square, Milton Park, Abingdon OR14 4RN, Oxon, England: Taylor & Francis Ltd, 2010.

Pakseresht, Mohammadreza, et al. "Improving vitamin A and D intake among Inuit and Inuvialuit in Arctic Canada: evidence from the Healthy Foods North study." *J Epidemiol Community Health* 69.5 (2015): 453-459.

Sharma, Sangita, et al. "Addressing the public health burden caused by the nutrition transition through the Healthy Foods North nutrition and lifestyle intervention programme." *Journal of Human Nutrition and Dietetics* 23 (2010): 120-127.

Sharma, Sangita, et al. "Inadequate diets in an Arctic population undergoing a drastic environmental change." (2010): 559-3.

Sharma, Sangita, et al. "Working with communities and the private sector in the Canadian arctic." *Population health intervention research casebook* (2011): 36.

Ugyuk, M., et al. "Implementing a nutrition intervention program among the Inuit in Nunavut: Store-centred activities of Healthy Foods North." *International journal of circumpolar health.* Vol. 69. 2-4 Park Square, Milton Park, Abingdon OR14 4RN, Oxon, England: Taylor & Francis Ltd, 2010.

#### Healthy Food Hawaii

* Gittelsohn, Joel, et al. "A food store intervention trial improves caregiver psychosocial factors and children's dietary intake in Hawaii." *Obesity* 18.S1 (2010): S84-S90.

Gittelsohn, Joel, et al. "Food preparation and psychosocial factors affect dietary intake in Native Hawaiian communities: baseline results from the Healthy Foods Hawaii intervention." (2006): A173-A173.

Novotny, Rachel, et al. "Development and Implementation of a Food System Intervention to Prevent Childhood Obesity in Rural Hawai ʻi." *Hawaii Medical Journal* 70.7 suppl 1 (2011): 42.

#### Italian EAT project

Briganti, S. "The Italian EAT Project: effectiveness of a multicomponent school-based health promotion study on measures of fatness and behavior in teenagers." *Eating and Weight Disorders* (2014): 444-5.

* Ermetici, Federica, et al. "Association between a school-based intervention and adiposity outcomes in adolescents: The Italian “EAT” project." *Obesity* 24.3 (2016): 687-695.

Malavazos, A. E., et al. "The Italian EAT project: Effectiveness of a multicomponent school-based health promotion study on measures of fatness and behaviour in teenagers.” *Acta paediatrica.* Vol 106. 11 River St, Hoboken 07030-5774, NJ USA: Wiley, 2017.

Romanelli, Massimiliano Marco Corsi, et al. "An Italian Obesity Prevention Intervention Study in Teenagers: the EAT project. Effectiveness of a School-Based Program on measures of fatness and behaviour over 2 school years." (2013): 344-5.

#### Kain-2014

Kain, Juliana, et al. "Evaluación de una intervención en educación alimentaria y actividad física para prevenir obesidad infantil en escuelas públicas de Santiago de Chile." *Archivos Latinoamericanos de Nutrición* 62.1 (2012): 60-67.

* Kain, Juliana, et al. "School-based obesity prevention intervention in Chilean children: effective in controlling, but not reducing obesity." *Journal of obesity* 2014 (2014).

#### Lobanco

de Lira Andrade, Mychelle, et al. "ROTULAGEM DE SALGADINHOS A BASE DE MILHO: ANÁLISE DA ADEQUAÇÃO AS LEGISLAÇÕES VIGENTES LABELING OF MAIZE-BASED SALGADINHANS: ANALYSIS OF FITNESS APPLICABLE LEGISLATION."

de Oliveira Sanches, Jonatas Ferreira. "Avaliação da rotulagem de diferentes marcas de biscoito frente à legislação nacional vigente." *Brazilian Journal of Development* 6.4 (2020): 22450-22468.

* Lobanco, Cássia Maria, et al. "Reliability of food labels from products marketed in the city of Sao Paulo, Southeastern Brazil." *Revista de Saúde Pública* 43 (2009): 499-505.

**Local wellness policy**

* Belansky, Elaine S., et al. "Early effects of the federally mandated local wellness policy on school nutrition environments appear modest in Colorado's rural, low-income elementary schools." *Journal of the American Dietetic Association* 110.11 (2010): 1712-1717.

Belansky, Elaine, Jamie F. Chriqui, and Marlene B. Schwartz. "Local school wellness policies: How are schools implementing the congressional mandate?." (2009).

Belansky, Elaine S., et al. "Local Wellness Policy 5 years later: is it making a difference for students in low-income, rural Colorado elementary schools?." *Preventing Chronic Disease* 10 (2013).

#### Mexico SSB tax

Barquera, Simón, Ismael Campos, and Juan A. Rivera. "M exico attempts to tackle obesity: the process, results, push backs and future challenges." *Obesity reviews* 14 (2013): 69-78.

Barrientos-Gutierrez, Tonatiuh, et al. "Expected population weight and diabetes impact of the 1-peso-per-litre tax to sugar sweetened beverages in Mexico." *PloS one* 12.5 (2017): e0176336.

Barrientos-Gutiérrez, Tonatiuh, et al. "Posicionamiento sobre los impuestos a alimentos no básicos densamente energéticos y bebidas azucaradas." *salud pública de méxico* 60.5 (2018): 586-591.

Basto-Abreu, Ana, et al. "Cost-effectiveness of the sugar-sweetened beverage excise tax in Mexico." *Health Affairs* 38.11 (2019): 1824-1831.

Basto-Abreu, Ana, et al. "Expected changes in obesity after reformulation to reduce added sugars in beverages: a modeling study." *PLoS medicine* 15.10 (2018): e1002664.

Batis, Carolina, et al. "Energy, added sugar, and saturated fat contributions of taxed beverages and foods in Mexico." *salud pública de méxico* 59 (2017): 512-517.

Batis, Carolina, et al. "First-year evaluation of Mexico’s tax on nonessential energy-dense foods: an observational study." *PLoS medicine* 13.7 (2016): e1002057.

Carriedo, Angela, et al. "The political economy of sugar-sweetened beverage taxation in Latin America: lessons from Mexico, Chile and Colombia." *Globalization and health* 17.1 (2021): 1-14.

Colchero, M. Arantxa, Mariana Molina, and Carlos M. Guerrero-López. "After Mexico implemented a tax, purchases of sugar-sweetened beverages decreased and water increased: difference by place of residence, household composition, and income level." *The Journal of nutrition* 147.8 (2017): 1552-1557.

Colchero, M. Arantxa, et al. "Beverage purchases from stores in Mexico under the excise tax on sugar sweetened beverages: observational study." *bmj* 352 (2016).

* Colchero, M. Arantxa, et al. "Beverages sales in Mexico before and after implementation of a sugar sweetened beverage tax." *PloS one* 11.9 (2016): e0163463.

Colchero, M. Arantxa, et al. "Changes in prices after an excise tax to sweetened sugar beverages was implemented in Mexico: evidence from urban areas." *PloS one* 10.12 (2015): e0144408.

Colchero, M. Arantxa, et al. "Changes in prices of taxed sugar-sweetened beverages and nonessential energy dense food in rural and semi-rural areas in Mexico." *Salud publica de Mexico* 59.2 (2017): 137-146.

Colchero, M. Arantxa, et al. "In Mexico, evidence of sustained consumer response two years after implementing a sugar-sweetened beverage tax." *Health Affairs* 36.3 (2017): 564-571.

Colchero, M. Arantxa, et al. "Price elasticity of the demand for sugar sweetened beverages and soft drinks in Mexico." *Economics & Human Biology* 19 (2015): 129-137.

Colchero, M. Arantxa, Shu Wen Ng, and Barry M. Popkin. "Sugar-sweetened beverage tax: the authors reply." *Health Affairs* 36.6 (2017): 1145-1145.

Fuster, Melissa, et al. "Understanding policy change for obesity prevention: learning from sugar-sweetened beverages taxes in Mexico and Chile." *Health Promotion International* 36.1 (2021): 155-164.

Guerrero-López, Carlos M., Mariana Molina, and M. Arantxa Colchero. "Employment changes associated with the introduction of taxes on sugar-sweetened beverages and nonessential energy-dense food in Mexico." *Preventive medicine* 105 (2017): S43-S49.

Hernández-F, Mauricio, et al. "Reduction in purchases of energy-dense nutrient-poor foods in Mexico associated with the introduction of a tax in 2014." *Preventive medicine* 118 (2019): 16-22.

James, Erin, Martín Lajous, and Michael R. Reich. "The politics of taxes for health: an analysis of the passage of the sugar-sweetened beverage tax in Mexico." *Health Systems & Reform* 6.1 (2020): e1669122.

Ng, Shu Wen, et al. "Did high sugar-sweetened beverage purchasers respond differently to the excise tax on sugar-sweetened beverages in Mexico?." *Public health nutrition* 22.4 (2019): 750-756.

Ortega-Avila, Ana G., Angeliki Papadaki, and Russell Jago. "Exploring perceptions of the Mexican sugar-sweetened beverage tax among adolescents in north-west Mexico: a qualitative study." *Public Health Nutrition* 21.3 (2018): 618-626.

Popkin, Barry M. "Mexican cohort study predates but predicts the type of body composition changes expected from the Mexican sugar-sweetened beverage Tax." *American journal of public health* 107.11 (2017): 1702.

Sánchez-Romero, Luz María, et al. "Association between tax on sugar sweetened beverages and soft drink consumption in adults in Mexico: open cohort longitudinal analysis of Health Workers Cohort Study." *bmj* 369 (2020).

Sánchez-Romero, Luz Maria, et al. "Projected impact of Mexico's sugar-sweetened beverage tax policy on diabetes and cardiovascular disease: a modeling study." *PLoS medicine* 13.11 (2016): e1002158.

Taillie, Lindsey Smith, et al. "Do high vs. low purchasers respond differently to a nonessential energy-dense food tax? Two-year evaluation of Mexico's 8% nonessential food tax." *Preventive medicine* 105 (2017): S37-S42.

Torres-Álvarez, Rossana, et al. "Body weight impact of the sugar-sweetened beverages tax in Mexican children: A modeling study." *Pediatric Obesity* 15.8 (2020): e12636.

#### MOVE/meMUEVO

Arellano-Morales, Leticia, Christine M. Wood, and John P. Elder. "Acculturation among Latino primary caregivers and physician communication: receipt of advice regarding healthy lifestyle behaviors." *Journal of community health* 38.1 (2013): 113-119.Corder, Kirsten, et al. "Active children use more locations for PA." *Health & place* 17.4 (2011): 911-919.

Corder, Kirsten, et al. "Parent awareness of young children's PA." *Preventive medicine* 55.3 (2012): 201-205.

Corder, Kirsten, et al. "Predictors of change in sports participation in Latino and non-Latino children." *British journal of sports medicine* 46.9 (2012): 684-688.

Crespo, Noe C., et al. "An examination of multilevel factors that may explain gender differences in children's PA." *Journal of PA and Health* 10.7 (2013): 982-992.

Eisenberg, Christina M., et al. "Examining multiple parenting behaviors on young children's dietary fat consumption." *Journal of Nutrition Education and Behavior* 44.4 (2012): 302-309.

* Elder, John P., et al. "Childhood obesity prevention and control in city recreation centres and family homes: the MOVE/me M uevo P roject." *Pediatric obesity* 9.3 (2014): 218-231.

Lopez, Nanette V., et al. "Parent support and parent-mediated behaviors are associated with children's sugary beverage consumption." *Journal of the Academy of Nutrition and Dietetics* 112.4 (2012): 541-547.

Morello, Monica I., et al. "Associations among parent acculturation, child BMI, and child fruit and vegetable consumption in a Hispanic sample." *Journal of Immigrant and Minority Health* 14.6 (2012): 1023-1029.

#### Netherlands Salt Reduction Strategy

* Hendriksen, Marieke AH, et al. "Monitoring salt and iodine intakes in Dutch adults between 2006 and 2010 using 24 h urinary sodium and iodine excretions." *Public health nutrition* 17.7 (2014): 1431-1438.

Hendriksen, M. A. H., E. C. Wilson-van den Hooven, and D. L. van der A. "Zout-en jodiuminname 2010: Voedingsstatusonderzoek bij volwassenen uit Doetinchem." (2011).

Temme, Elisabeth HM, et al. "Salt reductions in some foods in the Netherlands: monitoring of food composition and salt intake." *Nutrients* 9.7 (2017): 791.

#### OY Bike

* Noland, Robert B., and Muhammad M. Ishaque. "Smart bicycles in an urban area: evaluation of a pilot scheme in London." *Journal of Public Transportation* 9.5 (2006): 5.

#### PHRESH

Cantor, Jonathan, et al. "SNAP Participants Improved Food Security And Diet After A Full-Service Supermarket Opened In An Urban Food Desert: Study examines impact grocery store opening had on food security and diet of Supplemental Nutrition Assistance Program participants living in an urban food desert." *Health Affairs* 39.8 (2020): 1386-1394.

Cohen, Deborah A., et al. "Store impulse marketing strategies and body mass index." *American journal of public health* 105.7 (2015): 1446-1452.

Dubowitz, Tamara, et al. "A natural experiment opportunity in two low-income urban food desert communities: research design, community engagement methods, and baseline results." *Health Education & Behavior* 42.1_suppl (2015): 87S-96S.

Dubowitz, T., et al. "A new supermarket in a food desert: Is better health in store." *Santa Monica, CA: RAND Corporation* (2015).

* Dubowitz, Tamara, et al. "Changes in diet after introduction of a full service supermarket in a food desert." *Health Affairs (Project Hope)* 34.11 (2015): 1858.

Dubowitz, Tamara, et al. *Community Development Can Improve Resident Health*. RAND, 2018.

Dubowitz, Tamara, et al. "Diet and perceptions change with supermarket introduction in a food desert, but not because of supermarket use." *Health Affairs* 34.11 (2015): 1858-1868.

Dubowitz, Tamara, et al. "Healthy food access for urban food desert residents: examination of the food environment, food purchasing practices, diet and BMI." *Public health nutrition* 18.12 (2015): 2220-2230.

Dubowitz, Tamara, et al. "Using a grocery list is associated with a healthier diet and lower BMI among very high-risk adults." *Journal of nutrition education and behavior* 47.3 (2015): 259-264.

Ghosh-Dastidar, Bonnie, et al. "Distance to store, food prices, and obesity in urban food deserts." *American journal of preventive medicine* 47.5 (2014): 587-595.

Ghosh-Dastidar, Madhumita, et al. "Does opening a supermarket in a food desert change the food environment?." *Health & place* 46 (2017): 249-256.

Parisi, Sara M., Lisa M. Bodnar, and Tamara Dubowitz. "Weight resilience and fruit and vegetable intake among African-American women in an obesogenic environment." *Public health nutrition* 21.2 (2018): 391-402.

Richardson, Andrea S., et al. "Can the introduction of a full-service supermarket in a food desert improve residents' economic status and health?." *Annals of epidemiology* 27.12 (2017): 771-776.

Richardson, Andrea S., et al. "Improvements in neighborhood socioeconomic conditions may improve resident diet." *American journal of epidemiology* 190.5 (2021): 798-806.

Vaughan, Christine A., et al. "Does where you shop or who you are predict what you eat?: The role of stores and individual characteristics in dietary intake." *Preventive medicine* 100 (2017): 10-16.

Vaughan, Christine A., et al. "Where do food desert residents buy most of their junk food? Supermarkets." *Public health nutrition* 20.14 (2017): 2608-2616.

#### POP project

* De Coen, Valerie, et al. "Effects of a 2-year healthy eating and PA intervention for 3–6-year-olds in communities of high and low socio-economic status: the POP (Prevention of Overweight among Pre-school and school children) project." *Public health nutrition* 15.9 (2012): 1737-1745.

De Coen, Valerie, et al. "Parental socioeconomic status and soft drink consumption of the child. The mediating proportion of parenting practices." *Appetite* 59.1 (2012): 76-80.

De Coen, Valerie, et al. "Risk factors for childhood overweight: a 30-month longitudinal study of 3-to 6-year-old children." *Public health nutrition* 17.9 (2014): 1993-2000.

#### Project Joy

Patt, Madhavi R., et al. "Assessment of global coronary heart disease risk in overweight and obese African-American women." *Obesity research* 11.5 (2003): 660-667.

Patt, Madhavi R., et al. "Body image assessment: comparison of figure rating scales among urban Black women." *Ethnicity & disease* 12.1 (2002): 54-62.

* Yanek, Lisa R., et al. "Project Joy: faith based cardiovascular health promotion for African American women." *Public health reports* 116.Suppl 1 (2001): 68.

#### Quest to Lava Mountain

Beasley, Nancy, et al. "The Quest to Lava Mountain: Using video games for dietary change in children." *Journal of the Academy of Nutrition and Dietetics (Print)* 112.9 (2012): 1334-1336.

Chow, Joanne. *Computer use and food consumption pattern among economically disadvantaged children in Texas*. Diss. The University of Texas School of Public Health, 2013.

* Sharma, Shreela V., et al. "Effects of the Quest to Lava Mountain computer game on dietary and PA behaviors of elementary school children: a pilot group-randomized controlled trial." *Journal of the Academy of Nutrition and Dietetics* 115.8 (2015): 1260-1271.

#### Rock on Café

* Johnston, Yvonne, et al. "Steps to a healthier New York." *Health Promotion Practice* 10.2_suppl (2009): 100S-108S.

#### Route H

Escoto, Kamisha H., et al. "Work hours, weight status, and weight-related behaviors: a study of metro transit workers." *International Journal of Behavioral Nutrition and PA* 7.1 (2010): 1-10.

French, Simone A., et al. "Association between body weight, PA and food choices among metropolitan transit workers." *International Journal of Behavioral Nutrition and PA* 4.1 (2007): 1-12.

French, Simone A., et al. "Pricing and availability intervention in vending machines at four bus garages." *Journal of occupational and environmental medicine/American College of Occupational and Environmental Medicine* 52.Suppl 1 (2010): S29.

* French, Simone A., et al. "Worksite environment intervention to prevent obesity among metropolitan transit workers." *Preventive medicine* 50.4 (2010): 180-185.

Shimotsu, Scott T., et al. "Worksite environment PA and healthy food choices: measurement of the worksite food and PA environment at four metropolitan bus garages." *International Journal of Behavioral Nutrition and PA* 4.1 (2007): 1-8.

#### Ruelle

Ruelle, Morgan L., and Karim–Aly S. Kassam. "Diversity of plant knowledge as an adaptive asset: a case study with standing rock elders1." *Economic Botany* 65.3 (2011): 295-307.

Ruelle, Morgan L. "Ecological relations and Indigenous food sovereignty in standing rock." *American Indian Culture and Research Journal* 41.3 (2017): 113-125.

Ruelle, Morgan L., and Karim-Aly S. Kassam. "Foodways transmission in the standing Rock Nation." *Food and Foodways* 21.4 (2013): 315-339.

Ruelle, Morgan. "Plants and Foodways of the Standing Rock Nation: Diversity, Knowledge, and Sovereignty." (2011).

* Ruelle, Morgan, Stephen J. Morreale, and Karim-Aly S. Kassam. "Practicing food sovereignty: Spatial analysis of an emergent food system for the Standing Rock Nation." *Journal of Agriculture, Food Systems, and Community Development* 2.1 (2011): 163-179.

#### Safdie

Aburto, Nancy Jennings, et al. "Effect of a school-based intervention on PA: cluster-randomized trial." *Medicine and science in sports and exercise* 43.10 (2011): 1898-1906.

Safdie, Margarita, et al. "An ecological and theoretical deconstruction of a school-based obesity prevention program in Mexico." *International Journal of Behavioral Nutrition and PA* 11.1 (2014): 1-10.

Safdie, Margarita. *Childhood Obesity Prevention Intervention and Policy in the Mexican School System*. Diss. 2013.

* Safdie, Margarita, et al. "Impact of a school-based intervention program on obesity risk factors in Mexican children." *Salud publica de Mexico* 55.suppl 3 (2013): 374-387.

Safdie, Margarita, et al. "Promoting healthful diet and PA in the Mexican school system for the prevention of obesity in children." *salud pública de méxico* 55 (2013): 357-373.

#### Saksvig

Ho, Lara S., et al. "An integrated multi-institutional diabetes prevention program improves knowledge and healthy food acquisition in northwestern Ontario First Nations." *Health Education & Behavior* 35.4 (2008): 561-573.

Ho, L. S., et al. "Development of an integrated diabetes prevention program with First Nations in Canada." *Health Promotion International* 21.2 (2006): 88-97.

Kakekagumick, Kara E., et al. "Sandy lake health and diabetes project: a community-based intervention targeting type 2 diabetes and its risk factors in a first nations community." *Frontiers in endocrinology* 4 (2013): 170.

Macaulay, Ann C., et al. "Primary prevention of type 2 diabetes: experiences of 2 aboriginal communities in Canada." *Can J Diabetes* 27.4 (2003): 464-75.

* Saksvig, Brit I., et al. "A pilot school-based healthy eating and PA intervention improves diet, food knowledge, and self-efficacy for native Canadian children." *The Journal of nutrition* 135.10 (2005): 2392-2398.

#### Secrets to the Good Life

Ayala, Guadalupe X., et al. "Nutrition communication for a Latino community: Formative research foundations." *Family and Community Health* (2001): 72-87.

Ayala, Guadalupe X., et al. "Tuesday, November 14, 2000-Board 5 Abstract# 12896 Development of a communication-based nutrition intervention for Latinas." *The 128th Annual Meeting of APHA*. 2000.

Baquero, Barbara, et al. "Secretos de la Buena Vida: processes of dietary change via a tailored nutrition communication intervention for Latinas." *Health Education Research* 24.5 (2009): 855-866.

Elder, John P., et al. "Interpersonal and print nutrition communication for a Spanish-dominant Latino population: Secretos de la Buena Vida." *Health Psychology* 24.1 (2005): 49.

* Elder, John P., et al. "Long-term effects of a communication intervention for Spanish-dominant Latinas." *American Journal of Preventive Medicine* 31.2 (2006): 159-166.

#### Shape Up Rhode Island

* Leahey, Tricia M., et al. "Effect of teammates on changes in PA in a statewide campaign." *Preventive medicine* 51.1 (2010): 45-49.

Wing, Rena R., et al. "A statewide intervention reduces BMI in adults: Shape Up Rhode Island results." *Obesity* 17.5 (2009): 991-995.

#### Slusser

* Slusser, Wendy, et al. "Pediatric overweight prevention through a parent training program for 2–4 year old Latino children." *Childhood Obesity (Formerly Obesity and Weight Management)* 8.1 (2012): 52-59.

#### Small steps are easier together

Devine, Carol M., et al. "Process evaluation of an environmental walking and healthy eating pilot in small rural worksites." *Evaluation and Program Planning* 35.1 (2012): 88-96.

* Warren, Barbour S., et al. "Small steps are easier together: a goal-based ecological intervention to increase walking by women in rural worksites." *Preventive Medicine* 50.5-6 (2010): 230-234.

#### Shandong Ministry of Health Action on Salt Reduction and Hypertension (SMASH)

* Bi, Zhenqiang, et al. "Peer Reviewed: Hypertension Prevalence, Awareness, Treatment, and Control and Sodium Intake in Shandong Province, China: Baseline Results From Shandong–Ministry of Health Action on Salt Reduction and Hypertension (SMASH), 2011." *Preventing Chronic Disease* 11 (2014).

Chen, Xi, et al. "RETRACTED: Urinary sodium or potassium excretion and blood pressure in adults of Shandong province, China: preliminary results of the SMASH project." (2015): 754-762.

Hou, Lei, et al. "Associations between salt-restriction spoons and long-term changes in urinary Na+/K+ ratios and blood pressure: findings from a population-based cohort." *Journal of the American Heart Association* 9.14 (2020): e014897.

Xu, Aiqiang, et al. "Association of a province-wide intervention with salt intake and hypertension in shandong province, China, 2011-2016." *JAMA internal medicine* 180.6 (2020): 877-886.

Xu, Chunxiao, et al. "Iodine nutritional status in the adult population of Shandong Province (China) prior to salt reduction program." *European journal of nutrition* 55.5 (2016): 1933-1941.

Xu, Jianwei, et al. "Associations of usual 24-hour sodium and potassium intakes with blood pressure and risk of hypertension among adults in China’s Shandong and Jiangsu Provinces." *Kidney and Blood Pressure Research* 42.1 (2017): 188-200.

Yan, Liuxia, et al. "Relationships between blood pressure and 24-hour urinary excretion of sodium and potassium by body mass index status in Chinese adults." *The Journal of Clinical Hypertension* 17.12 (2015): 916-925.

Zhang, Jiyu, et al. "Cardiovascular diseases deaths attributable to high sodium intake in Shandong Province, China." *Journal of the American Heart Association* 8.1 (2019): e010737.

Zhang, Juan, et al. "Inaccuracy of self-reported low sodium diet among Chinese: Findings from baseline survey for Shandong & Ministry of Health Action on Salt and Hypertension (SMASH) project." *Biomedical and Environmental Sciences* 28.2 (2015): 161-167.

#### South Carolina Farmers Market

* Kunkel, Mary Elizabeth, Barbara Luccia, and Archie C. Moore. "Evaluation of the South Carolina seniors farmers' market nutrition education program." *Journal of the American Dietetic Association* 103.7 (2003): 880-883.

#### Speck

Hines-Martin, Vicki P., et al. "Building Rapport and Successful Recruitment in a Low-Income, Underserved Community." *37th Biennial Convention-Scientific Session*. 2003.

Hines-Martin, Vicki, et al. "Understanding systems and rhythms for minority recruitment in intervention research." *Research in nursing & health* 32.6 (2009): 657-670.

Speck, Barbara, et al. "Advanced registered nurse practitioner as facilitator of a PA intervention with low-income women." *Annual Meeting*.

* Speck, Barbara J., et al. "An environmental intervention aimed at increasing PA levels in low-income women." *Journal of Cardiovascular Nursing* 22.4 (2007): 263-271.

#### Steps to a healthier Arizona

* Drummond, Rebecca L., et al. "Steps to a healthier Arizona." *Health Promotion Practice* 10.2_suppl (2009): 156S-167S.

#### Thai Salt Reduction Strategy

* Supornsilaphachai, C. "Evolution of salt reduction initiatives in Thailand: lessons for other countries in the South-East Asia Region." *WHO Regional Health Forum*. Vol. 17. No. 1. 2013.

#### Tioga County Fit for Life

* Gombosi, Russell L., Regina M. Olasin, and Jason L. Bittle. "Tioga County Fit for Life: a primary obesity prevention project." *Clinical pediatrics* 46.7 (2007): 592-600.

#### TravelSMART schools

Di Pietro, G., and Ian Hughes. "TravelSMART schools: there really is a better way to go!." *WIT Transactions on Ecology and the Environment* 72 (2004).

Hughes, Ian, and Gayle Di Pietro. "Ongoing development of the TravelSmart curriculum project in Victoria." *Proceedings of the 28th Australasian Transport Research Forum. Sydney: PATREC. http://atrf. info/papers/2005/2005_Hughes_DiPietro_b. pdf*. 2005.

* Moodie, Marj, et al. "Assessing cost-effectiveness in obesity: active transport program for primary school children—TravelSMART schools curriculum program." *Journal of PA and health* 8.4 (2011): 503-515.

#### UK National Cycle Network

* Jones, Tim. "Getting the British back on bicycles—The effects of urban traffic-free paths on everyday cycling." *Transport Policy* 20 (2012): 138-149.

Jones, Tim. *The role of national cycle network traffic-free paths in creating a cycling culture: the case of NCN Route 5 Stafford*. Diss. Oxford Brookes University, 2008.

#### UK Salt Reduction Strategy

Brinsden, Hannah C., et al. "Surveys of the salt content in UK bread: progress made and further reductions possible." *BMJ open* 3.6 (2013): e002936.

Collins, Marissa, et al. "An economic evaluation of salt reduction policies to reduce coronary heart disease in England: a policy modeling study." *Value in Health* 17.5 (2014): 517-524.

Eyles, Helen, et al. "Impact of the UK voluntary sodium reduction targets on the sodium content of processed foods from 2006 to 2011: Analysis of household consumer panel data." *Preventive medicine* 57.5 (2013): 555-560.

* He, Feng J., Sonia Pombo-Rodrigues, and Graham A. MacGregor. "Salt reduction in England from 2003 to 2011: its relationship to blood pressure, stroke and ischaemic heart disease mortality." *BMJ open* 4.4 (2014): e004549.

He, F. J., H. C. Brinsden, and G. A. MacGregor. "Salt reduction in the United Kingdom: a successful experiment in public health." *Journal of human hypertension* 28.6 (2014): 345-352.

Jaworowska, Agnieszka, et al. "Determination of salt content in hot takeaway meals in the United Kingdom." *Appetite* 59.2 (2012): 517-522.

MacGregor, Graham A., and Kawther M. Hashem. "Action on sugar—lessons from UK salt reduction programme." *The Lancet* 383.9921 (2014): 929-931.

MacGregor, Graham A., Feng J. He, and Sonia Pombo-Rodrigues. "Food and the responsibility deal: how the salt reduction strategy was derailed." *Bmj* 350 (2015).

Marshall, Stephanie, John A. Bower, and Monika JA Schröder. "Consumer understanding of UK salt intake advice." *British Food Journal* (2007).

Millett, Christopher, et al. "Impacts of a national strategy to reduce population salt intake in England: serial cross sectional study." *PLoS One* 7.1 (2012): e29836.

Shankar, Bhavani, et al. "An evaluation of the UK Food Standards Agency's salt campaign." *Health Economics* 22.2 (2013): 243-250.

Sharma, Abhijit, Salvatore Di Falco, and Iain Fraser. "Consumption of salt rich products: impact of the UK reduced salt campaign." *International Journal of Health Economics and Management* 19.3 (2019): 341-357.

Srinivasan, C. S., and Guiseppe Nocella. *Contribution of Product Reformulation to the EU Salt Campaign: Empirical Evidence from the UK*. No. 358-2016-18346. 2016.

Sutherland, Jennifer, et al. "Fewer adults add salt at the table after initiation of a national salt campaign in the UK: a repeated cross-sectional analysis." *British Journal of Nutrition* 110.3 (2013): 552-558.

World Health Organization. "Creating an enabling environment for population-based salt reduction strategies: report of a joint technical meeting held by WHO and the Food Standards Agency, United Kingdom, July 2010." (2010).

Wyness, Laura A., Judith L. Butriss, and Sara A. Stanner. "Reducing the population's sodium intake: the UK Food Standards Agency's salt reduction programme." *Public health nutrition* 15.2 (2012): 254-261.

#### University of Bristol Travel Plan

* Brockman, R., and K. R. Fox. "PA by stealth? The potential health benefits of a workplace transport plan." *Public health* 125.4 (2011): 210-216.

#### VIASANO

Borys, J., et al. "Childhood obesity prevention: A significant decrease of overweight and obesity in the VIASANO programme after 2 years of intervention." *Journal of Science and Medicine in Sport* 15 (2012): S331.

* Vinck, Jan, et al. "Downward trends in the prevalence of childhood overweight in two pilot towns taking part in the VIASANO community-based programme in Belgium: data from a national school health monitoring system." *Pediatric obesity* 11.1 (2016): 61-67.

Vinck, J., et al. "PA promotion in a community based programme to reduce overweight prevalence in Belgian children." *Science & Sports* 29 (2014): S9.

#### Walk to work day campaign

* Merom, Dafna, et al. "Effect of Australia's Walk to Work Day campaign on adults' active commuting and PA behavior." *American Journal of Health Promotion* 19.3 (2005): 159-162.

Merom, Dafna, et al. "Predictors of initiating and maintaining active commuting to work using transport and public health perspectives in Australia." *Preventive medicine* 47.3 (2008): 342-346.

#### Walk your heart to health

Izumi, Betty T., et al. "Leader behaviors, group cohesion, and participation in a walking group program." *American journal of preventive medicine* 49.1 (2015): 41-49.

Kwarteng, Jamila L., et al. "Does perceived safety modify the effectiveness of a walking-group intervention designed to promote PA?." *American Journal of Health Promotion* 32.2 (2018): 423-431.

LeBron, Alana MW, et al. "Storytelling in community intervention research: lessons learned from the walk your heart to health intervention." *Progress in community health partnerships: research, education, and action* 8.4 (2014): 477.

Schulz, Amy, et al. "Building the healthiest nation by promoting health in vulnerable communities: Impact and outcome evaluation of the Walk Your Heart to Health community-based multilevel intervention." *APHA 2016 Annual Meeting & Expo (Oct. 29-Nov. 2, 2016)*. APHA, 2016.

Schulz, Amy J., et al. "Do neighborhood demographics modify walking group intervention effectiveness in urban neighborhoods?." *Health promotion practice* 18.1 (2017): 62-74.

* Schulz, Amy J., et al. "Effectiveness of a walking group intervention to promote PA and cardiovascular health in predominantly non-Hispanic black and Hispanic urban neighborhoods: findings from the walk your heart to health intervention." *Health Education & Behavior* 42.3 (2015): 380-392.

#### Walkadoo

* Poirier, Josée, et al. "Effectiveness of an activity tracker-and internet-based adaptive walking program for adults: a randomized controlled trial." *Journal of medical Internet research* 18.2 (2016): e5295.

#### Wang

Wang, Guijing, et al. "A cost-benefit analysis of PA using bike/pedestrian trails." *Health promotion practice* 6.2 (2005): 174-179.

Wang, Guijing, et al. "Cost analysis of the built environment: the case of bike and pedestrian trials in Lincoln, Neb." *American Journal of Public Health* 94.4 (2004): 549-553.

* Wang, Guijing, et al. "Cost effectiveness of a bicycle/pedestrian trail development in health promotion." *Preventive Medicine* 38.2 (2004): 237-242.

**Wen-2005**

* Wen, Li Ming, et al. "Promoting active transport in a workplace setting: evaluation of a pilot study in Australia." *Health promotion international* 20.2 (2005): 123-133.

**West Virginia Healthy Lifestyles**

* Harris, Carole V., et al. "Evaluating the West Virginia healthy lifestyles act: Methods and procedures." *Journal of PA and Health* 7.s1 (2010): S31-S39.

Matthews-Ewald, Molly R., et al. "Assessing moderate to vigorous PA in rural West Virginia elementary school physical education classes." *West Virginia Medical Journal* 109.4 (2013): 12-17.

**Winning with Wellness**

Dalton III, William T., Karen Schetzina, and Elizabeth Conway-Williams. "A coordinated school health approach to obesity prevention among Appalachian youth: middle school student outcomes from the winning with wellness project." *International Journal of Health Sciences Education* 2.1 (2014): 2.

Schetzina, Karen E., et al. "A coordinated school health approach to obesity prevention among Appalachian youth: the Winning with Wellness Pilot Project." *Family and Community Health* 32.3 (2009): 271-285.

* Schetzina, Karen E., et al. "Developing a coordinated school health approach to child obesity prevention in rural Appalachia: results of focus groups with teachers, parents, and students." *Rural and Remote Health* 9.4 (2009): 1-13.

Schetzina, Karen E., et al. "The" Winning With Wellness" Pilot Project: Rural Appalachian Elementary Student PA and Eating Behaviors and Program Implementation 4 Years Later." *Family and Community Health* 34.2 (2011): 154-162.

**ZATP**

* Ho, Lara S., et al. "An integrated multi-institutional diabetes prevention program improves knowledge and healthy food acquisition in northwestern Ontario First Nations." *Health Education & Behavior* 35.4 (2008): 561-573.

Ho, Lara S. *Diabetes prevention in northwestern Ontario first nations: a multi-institutional program to improve diet and increase PA*. The Johns Hopkins University, 2007.

Ho, Lara, et al. "Food-related behavior, PA, and dietary intake in First Nations1–a population at high risk for diabetes." *Ethnicity & health* 13.4 (2008): 335-349.

Rosecrans, A. M., et al. "Process evaluation of a multi-institutional community-based program for diabetes prevention among First Nations." *Health education research* 23.2 (2008): 272-286.

Sharma, Sangita, et al. "Dietary intake and development of a quantitative food-frequency questionnaire for a lifestyle intervention to reduce the risk of chronic diseases in Canadian First Nations in north-western Ontario." *Public health nutrition* 11.8 (2008): 831-840.

### Supplementary Appendix 1.5 – Detailed description of draft framework

#### Conceptual diagram underpinning the draft framework

Figure 1.5A presents a conceptual diagram outlining the key concepts underpinning the draft framework. It should be read alongside the definitions and examples in Table 1.5A.

##### *Actors*

In Figure 1.5A, actors involved in PHIs are listed on the far left. Actors are individuals or representatives of groups or organisations required to perform an action within the intervention sequence to change an individuals' diet or PA. We identify: actors who operate within macro- or micro-environments, and informal gatekeepers in the individual environment. The primary recipient is the individual whose diet or activity practice the intervention is trying to change. The figure lists all possible actors. Not all actors will be involved in every intervention. Actors vary in their potential population reach with those placed higher in Figure 1.5A having greater potential reach. For example actions by a town planner (macro-environmental actor) can impact on everyone in a city, whereas actions by a parent (individual gatekeeper) may only impact on a few children.

##### *Actions*

The actions required of actors to achieve intended intervention effects are shown in the middle and right of Figure 1.5A. Actors within macro-, micro- and individual environments make use of their resources to bring about environmental change. Interventions may then require primary recipients to complete a further sequence of actions to achieve diet or PA change. All actions occur within a unique context, which may place additional resource demands on actors. The nature of the environmental change or intervention will determine what actions are required from primary recipients, and the demand these actions place on their resources. Individual behaviour change theory is applicable within this section of the framework but we do not detail psychological mechanisms here. Our framework is not intended to replace existing individual behaviour change theory, nor do we advocate a particular theory.

##### *Exposure*

In order for an intervention to change diet or PA, primary recipients must first be exposed to it. Exposure can be routine or non-routine. Routine exposure requires little or no additional action from primary recipients, meaning limited agency demands. In contrast, non-routine exposure requires primary recipients to act to become exposed, and hence place higher agency demands on them.

##### *Interaction*

Once exposed, primary recipients engage with the intervention or environmental change through conscious or non-conscious mechanisms. Non-conscious mechanisms require no conscious deliberation (Marteau, Hollands and Fletcher, 2012) and therefore place low agency demands on primary recipients. Conscious mechanisms require primary recipients to engage in, and act on, reflective processes (Marteau, Hollands and Fletcher, 2012) and so have higher agency demands.

##### *Intervention target*

An intervention can aim to change one of three targets in primary recipients:

(1) **Resources**. Interventions may change the resources available to primary recipients to facilitate a change in diet or PA. Changing resources alone is insufficient to change behaviour and individuals must make use of their agency to use changed resources to achieve behaviour change.

(2) **Intermediate behavioural steps**. Interventions may aim to change a behaviour or cognitive strategy that enables or prepares a primary recipient to change diet or PA. It is possible, indeed highly likely, that this pathway will comprise of multiple behaviours, varying in length and complexity dependent on the intervention. Thus the actions required from individuals will be variable.

(3) **Diet or PA practice.** Environmental changes that directly target changes to diet or PA require no action from primary recipients and therefore do not place agency demands on primary recipients for them to benefit.

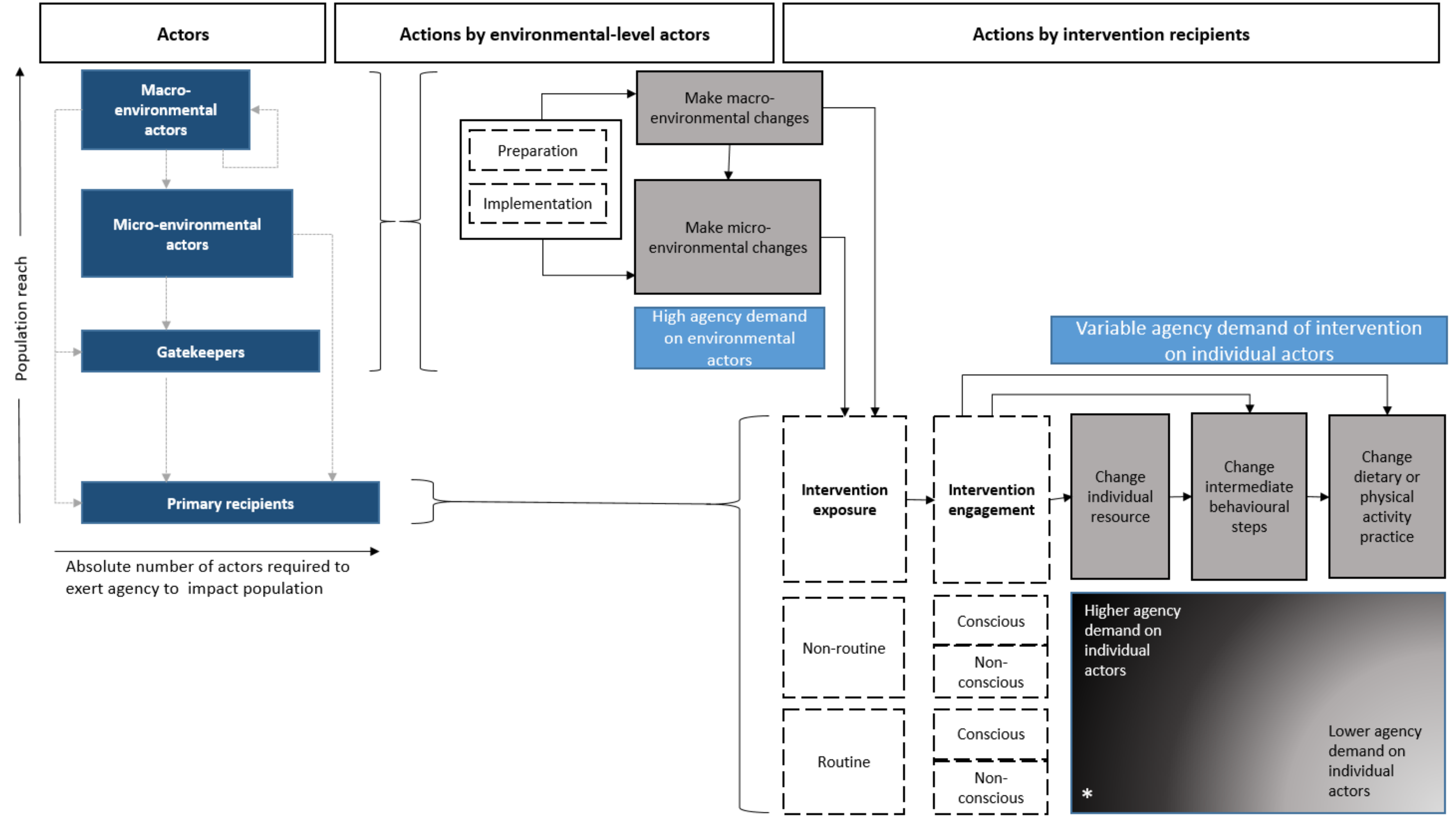


**Figure 1.5A** – *Conceptual diagram to describe the actors, their actions and resulting agency demands of PHIs to promote a healthy diet and PA.*

* Not all combinations have been identified in the literature and to our knowledge hypothesised to be possible. Only plausible and hypothesised combinations are included in the final framework and its application guide.

*Table 1.5A – Definitions and examples of components in Figures 1.5A and 1.5B*

| Component | Definition | Example |
|---|---|---|
| Consider a *single intervention component* | A single pathway or chain of action within an intervention with an intended ultimate endpoint of diet or PA change | Three components of a multi-component school based intervention: (1) changes to canteen menu; (2) nutrition education in curriculum; (3) provision of additional PA equipment at break times |
| **STEP 1 – DETERMINE ENVIRONMENTAL AGENCY DEMAND**<br>Does the intervention… | | |
| …require a gatekeeper to act? | A Gatekeeper is as an individual required to change their behaviour to influence the diet or PA practice of the primary recipient. This action is reflects what could be seen as their personal or citizen (but not professional) responsibilities | Parents change what groceries they buy or how they take children to school; |
| …involve *micro-environmental actors*? | Micro-environmental settings are places or naturally occurring groups of places where people gather for specific purposes. They are usually geographically distinct, relatively small and influenced by individuals.<br>Micro-environmental actors have authority to decide whether and how an intervention is implemented (decision makers) *or* are responsible for implementing (implementers) in micro-environmental settings. | Workplace manager (decision maker) decides to implement healthier eating policy and canteen staff (implementers) change recipes to reduce salt content of foods.<br><br>Church volunteers serve healthier food at church events |
| …involve *macro-environmental actors*? | Macro-environmental sectors are industries, services or supporting infrastructure, which influence the food eaten or PA carried out. They can operate at international, national or local level.<br>Macro-environmental actors are individuals or groups with a professional role an industry, service or supporting infrastructure that influence diet or PA. | Individuals within marketing departments of supermarkets focus on promotion of fruit and vegetables; Town planner implements design code that encourages walking and cycling; |
| **STEP 2 – DETERMINE THE DISTRIBUTION OF AGENCY DEMANDS** | | |
| …target a change in *individual resource?* | Interventions that aim to add or remove personal resource (e.g. financial, cognitive, social or material) to facilitate dietary or PA change | Food vouchers; provision of PA equipment; facilitation of peer support; provision of information |
| …target a change in *intermediate steps?* | Interventions that aim to change behaviours that prepare or enable an individual to change their diet or PA | Change supermarket layout to influence food selection; lighting on cycle path to increase perceived safety |
| …directly target a change in *dietary or PA behaviour?* | Practice of doing the behaviour, consuming a target food or engaging in PA. | Reduce fat content of foods served in canteens; Change structure of PE lesson to increase time spent in vigorous PA. |
| …require *non-routine or routine interaction?* | *Non-routine:* Requires an individual to make a change to their existing tasks or destinations to be exposed to the intervention. | After school PA sessions; Cooking skill development sessions at community centre. |
| | *Routine:* Requires little or no deviation from the individuals existing tasks or destinations. | Changes to workplace canteen; Inclusion of training materials in routine workplace induction. |
| …require *conscious or non-conscious processing?* | *Non-conscious:* No action required from primary recipients for them to benefit from intervention | Removal of less healthy products from vending machines. |
| | *Conscious:* Primary recipients must engage in reflective process and act upon these to benefit from the intervention | Provision of changing facilities to support cycling to work. |

Interventions that target resources have the longest chain of action because they also require primary recipients to complete intermediate steps, and finally change their diet or PA practice. Interventions targeting intermediate steps have shorter chains of action, and those targeting a change in diet or PA practice require no action from individuals.

Taken as a whole, the nature of the exposure, interaction and intervention target of an intervention has implications for actions required from primary recipients. To exemplify the extremes, an intervention requiring non-routine exposure, conscious interaction, and targeting a change in individual resource will demand many actions. In comparison, an intervention for which the exposure is routine, interaction is non-conscious, and which targets a change in dietary or PA practice will require little or no action from primary recipients. The former make higher agency demands on primary recipients whereas the latter make lower agency demands.

**Draft conceptual framework**

Figure 1.5A introduces the concepts that underpin the draft conceptual framework (figure 1.5B). The framework aims to classify single intervention components according to the environmental actors involved in the intervention and the actions required from primary recipients in order for the intervention to achieve its intended effect. Using the framework, each component of a PHI can be categorised according to the collective action required from all actors involved. We have intentionally not named or scored the categories at this stage.

The framework application tool provides a decision tree to aid application and should be used alongside the definition table and instructions. The decision tree asks users to complete two sets of questions, the first to identify environmental actors and the second to identify actions required from individual recipients.

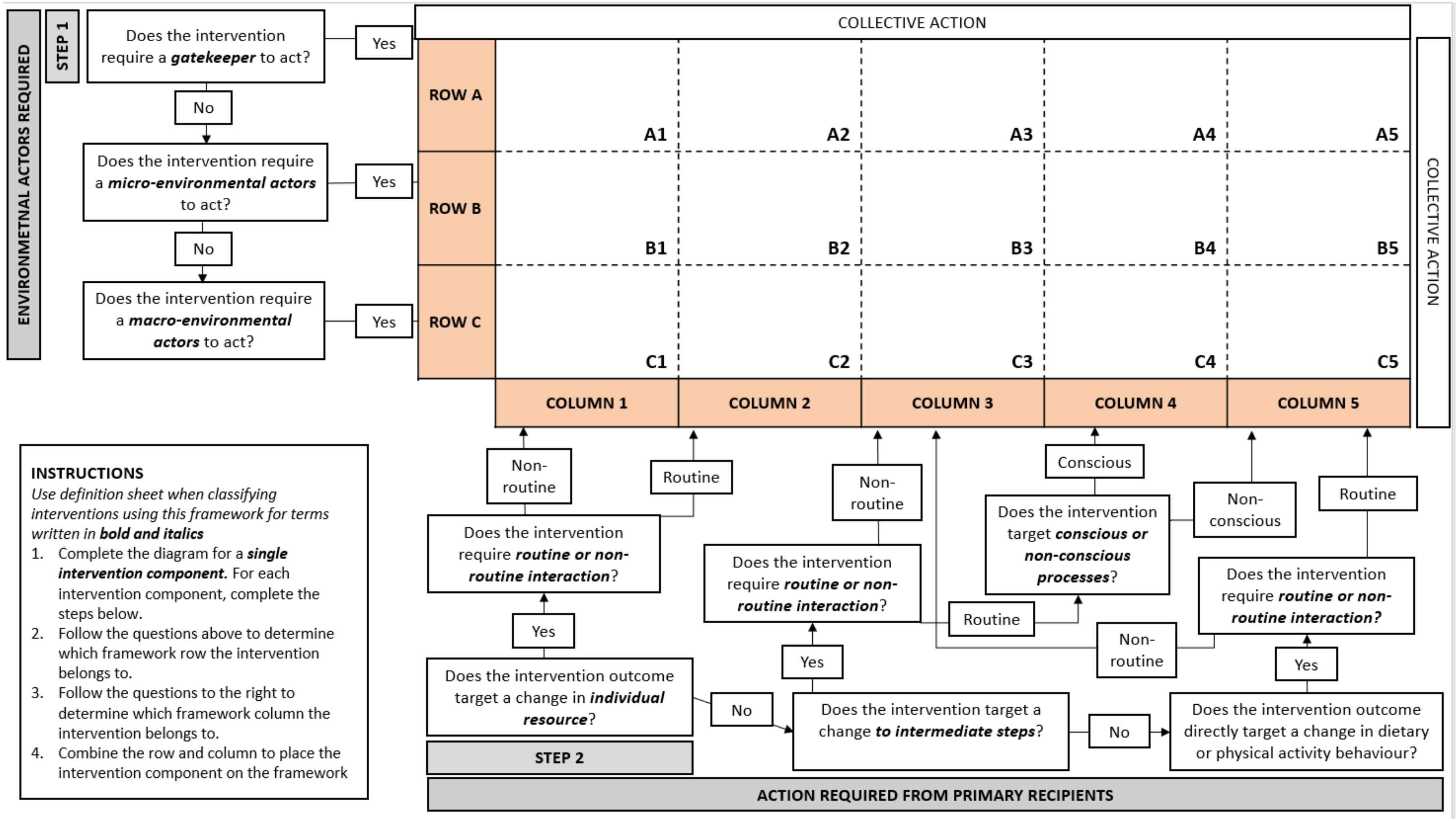


**Figure 1.5B** – *Draft framework*

### Supplementary Appendix 1.6 – Individual intervention component distribution across the DePtH framework

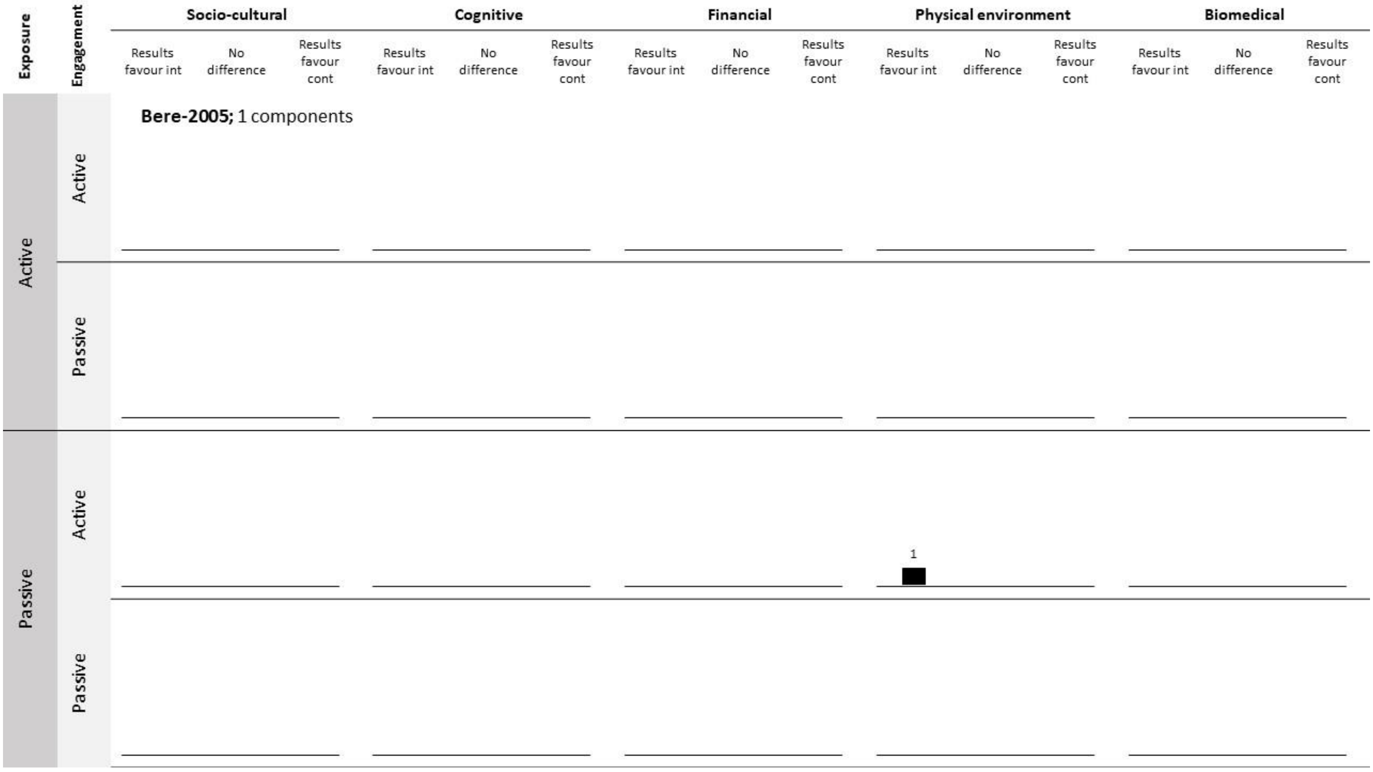


Bere E, Veierød MB, Klepp KI. The Norwegian School Fruit Programme: evaluating paid vs. no-cost subscriptions. Preventive medicine. 2005 Aug 1;41(2):463-70.

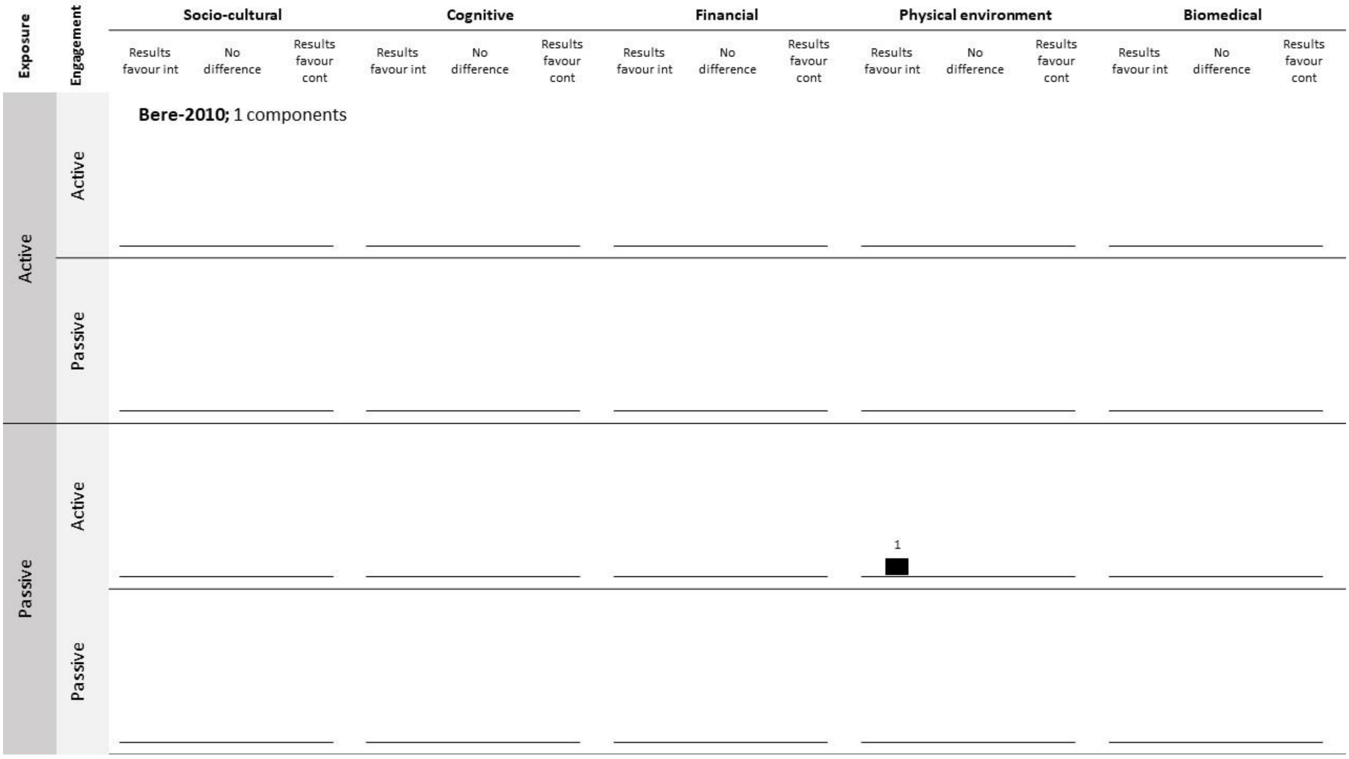


Bere E, Hilsen M, Klepp KI. Effect of the nationwide free school fruit scheme in Norway. British Journal of Nutrition. 2010 Aug;104(4):589-94.

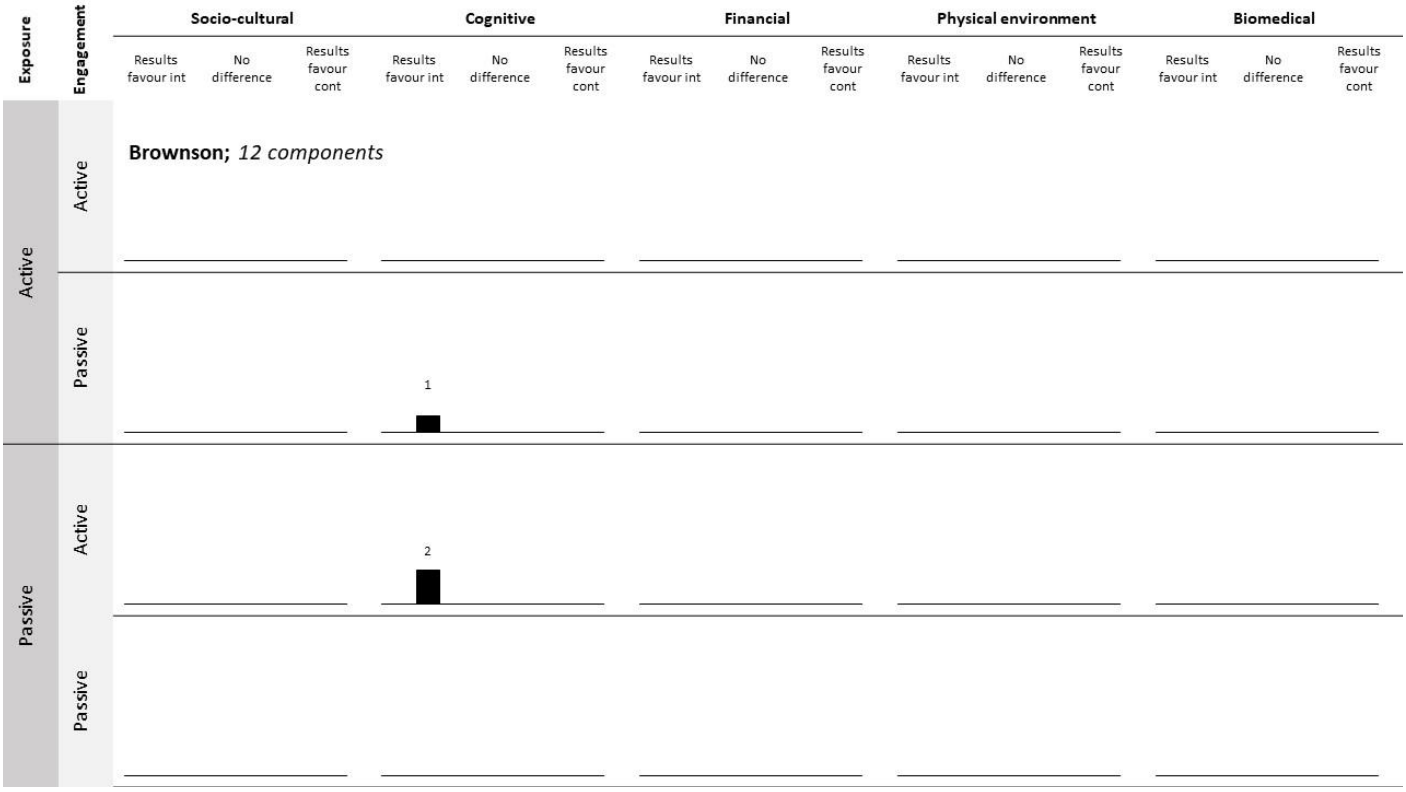


Brownson RC, Smith CA, Pratt M, Mack NE, Jackson-Thompson J, Dean CG, et al. Preventing cardiovascular disease through community-based risk reduction: the Bootheel Heart Health Project. American Journal of Public Health. 1996;86(2):206-13.

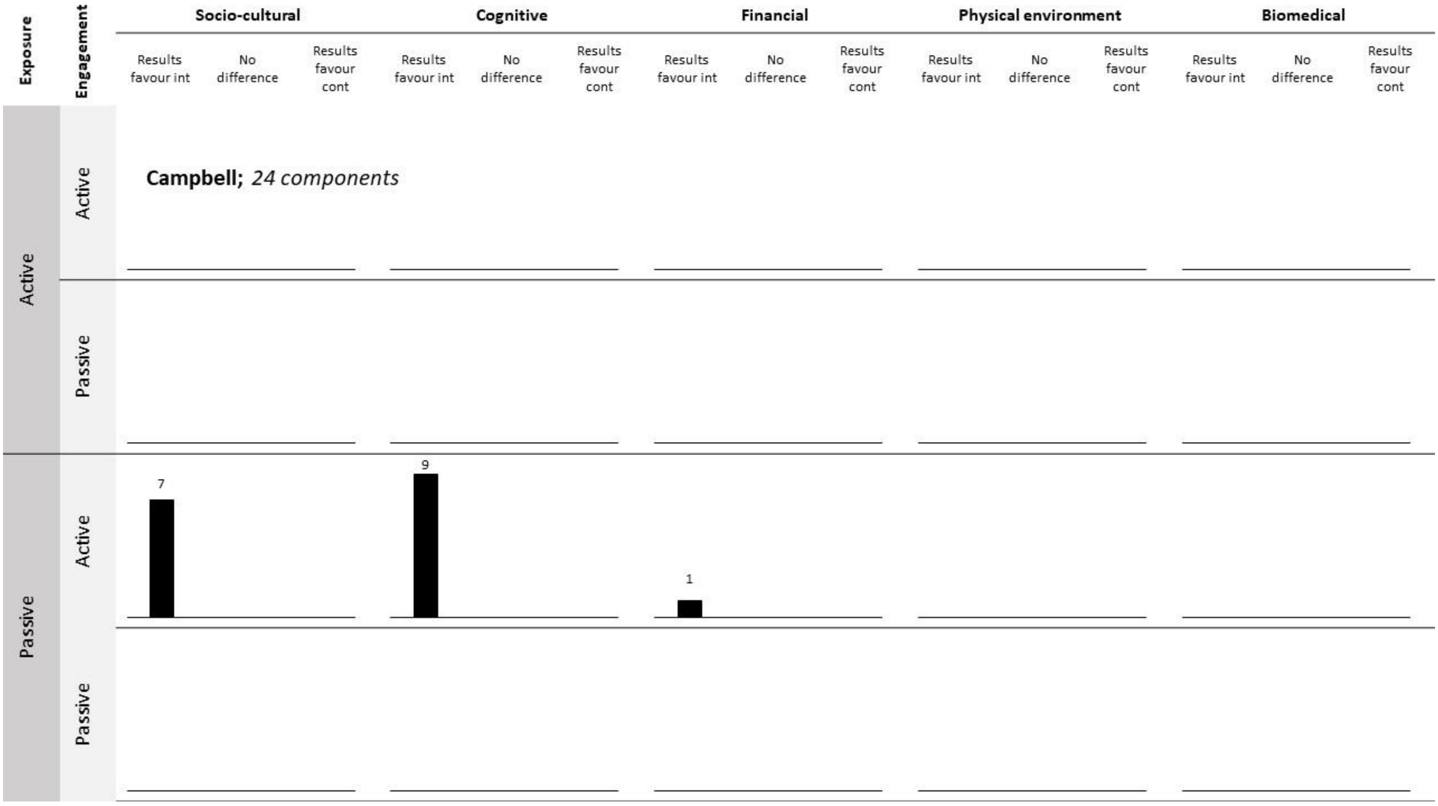


Campbell MK, Demark-Wahnefried W, Symons M, Kalsbeek WD, Dodds J, Cowan A, et al. Fruit and vegetable consumption and prevention of cancer: the Black Churches United for Better Health project. American journal of public health. 1999;89(9):1390-6.

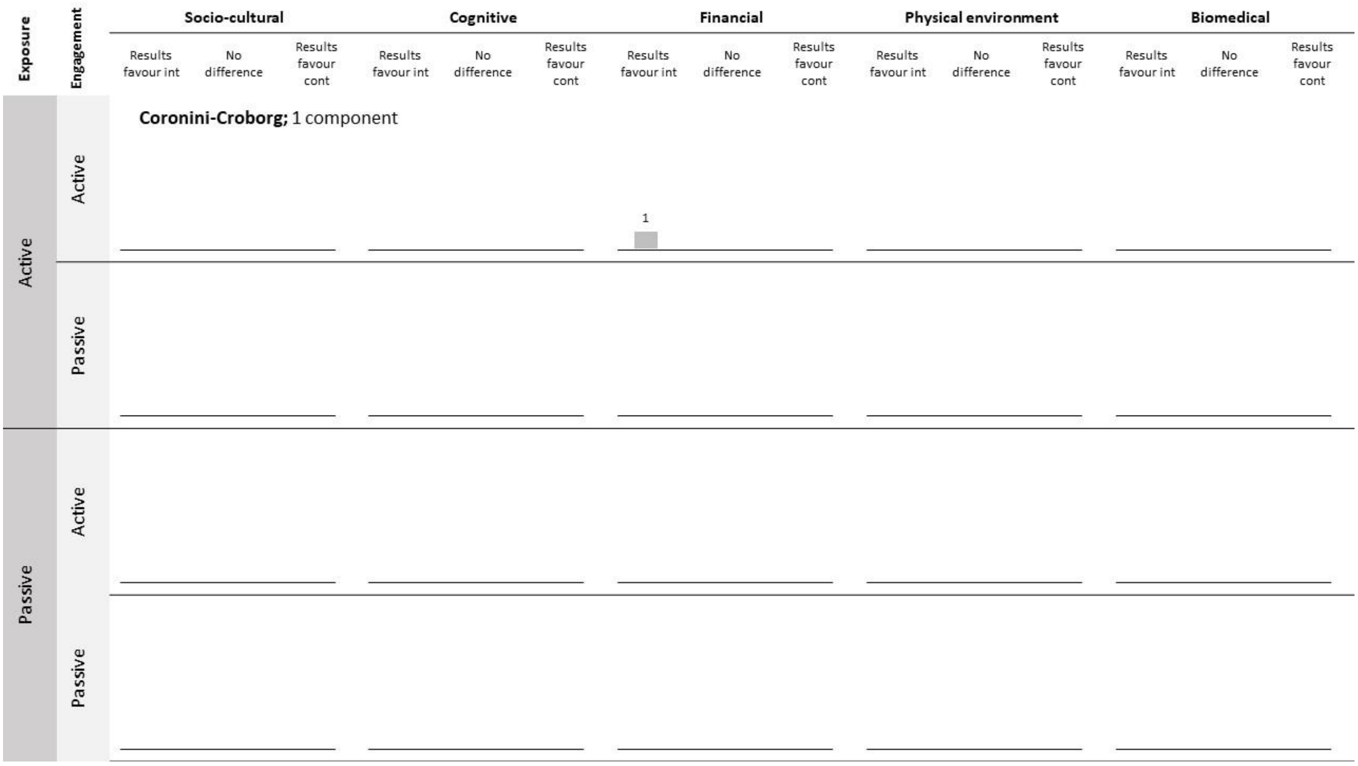


Coronini-Cronberg S, Millett C, Laverty AA, Webb E. The impact of a free older persons' bus pass on active travel and regular walking in England. American Journal of Public Health. 2012;102(11):2141-8.

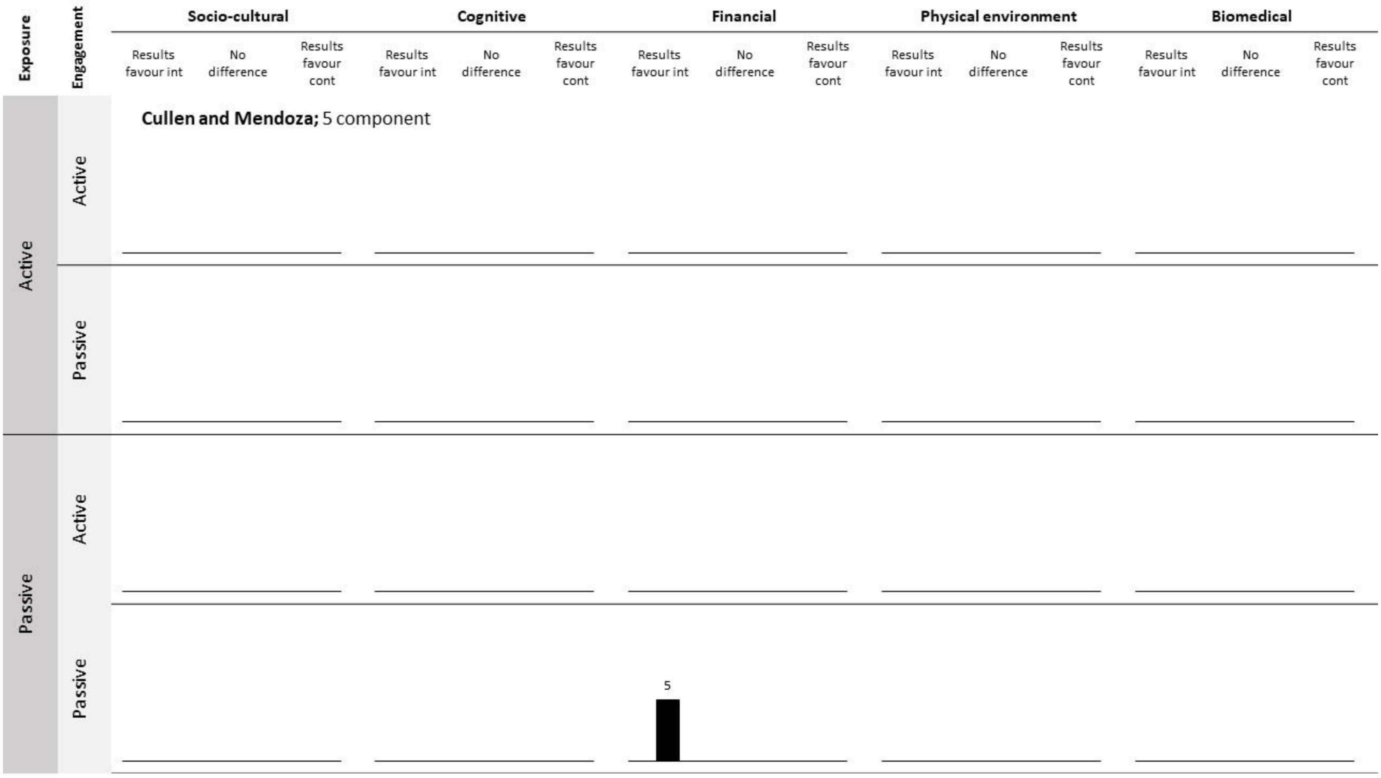


Cullen KW, Watson K, Zakeri I. Improvements in middle school student dietary intake after implementation of the Texas Public School Nutrition Policy. American Journal of Public Health. 2008;98(1):111-7.
Cullen KW, Watson KB, Fithian AR. The impact of school socioeconomic status on student lunch consumption after implementation of the Texas Public School Nutrition Policy. Journal of School Health. 2009;79(11):525-31.
Mendoza JA, Watson K, Cullen KW. Change in dietary energy density after implementation of the Texas Public School Nutrition Policy. Journal of the American Dietetic Association. 2010;110(3):434-40.

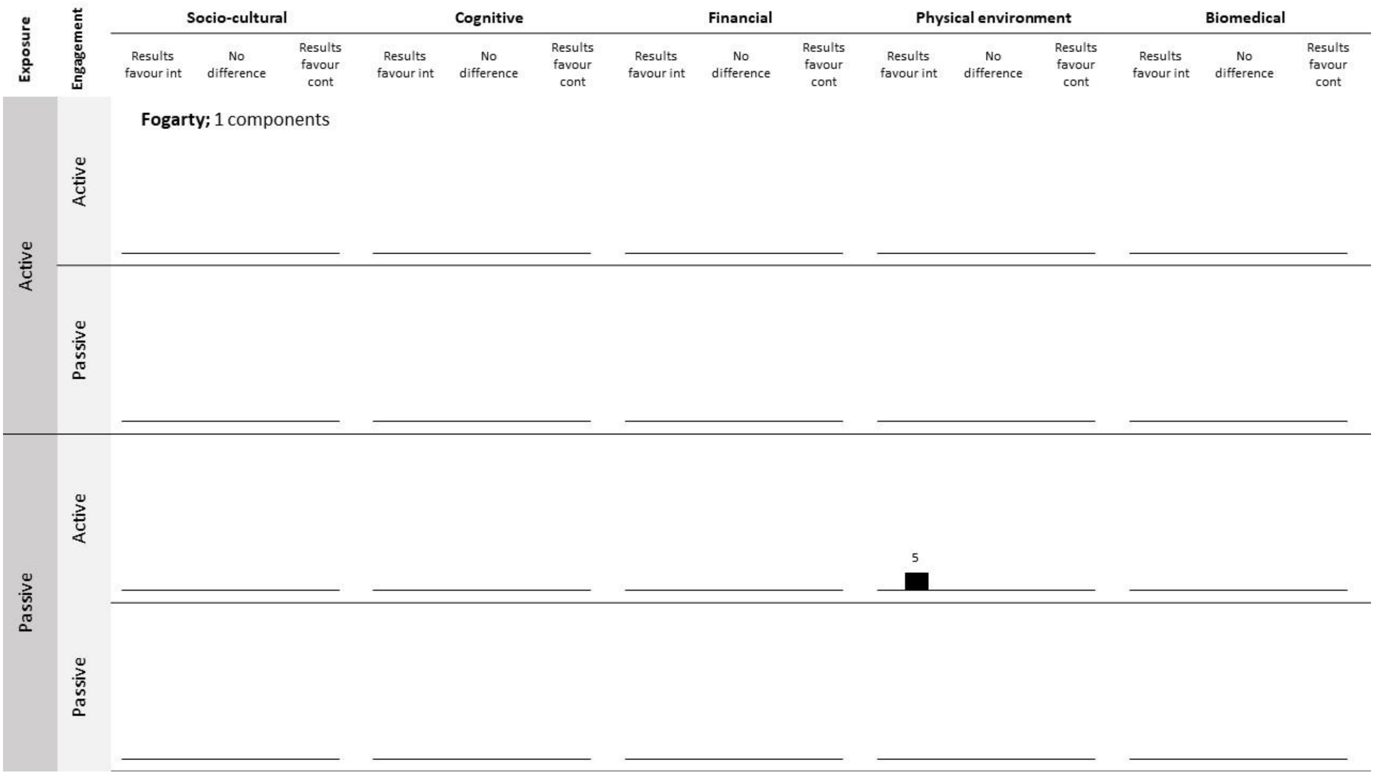


Fogarty A, Antoniak M, Venn A, Davies L, Goodwin A, Salfield N, et al. Does participation in a population-based dietary intervention scheme have a lasting impact on fruit intake in young children? International Journal of Epidemiology. 2007;36(5):1080-5.

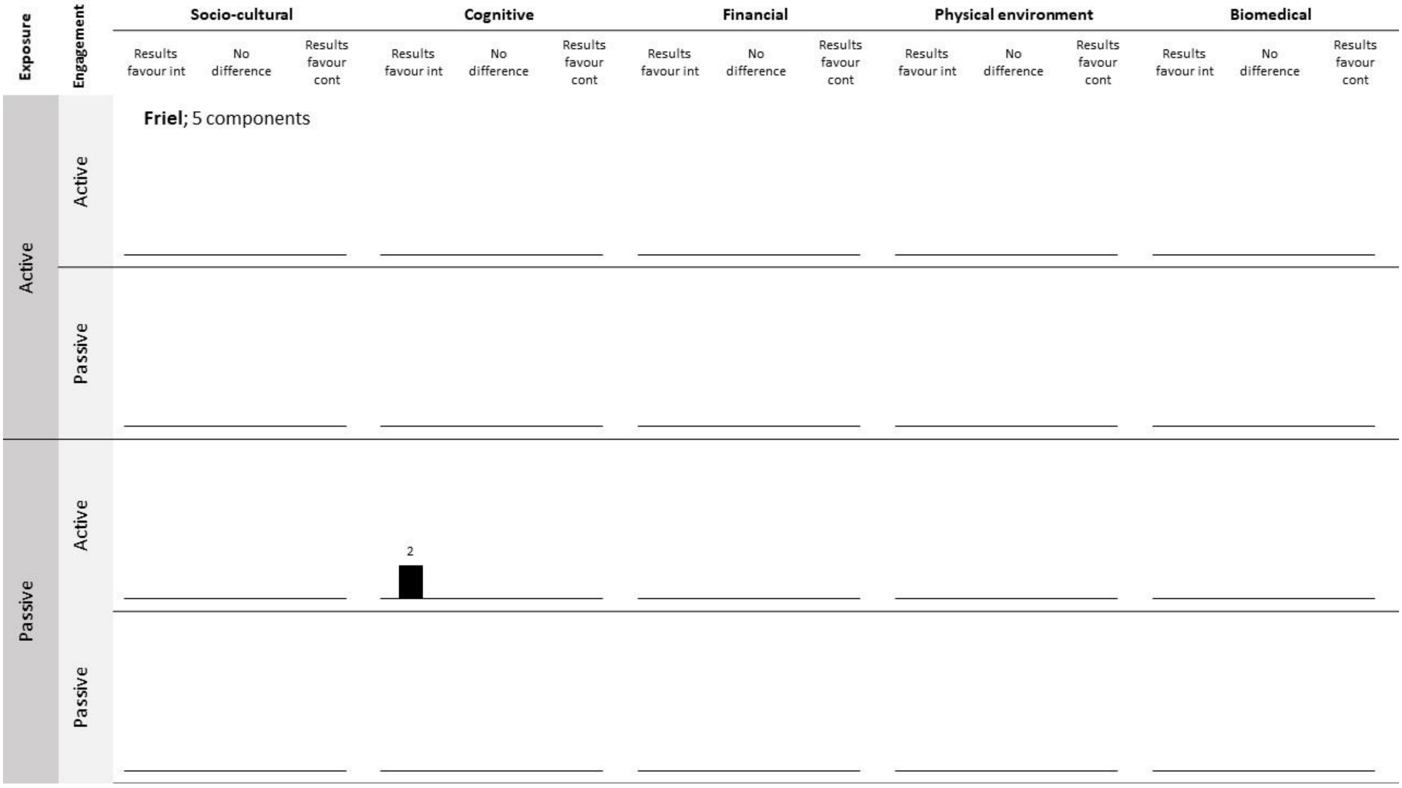


Friel S, Kelleher C, Campbell P, Nolan G. Evaluation of the nutrition education at primary school (NEAPS) programme. Public health nutrition. 1999;2(4):549-55.

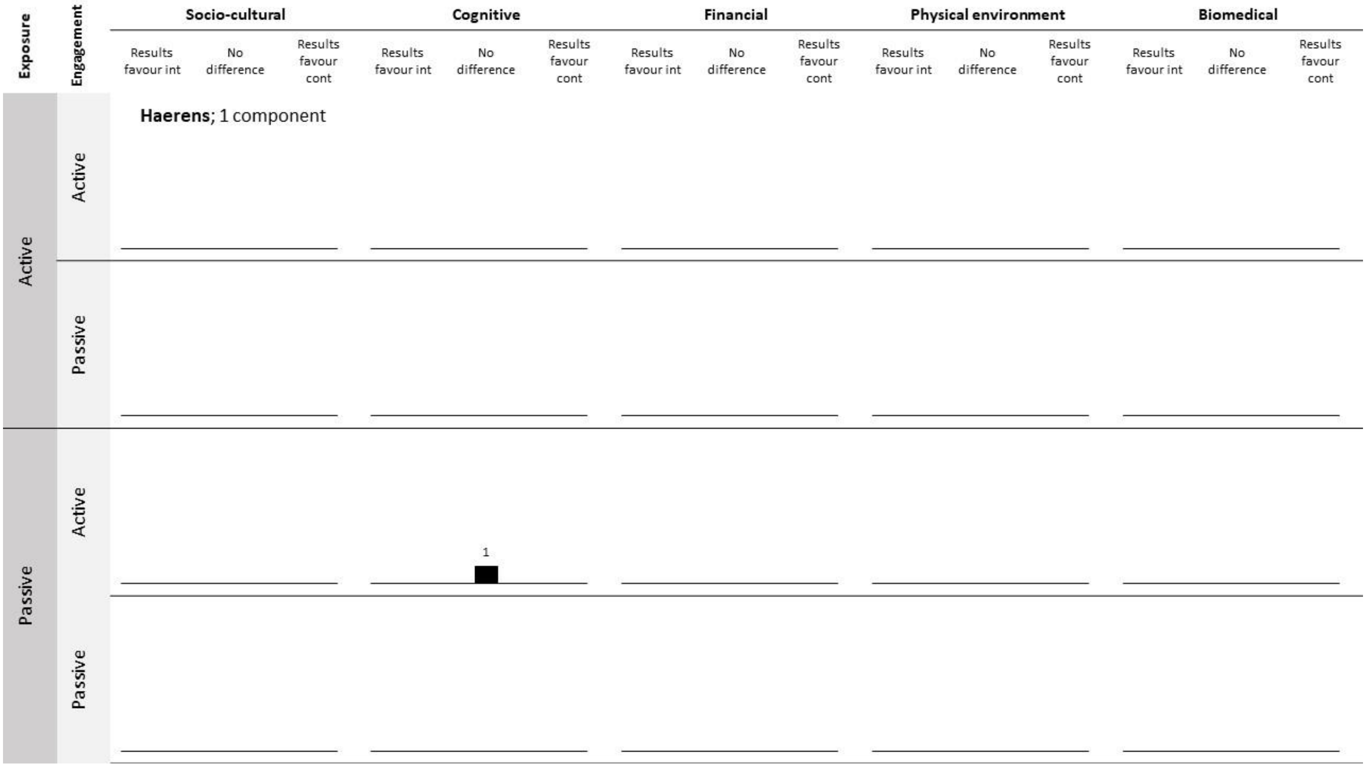


Haerens L, Deforche B, Maes L, Brug J, Vandelanotte C, De Bourdeaudhuij I. A computer-tailored dietary fat intake intervention for adolescents: results of a randomized controlled trial. Annals of Behavioral Medicine. 2007;34(3):253-62.

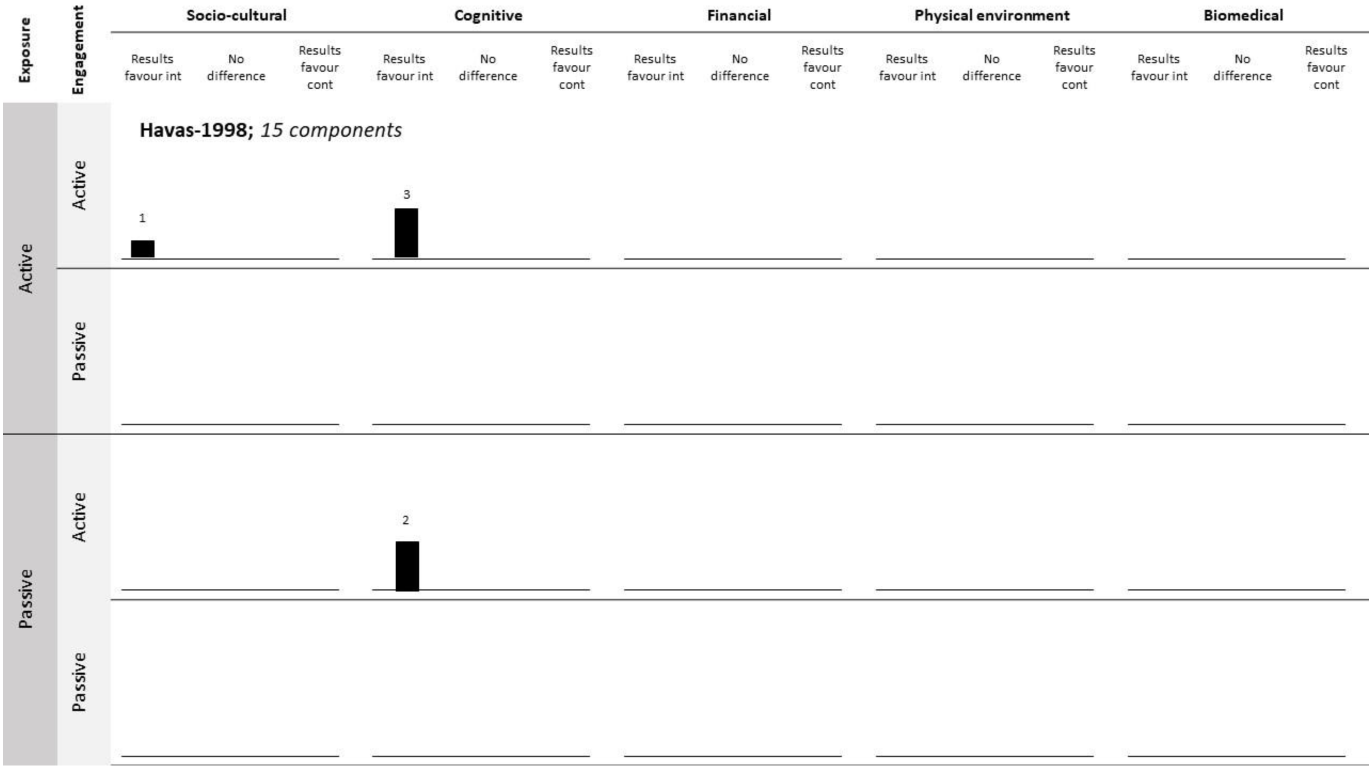


Havas S, Anliker J, Damron D, Langenberg P, Ballesteros M, Feldman R. Final results of the Maryland WIC 5-a-day promotion program. American journal of public health. 1998;88(8):1161-7.

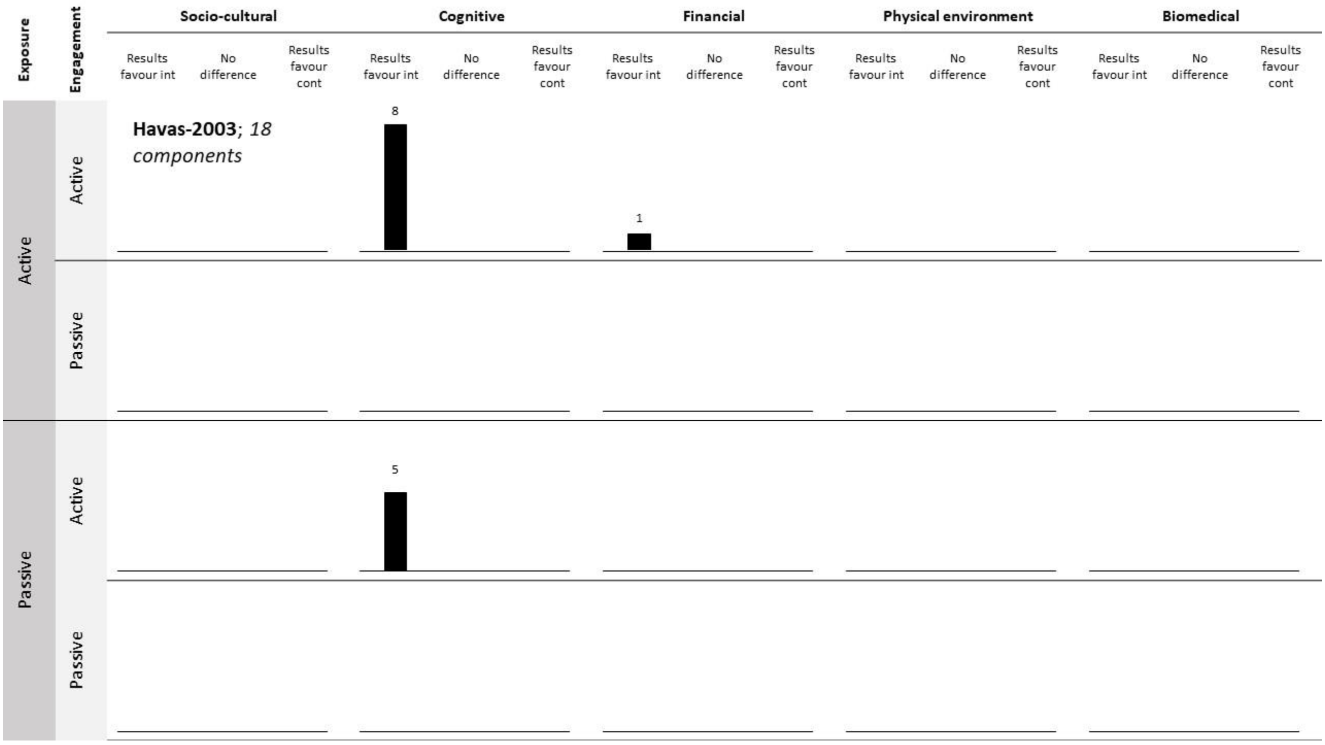


Havas S, Anliker J, Greenberg D, Block G, Block T, Blik C, et al. Final results of the Maryland WIC food for life program. Preventive Medicine. 2003;37(5):406-16.

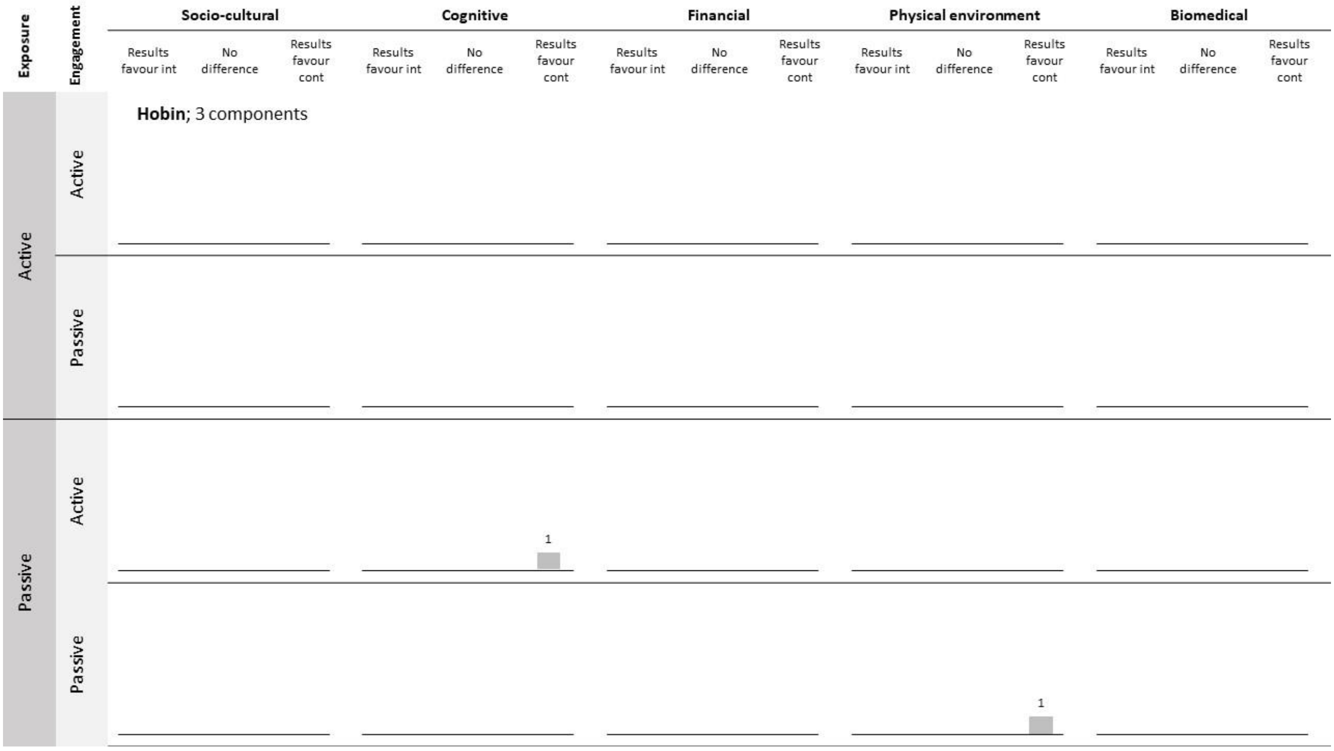


Hobin E, So J, Rosella L, Comte M, Manske S, McGavock J. Trajectories of objectively measured physical activity among secondary students in Canada in the context of a province-wide physical education policy: a longitudinal analysis. Journal of Obesity. 2014;2014.

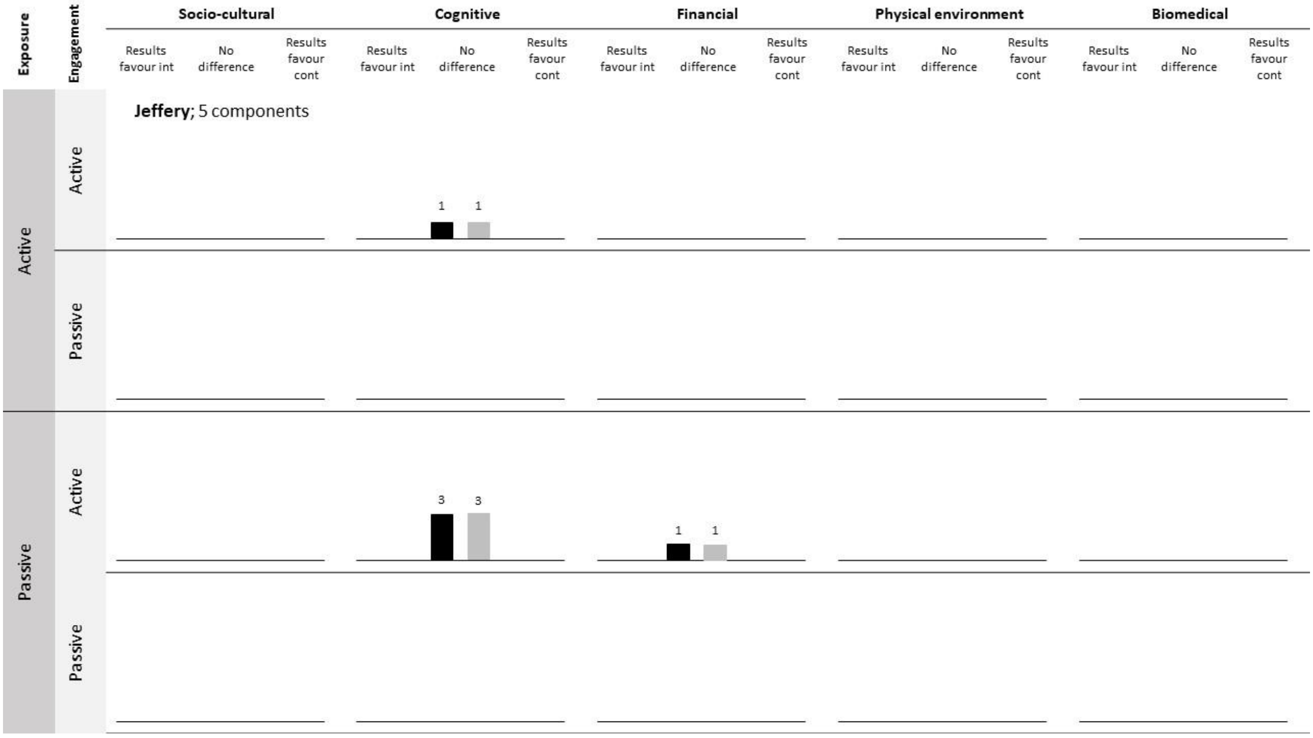


Jeffery R, French S. Preventing weight gain in adults: design, methods and one year results from the Pound of Prevention study. International Journal of Obesity. 1997;21(6):457-64.

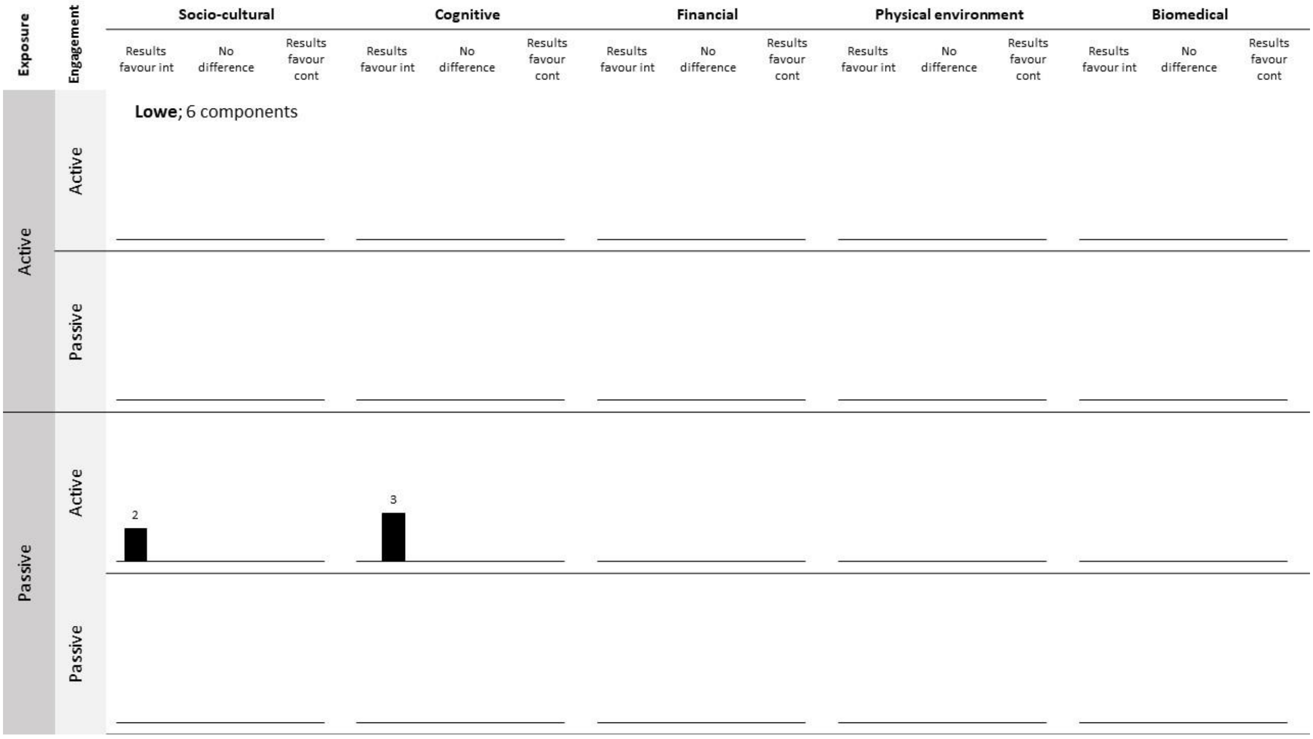


Lowe CF, Horne PJ, Tapper K, Bowdery M, Egerton C. Effects of a peer modelling and rewards-based intervention to increase fruit and vegetable consumption in children. European journal of clinical nutrition. 2004;58(3):510-22.

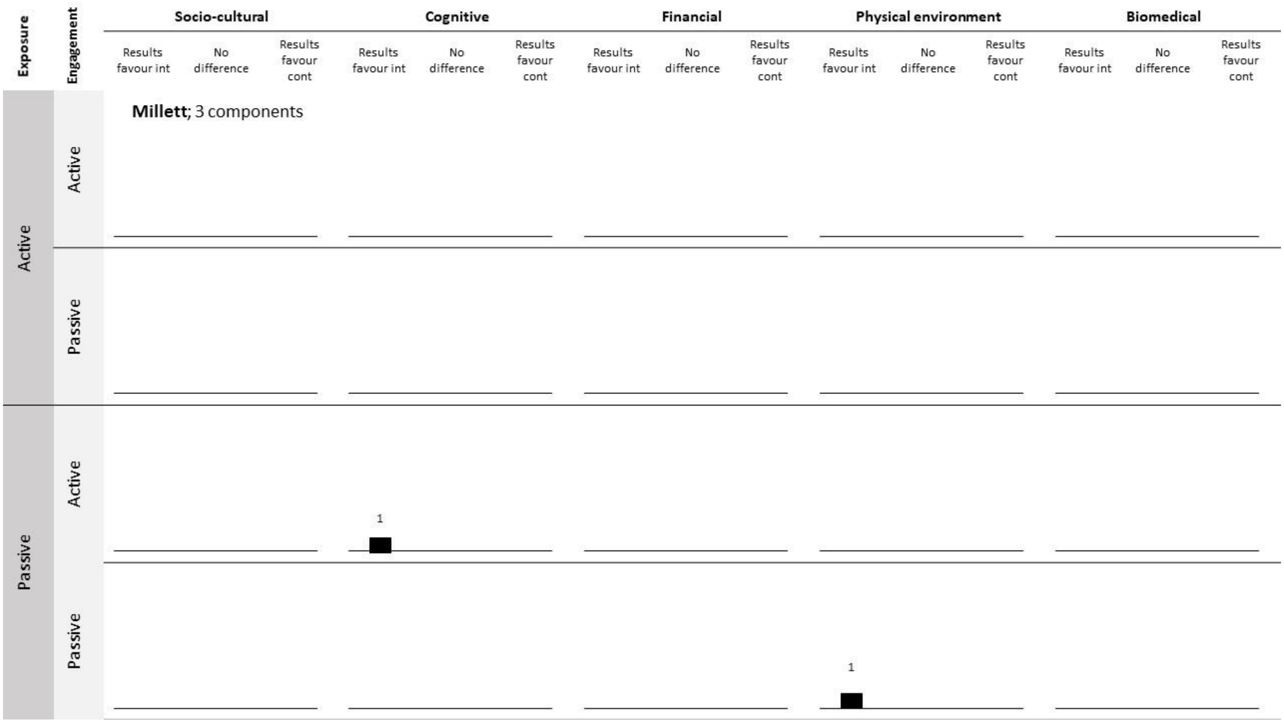


Millett C, Laverty AA, Stylianou N, Bibbins-Domingo K, Pape UJ. Impacts of a national strategy to reduce population salt intake in England: serial cross sectional study. PLoS One. 2012;7(1):e29836.

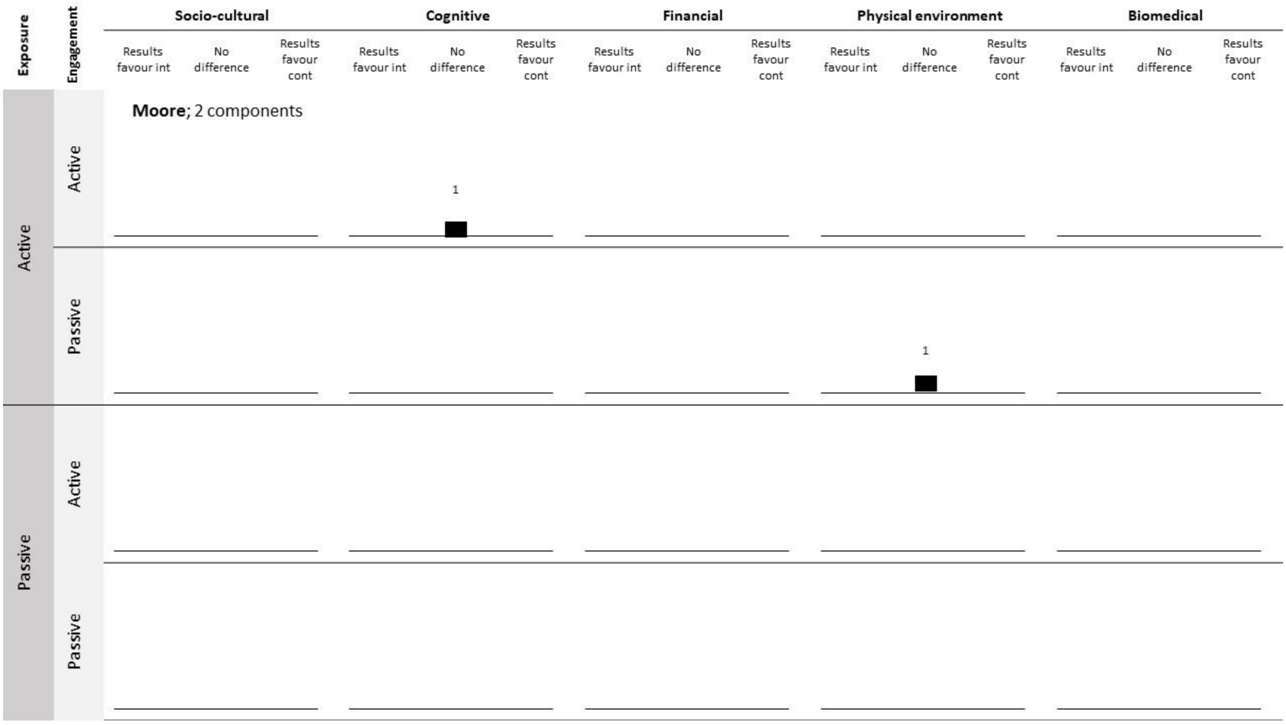


Moore GF, Murphy S, Chaplin K, Lyons RA, Atkinson M, Moore L. Impacts of the Primary School Free Breakfast Initiative on socio-economic inequalities in breakfast consumption among 9–11-year-old schoolchildren in Wales. Public health nutrition. 2014;17(6):1280-9.

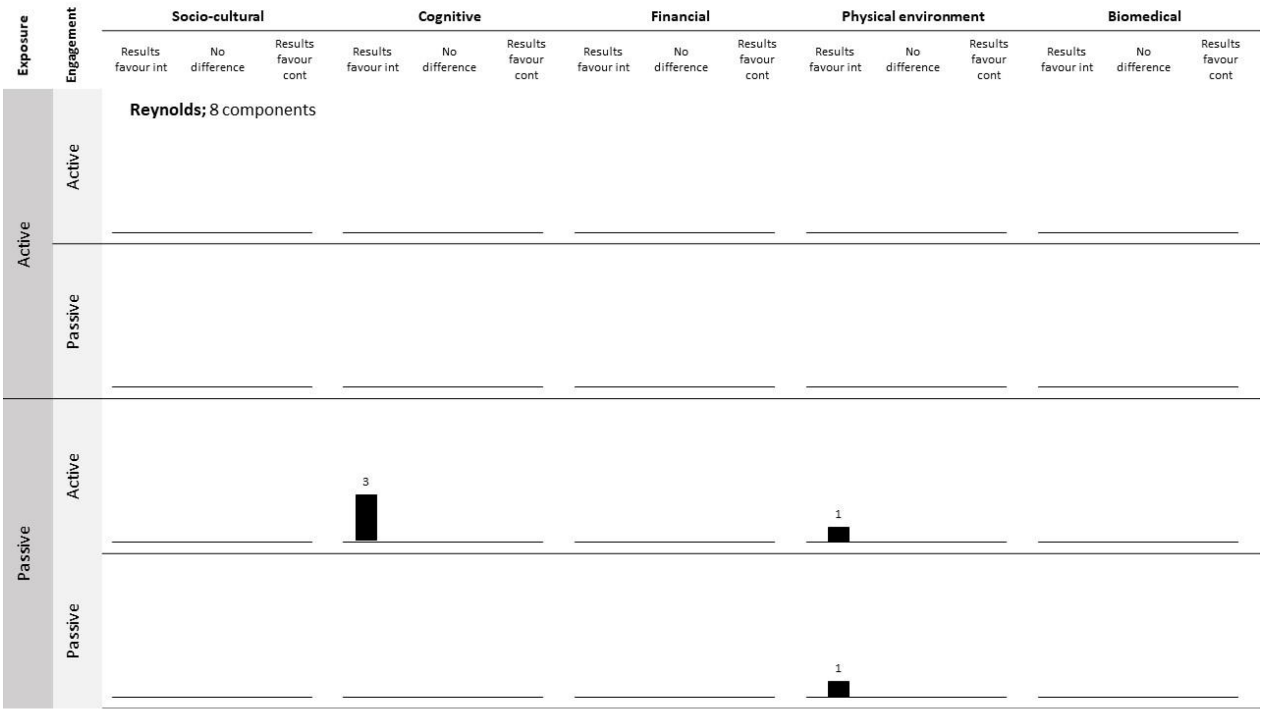


Reynolds KD, Franklin FA, Binkley D, Raczynski JM, Harrington KF, Kirk KA, et al. Increasing the fruit and vegetable consumption of fourth-graders: results from the high 5 project. Preventive medicine. 2000;30(4):309-19.

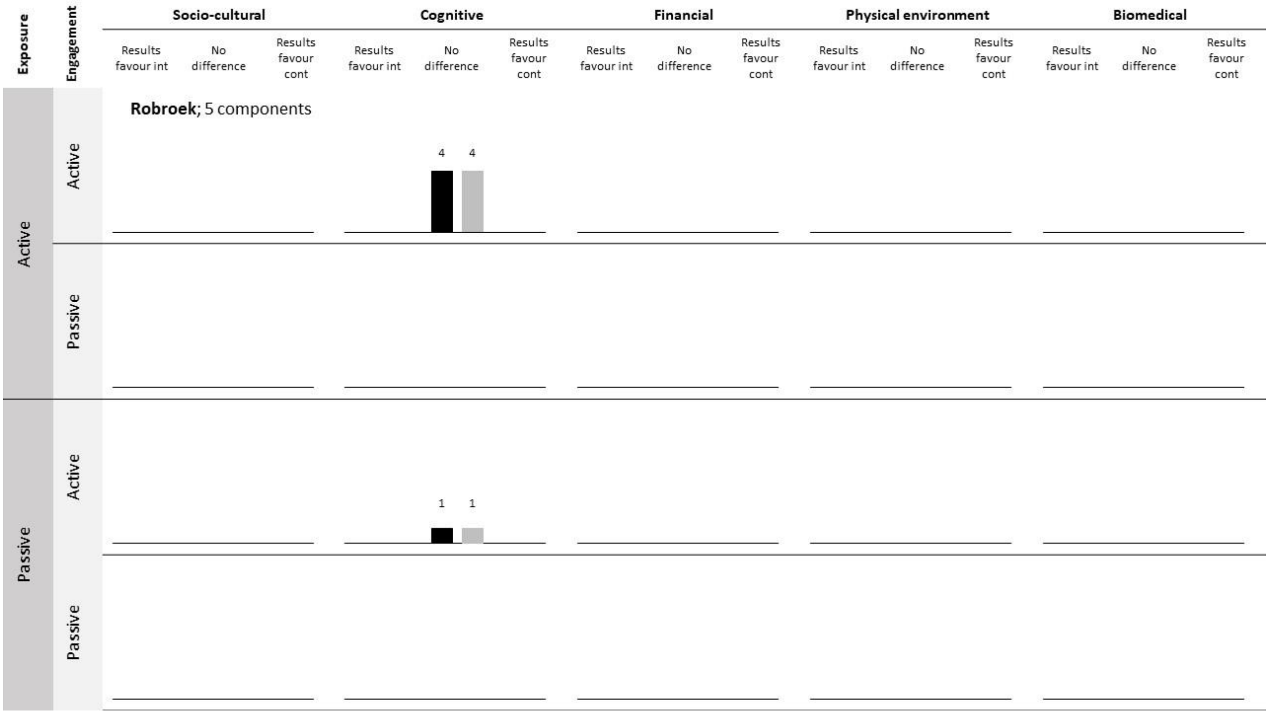


Robroek SJ, Polinder S, Bredt FJ, Burdorf A. Cost-effectiveness of a long-term Internet-delivered worksite health promotion programme on physical activity and nutrition: a cluster randomized controlled trial. Health education research. 2012;27(3):399-410.

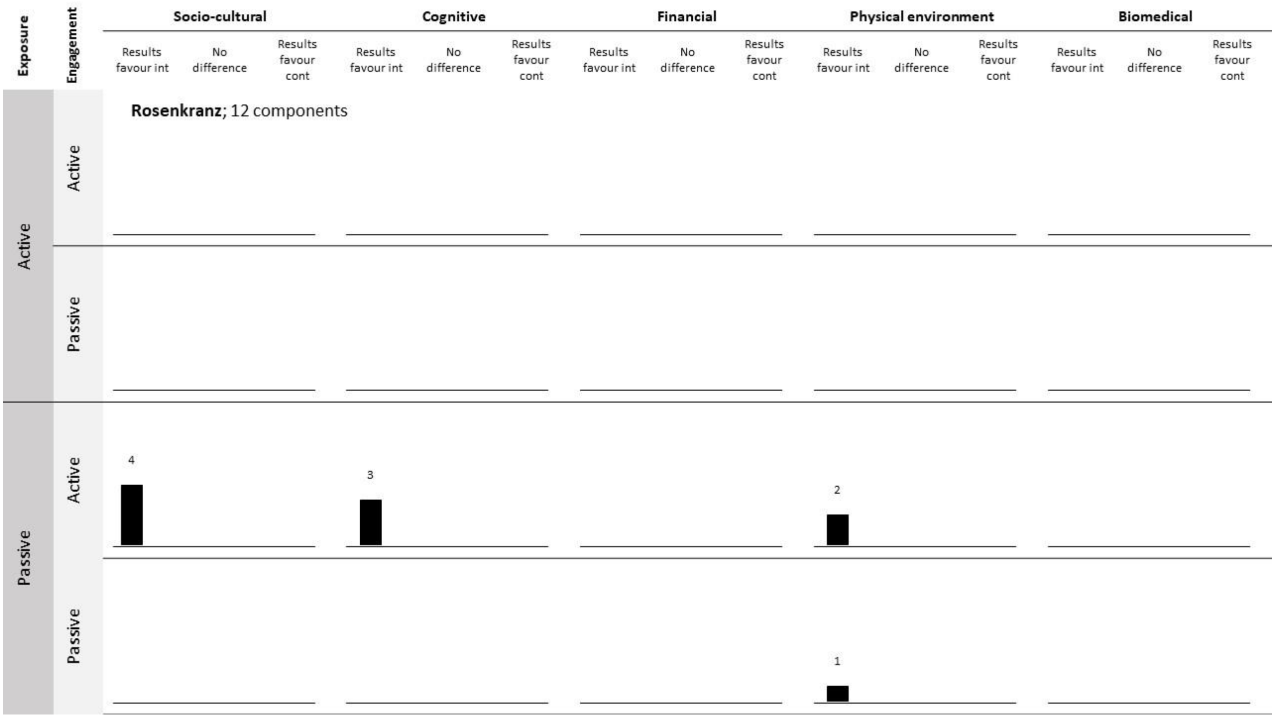


Rosenkranz RR, Behrens TK, Dzewaltowski DA. A group-randomized controlled trial for health promotion in Girl Scouts: healthier troops in a SNAP (Scouting Nutrition & Activity Program). BMC public health. 2010;10:1-13.

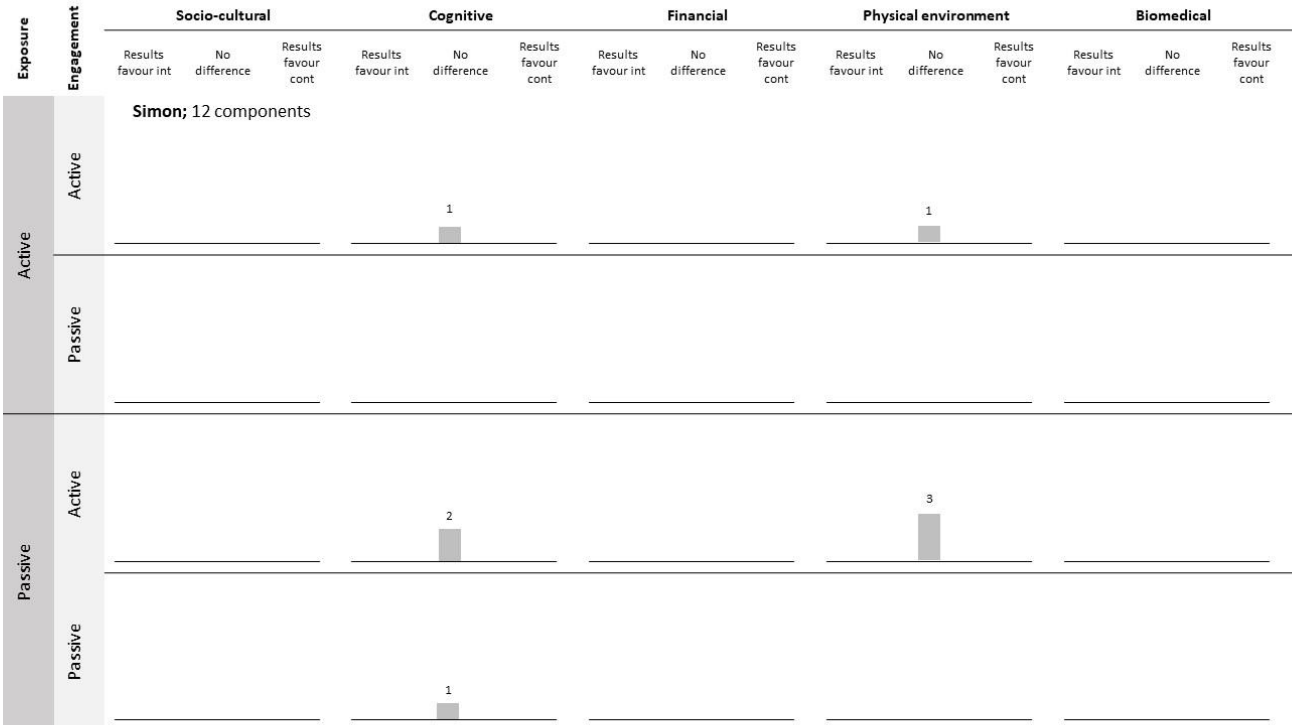


Simon C, Schweitzer B, Oujaa M, Wagner A, Arveiler D, Triby E, et al. Successful overweight prevention in adolescents by increasing physical activity: a 4-year randomized controlled intervention. International journal of obesity. 2008;32(10):1489-98.

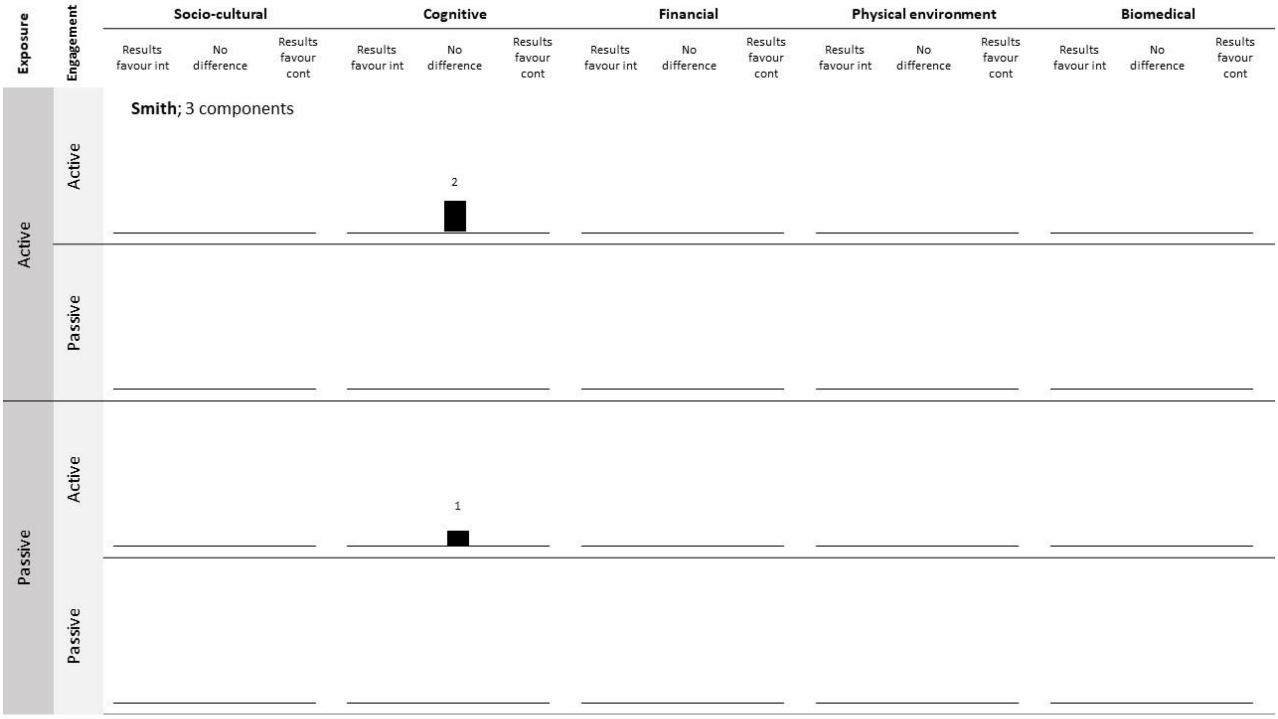


Smith AM, Owen N, Baghurst KI. Influence of socioeconomic status on the effectiveness of dietary counselling in healthy volunteers. Journal of Nutrition Education. 1997;29(1):27-35.

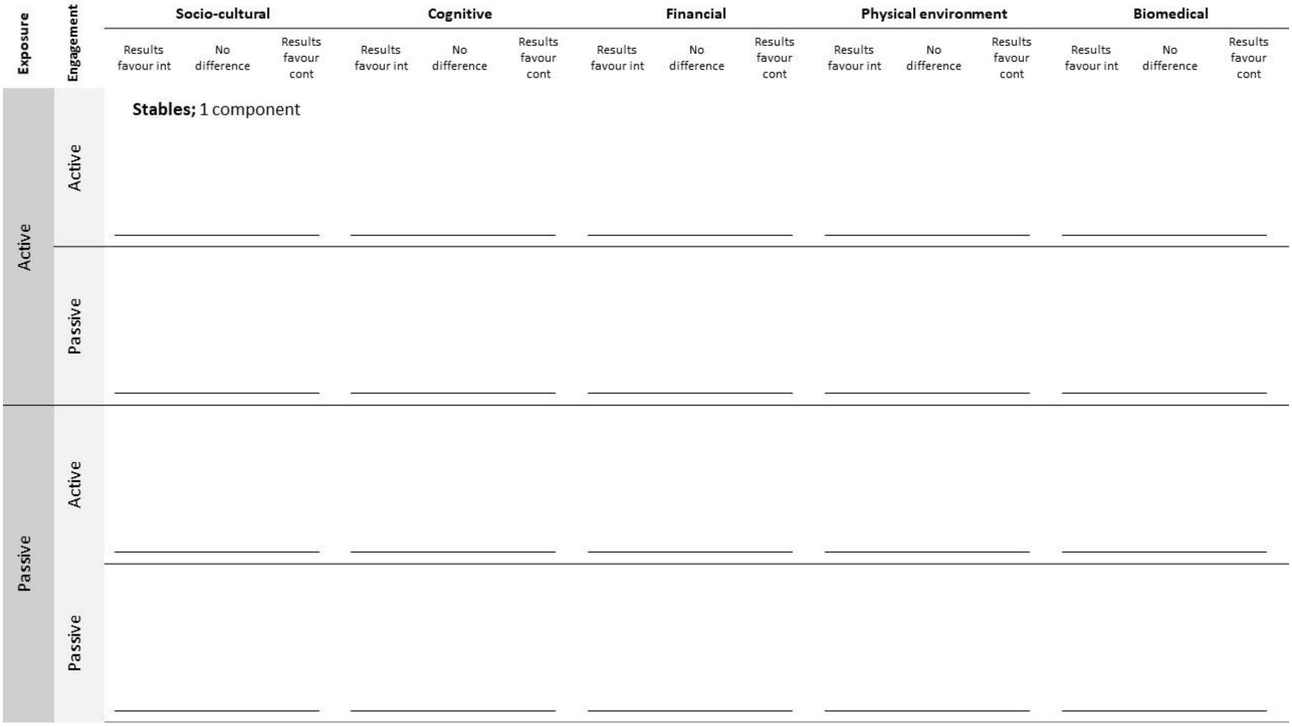


Stables GJ, Subar AF, Patterson BH, Dodd K, Heimendinger J, Van Duyn MAS, et al. Changes in vegetable and fruit consumption and awareness among US adults: results of the 1991 and 1997 5 A Day for Better Health Program surveys. Journal of the American Dietetic Association. 2002;102(6):809-17.

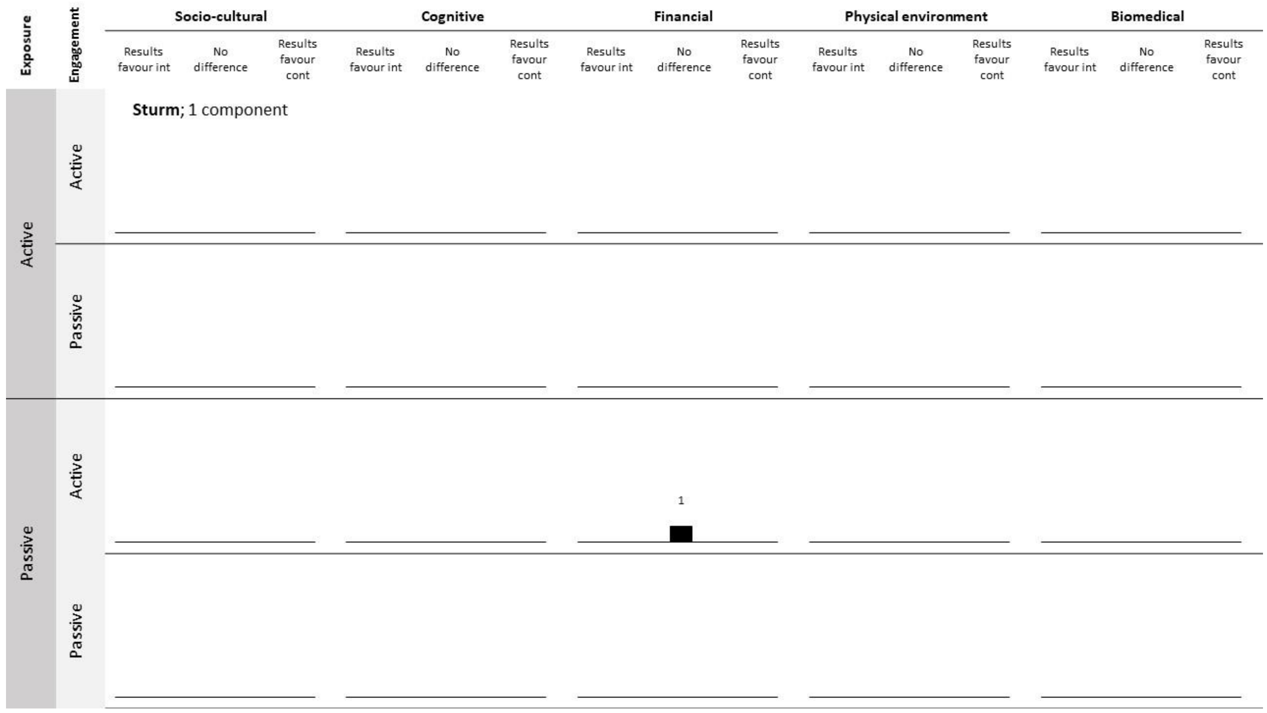


Sturm R, Powell LM, Chriqui JF, Chaloupka FJ. Soda taxes, soft drink consumption, and children's body mass index. Health Affairs. 2010;29(5):1052-8.

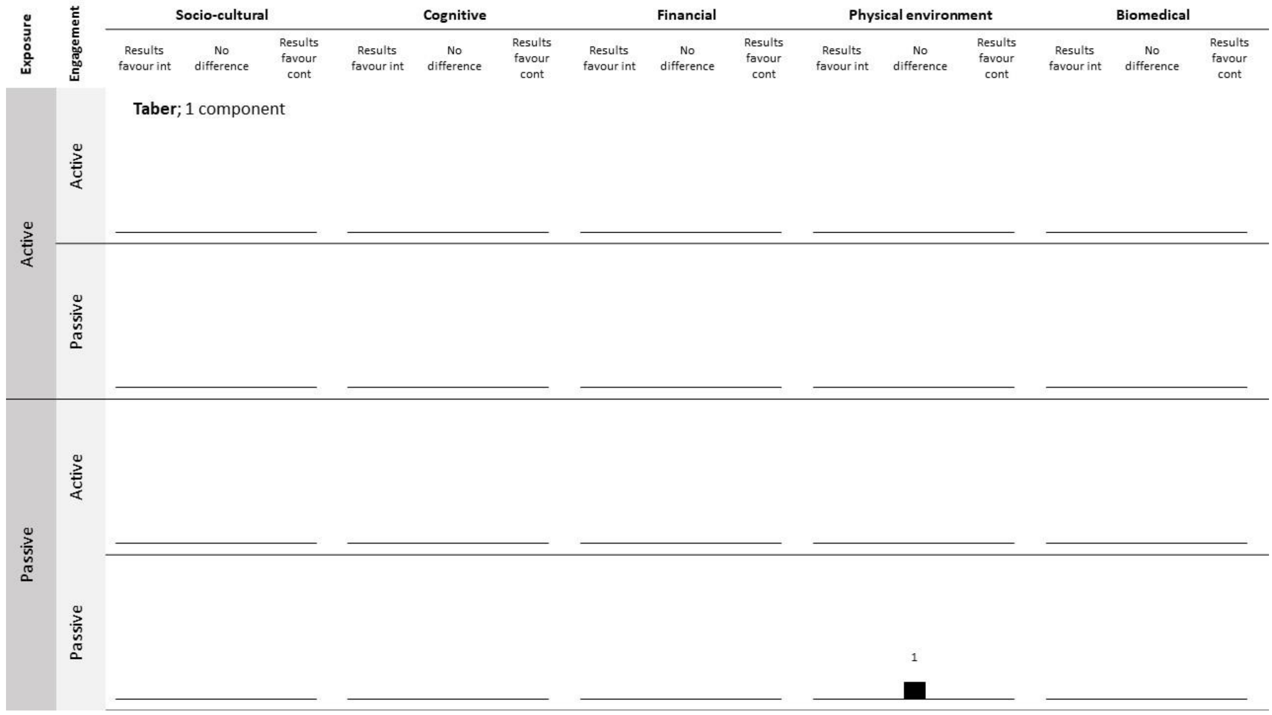


Taber DR, Chriqui JF, Powell L, Chaloupka FJ. Association between state laws governing school meal nutrition content and student weight status: implications for new USDA school meal standards. JAMA pediatrics. 2013;167(6):513-9.

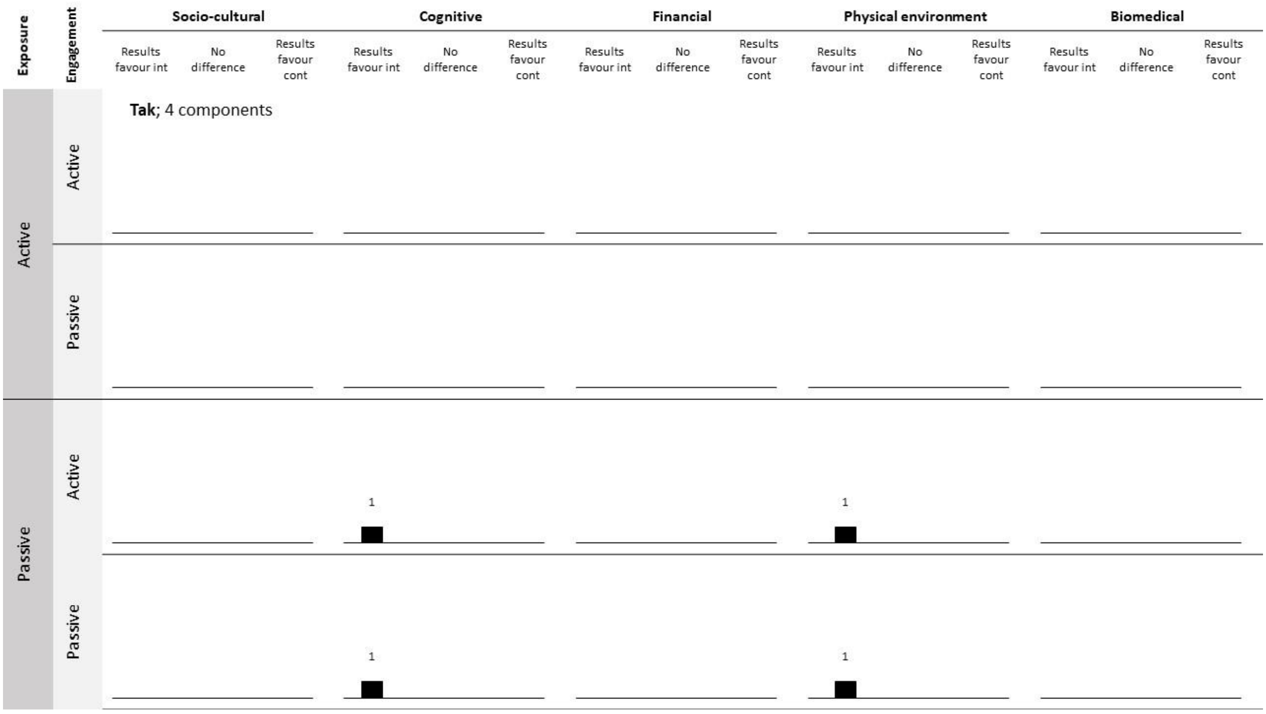


Tak N, Te Velde S, Singh A, Brug J. The effects of a fruit and vegetable promotion intervention on unhealthy snacks during mid-morning school breaks: results of the Dutch Schoolgruiten Project. Journal of human nutrition and dietetics. 2010;23(6):609-15.
Tak NI, Te Velde SJ, Brug J. Long-term effects of the Dutch Schoolgruiten Project–promoting fruit and vegetable consumption among primary-school children. Public health nutrition. 2009;12(8):1213-23.

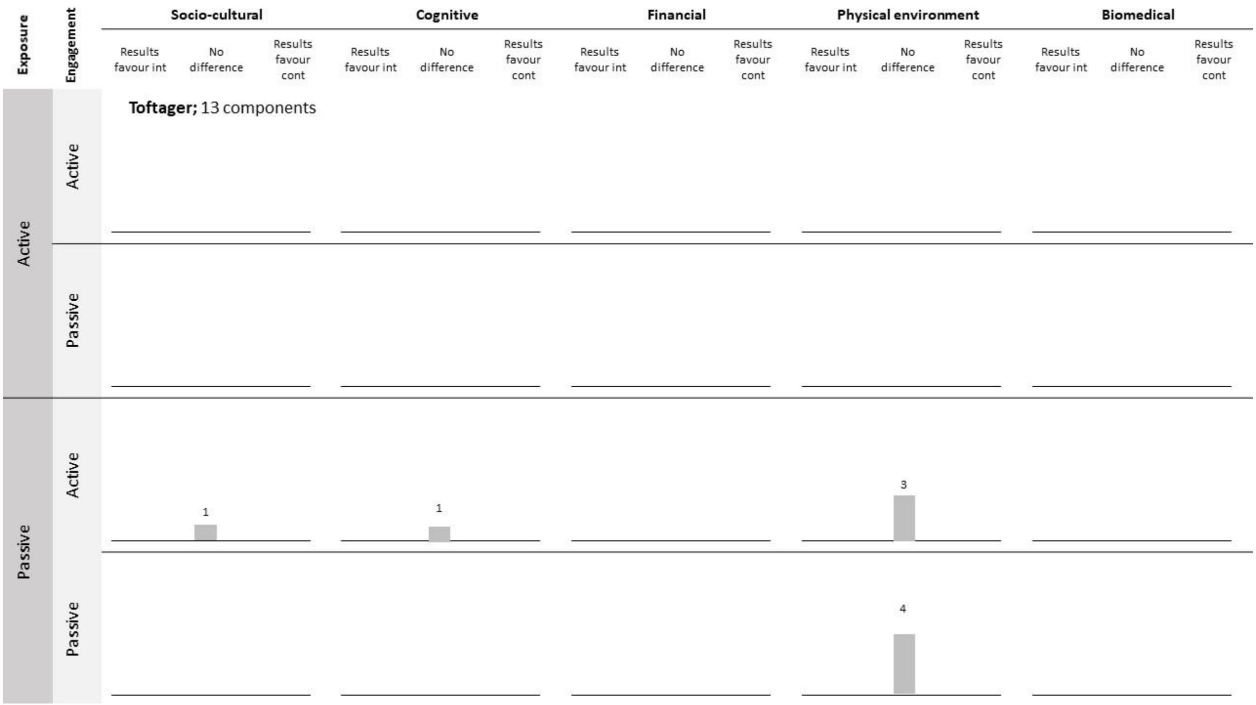


Toftager M, Christiansen LB, Ersbøll AK, Kristensen PL, Due P, Troelsen J. Intervention effects on adolescent physical activity in the multicomponent SPACE study: a cluster randomized controlled trial. PLoS One. 2014;9(6):e99369.
