## Supplementary material 2 for "Development and application of the DePtH framework for categorising the agentic demands of population health interventions"

### Supplementary material 2 – Final DePtH framework

The document provides a detailed description of the final DePtH framework. This includes detailed descriptions and applied examples of the framework constructs. We also provide further details on the principles and rules adopted when applying the framework for the proof-of-concept systematic review in step 3 throughout the document. These reflect our learning and are an evolution from the rules and principles provided to participants in the inter-rater reliability study conducted in step 2b (Supplementary Appendix 2.1). We provide the application rules developed for this study for transparency and do not intend them to be prescriptive. Framework users may wish to develop their own rules and guidance in line with their expertise and project aims.

#### Framework overview

The framework has three application stages, some of which have a number of sub-stages:

1. Identify all relevant intervention component-recipient combinations
    1.1. Identify all relevant intervention components
    1.2. Identify recipient group(s)
    1.3. Cross-tabulate intervention components and recipient groups

2. Identify agentic demand of each component-recipient combination on intervention recipients
    2.1. Categorise exposure of each component-recipient combination
    2.2. Identify the mechanism(s) of action for each component-recipient combination
    2.3. Categorise each mechanism of action
    2.4. Categorise engagement with each mechanism of action

3. Identify other actors involved in each component-recipient combination.

Figure 2.1 shows how application stages 1-3 combine to identify the agentic demands of interventions. Definitions and examples of key terms are provided in Table 2.1 and Supplementary Appendix 2.2 provides a worked example.

**Application rules within proof-of-concept review**

*Phased approach:* We adopted a phased approach within our proof-of-concept systematic review to maximise agreement. We finalised agreement on application stage 1 (identifying all component-recipient combinations) before applying the remaining steps.

*Reviewing application examples:* Prior to data extraction for the proof-of-concept review, all reviewers met and discussed the application of the DePtH framework to intervention examples. This process allowed reviewers to identify potential areas of disagreement and discuss application rules prior to commencing data extraction. During this stage, we also reviewed common reasons for disagreement during the previous online inter-rater reliability.

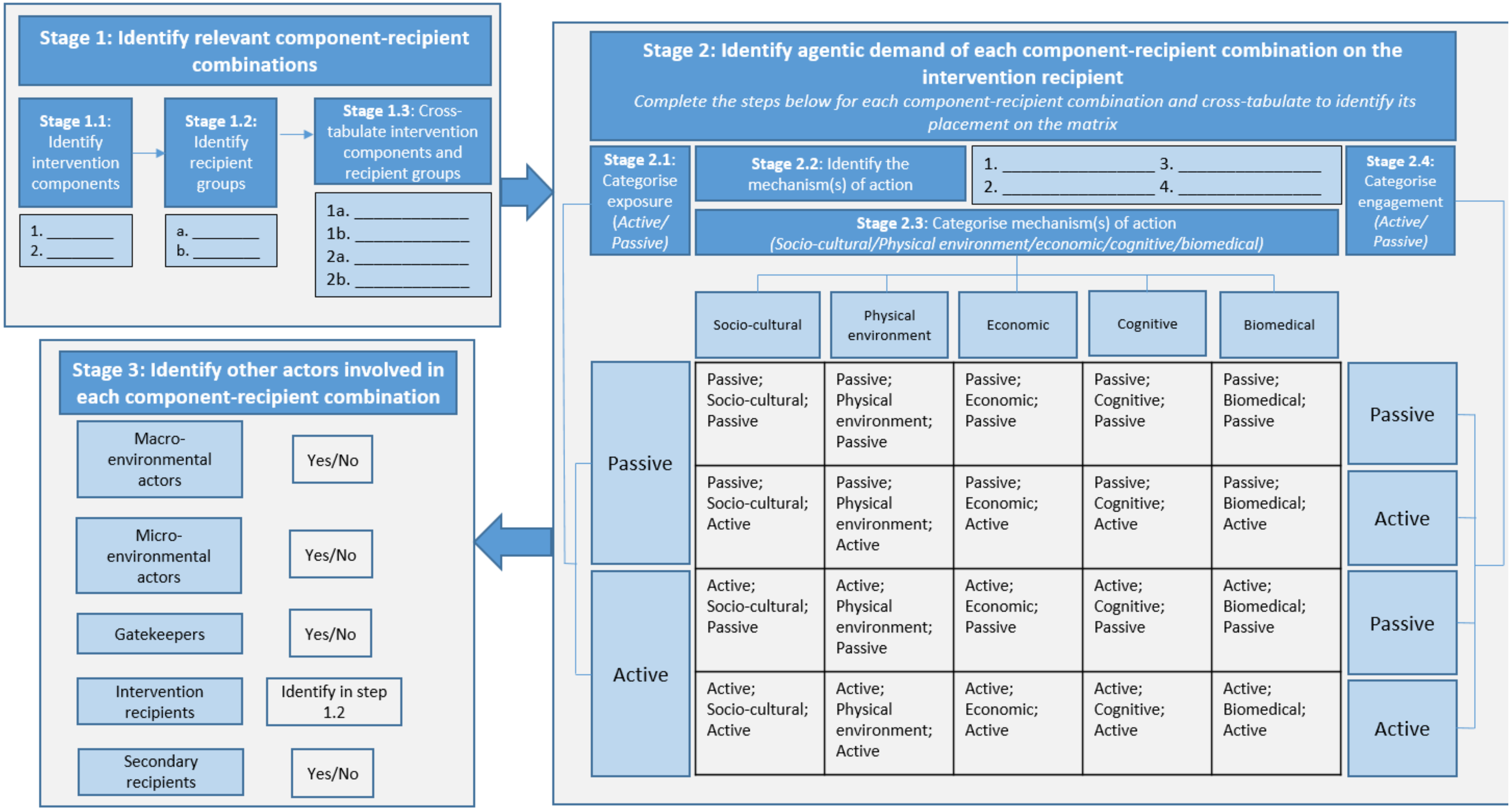


***Figure 2.1** – Outline of stages required to apply the framework to population health interventions*

#### Application Stage 1: Identify all relevant intervention component-recipient combinations

*Application Stage 1.1: Identify all relevant intervention components*

The framework aims to classify the agentic demands of single intervention components and therefore the first step is to identify each intervention component. Each intervention component is likely to have a different form and describes what recipients might 'see' of the intervention. For example, a scheme to increase attendance at a leisure centre may include two components; (1) free access to leisure facilities; and (2) community outreach to engage the local community and let them know about activities at the leisure centre.

**Application rules within proof-of-concept review**

*Phased approach:* Within Application Stage 1, we adopted a phased approach. We first independently identified in duplicate intervention components for 50% of the included studies and identified disagreements. We resolved disagreements via discussion (Phase 1). Following the discussion, we extracted intervention components for the remaining 50% of included studies (Phase 2).

*Greatest granularity:* Although we instructed participants in the online inter-rater reliability study to identify intervention components at the 'highest level', this was interpreted differently by different participants. Therefore, within the proof-of-concept review we revised wording to extract intervention components to the 'greatest granularity'. For example, where an intervention describes 'printed materials including posters, personalised letters and tip sheets' we would identify three intervention components (1) posters; (2) personalised letters; and (3) tip sheets, and classify each according to the DePtH framework.

*Application Stage 1.2: Identify the intended recipient group(s)*

The intended intervention recipients are defined as the individuals whose health is intended to benefit from the intervention as a result of dietary or physical activity behaviour change. In simple cases, intervention recipients will be the target population of an intervention. In others, intervention recipients may be reached via informal gatekeepers e.g. parents provided with guidance on how to limit their children's sugar intake. While the parents may have to change their behaviour, the children will be identified as the intervention recipients because the intention is to improve the health of the children through reducing their sugar intake. In examples like this, framework users should code informal gatekeepers in Application Stage 3. All intervention recipient groups should be identified for each intervention component identified in Application Stage 1.1.

*Application Stage 1.3: Cross-tabulate intervention components and recipient groups*

All relevant combinations of intervention components and recipient groups should be identified by cross-tabulating responses for Application Stages 1.1 and 1.2.

#### Application Stage 2: Identify agentic demand of each component-recipient combination on intervention recipients

Application Stage 2 involves identifying the agentic demand of the intervention component on the intended recipients and involves determining each of exposure, mechanism of action and engagement. Application Stage 2 should be completed for each component-recipient combination identified in Application Stage 1.3.

*Application Stage 2.1: Categorise exposure of each component-recipient combination*

For each component-recipient combination identified in step 1, framework users should determine the type of exposure. Exposure describes how the recipient group first come in to contact with the intervention component. Framework users should distinguish between active and passive exposure.

Active exposure occurs when recipients must change their existing daily activities or initiate new activities to come into contact with the relevant intervention component. Active exposure typically occurs when interventions aim to attract new users to existing or altered settings.

An example of active exposure is an employer provided lunchtime bike maintenance demonstration in an on-site carpark. Here employees have to actively take time out of their normal lunchtime routine (e.g. of attending the canteen, going for a walk) to attend the demonstration.

Passive exposure occurs when no change from existing daily activities is required of recipients for them to come into contact with the intervention component. Passive exposure typically occurs when interventions aim to alter settings for existing users.

An example of passive exposure is free fruit provided in school classrooms. Here school children do not have to change their existing activities to be exposed to the fruit.

**Application rules within proof-of-concept review**

*Insufficient information:* If an intervention description does not detail the exposure, it may be possible for the reviewer to decide based on logical inference and professional expertise. If it is only possible to think of scenarios where exposure is active but not passive (or vice versa) then code as active (or passive if vice versa). However, if insufficient information is provided and reviewers can identify potential ways the intervention is implemented where exposure is passive and other potential ways where exposure is active, then this should be coded as ‘insufficient information to decide’.

**Table 2.1 –** *Definitions and applied examples of DePtH constructs*

| **Term** | **Definition** | **Examples** |
|---|---|---|
| Intervention component | Describes the form of an intervention and what recipients might 'see' of the intervention. | School based intervention that includes three components (1) removal of sugar sweetened beverages in canteen; (2) Nutrition education within curriculum; (3) Provision of physical activity equipment during break. |
| **Exposure** | *How the recipient group first come into contact with the intervention component.* | |
| Active exposure | Intervention recipients must change their existing daily activities or initiate new activities to come into contact with the intervention component. | New park users engaging with park improvements<br>Sign up to financial subsidies for farmers markets<br>Extra-curricular activity encouraging students to log on to educational website |
| Passive exposure | Intervention recipients are not required to change their existing daily activities to come into contact with the intervention component. | Free bus pass delivered directly via mail to recipients<br>Provision of fruit at school break times<br>Menu calorie labelling at fast food restaurants that recipients frequently visit |
| **Intervention mechanism of action** | *How the intervention aims to enable the intended behaviour or discourage an alternative behaviour in order to improve health, ie, the function of the intervention.* | |
| Socio-cultural | Intervention components that aim to change a community or society's attitudes, beliefs, norms and values related to the intended behaviour. | Advertising campaign with celebrity endorsement for fruit and vegetables. |
| Biomedical | Intervention components involving drug or medical techniques that aim to alter the intended behaviour or biological system. | Fluoridation of tap water supply |
| Cognitive | Intervention components that aim to change individual knowledge, attitudes, beliefs or skills concerning the intended behaviour. | Cooking demonstration classes to provide knowledge and skills to enable home cooking. |
| Physical environmental | Intervention components that aim to change the availability, accessibility, safety, placement or properties of infrastructure, facilities, objects or stimuli in the wider environment, including the digital environment. These include intervention components that aim to increase the accesisbililty of intended behaviours. | Provision of a new supermarket in a food desert.<br>Improvements to facilities and pathways in parks. |
| Financial | Intervention components that aim to change the relative monetary cost of intended behaviours. This includes | Provision of bicycles to employees at workplaces to facilitate cycling to work. |

| | | |
|---|---|---|
| | reducing the monetary cost of engaging in the desired behaviour or increasing the monetary cost of alternative behaviours. The provision of free or reduced price tangible goods are also included here. | |
| **Engagement** | *The degree to which recipients are required to be aware of or interact with, the intervention component's mechanism of action in order to benefit as intended.* | |
| Active engagement | Intervention recipients are required to be aware of the intervention mechanism of action and have purposive interaction with it in order to benefit. | Provision of new cycle lane in city<br>Price increase on SSBs resulting from taxation<br>Increase availability of fruit at school lunch time, but keep choice of alternative less healthy desert. |
| Passive engagement | Intervention recipients are not required to be aware of the intervention mechanisms of action (although they may be aware) and will benefit as intended unless they make a purposive action to avoid doing so. | Change number of holes in salt shaker at restaurant to reduce added salt<br>Change PE lesson structure to increase time spent in MVPA<br>Ban sale of HFSS foods in school vending machines<br>Reduce portion size of an existing dish in canteen<br>Reformulate ready meals to reduce salt content |

*Application Stage 2.2: Identify the mechanism(s) of action for each component-recipient combination*

For each component-recipient combination identified in Application Stage 1.3, framework users should identify the relevant mechanism(s) of action. Mechanism of action describes how the intervention component aims to enable the intended behaviour or discourage an alternative behaviour in order to improve health. It is the 'function' that the intervention component performs or 'the causal link between an intervention component and intended outcomes'.[1] Intervention mechanisms of action may occur at the individual level or within the wider system and both should be considered. Some intervention components may have more than one mechanism of action. For example, new off-road cycle infrastructure may be intended to increase perceptions of safety and increase connectivity. In another example, a restaurant calorie labelling intervention may be intended both to inform consumers and incentivise reformulation of restaurant dishes. The former occurs at the individual level and the latter within the wider system.

**Application rules within proof-of-concept review**

*Explicit Mechanisms of action:* Within intervention descriptions included in the proof-of-concept review, we coded only mechanisms of action that are described in the intervention description. It is possible that these are detailed in the introduction, methods, results or discussion section of the paper. At times, we noted that a mechanism of action could be inferred, but that were not explicitly mentioned and following our application rules, we did not code these. Others may wish to apply this rule differently depending on the aims of the review.

*Application Stage 2.3: Categorise the mechanism of action for each component-recipient combination*

Framework users should categorise each mechanism of action identified in the previous stage (Application Stage 2.2) into one of the five categories, adapted from the ANGELO framework.[2] These are socio-cultural, biomedical, cognitive, physical environmental and financial and are described in Table 2.1.

*Application Stage 2.4: Categorise engagement with each mechanism of action*

For each mechanism of action identified in step 2.2 and categorised in step 2.3, framework users should categorise the engagement required with the mechanism of action. Engagement describes the degree to which recipients are required to be aware of, or interact with, the intervention's mechanism of action in order to benefit from it as intended. Framework users should distinguish between active and passive engagement for each mechanism identified in the previous step.

Active engagement occurs when recipients must be aware of the intervention's mechanism of action and have purposive interaction with it in order to benefit as intended.

For example, in order to benefit from the reduced price bike (financial mechanism) provided in a tax-free bike scheme, recipients must purposively interact with the intervention in order to purchase the bicycle and regularly use the bicycle to commute. The nature of population health interventions means that most interventions will be categorised as active engagement.

Passive engagement occurs when recipients do not have to be aware of the intervention's mechanism of action in order to benefit as intended. It is possible for recipients to actively engage with the intervention and they may still benefit if they act in line with the intention of the mechanism of action. Alternatively, recipients may purposively engage to avoid the intentions of the mechanism of action.

For example, an intervention component that reduces the portion size of restaurant food operates through the mechanism of reducing the availability of calories in the serving (physical environmental mechanism). Customers do not have to be aware of the reduction in availability of calories in order to benefit from the intervention. If they become aware of the intervention component, they can continue to order as planned and they will still benefit. However, if they become aware and choose to act to avoid the benefit by deliberately selecting a larger portion size or ordering additional food, they will not benefit as intended.

Results from Application Stages 2.1 (exposure), 2.2 & 2.3 (mechanism of action) and 2.4 (engagement) can be used to categorise the agentic demands on recipients into the matrix in Figure 2.1.

Application Stage 2 should be repeated for all relevant component-recipient combinations identified in Application Stage 1.3.

**Application rules within proof-of-concept review**

*Insufficient information:* An intervention description may not always describe an intervention in sufficient detail to categorise engagement as active or passive, yet as in application stage 2.1 (exposure), it may be possible to decide based on logical inference and professional expertise. If it is possible to think of a scenario where the engagement is passive and another where the engagement is active, then code as insufficient information to decide. If you can only think of scenarios where engagement is active but not passive (or vice versa) then code as active (or passive).

**Step 3: Identify other actors involved for the intervention to achieve its intended effect**

Alongside the agentic demand of interventions on intended recipients, interventions may place an agentic demand on other actors to achieve their intended effects. Figure 3.1 lists other possible actors. As with intended recipients, the ability of additional actors to execute the actions required for the intervention to achieve its intended effects will be influenced by structural factors, and the ability of each actor to act in accordance with the intervention aims will be variable. Not all actors described in Figure 2.1 are required in all interventions. For each relevant mechanism identified in Application Stage 2.2, framework users should identify whether each additional class of actor, described below, is required to act (yes/no).

***Macro-environmental actors*** are those acting at organisational level such as in industries, services or supporting infrastructure, which operate at international, national or local level. It is often not possible to identify specific macro-environmental actors, rather it is clear that action was required at this level. For example, in order for a multi-national fast-food chain to reduce the salt content of their food, there must have been action at the organisational level, but the specific involvement of e.g. individual food scientists may be left unspecified. Macro-environmental actors may also reflect multiple micro-environmental settings working together. For example, a group of leisure centres may act together to subsidise costs for certain users.

**Application rules within proof-of-concept review**

Code as present even if they are not mentioned if it is not feasible that the intervention can be delivered without a macro-environmental actor.

***Micro-environmental actors*** are those acting at the level of individual places or naturally occurring groups of places where people gather for specific purposes,[2] e.g. individual supermarkets or parks. Micro-environmental settings are usually geographically distinct and are relatively small. In some micro-environmental settings, the purpose of gathering is primarily related to the behaviour targeted by interventions e.g. gyms (PA) or restaurants (diet). In others the primary purpose for gathering may be unrelated to target behaviour e.g. schools and workplaces.

**Application rules within proof-of-concept review**

Code as present even if they are not mentioned if it is not feasible that the intervention can be delivered without a micro-environmental actor.

***Informal gatekeepers*** some intervention mechanisms of action require action from other individuals linked to intended recipient(s) in a non-professional manner. These informal gatekeepers may be e.g. parents or peers whose behaviour must change in order to achieve the desired behaviour change in recipients. For example, for a walking school bus to increase children’s physical activity, parents must stop driving children to school and instead make use of the walking bus.

**Application rules within proof-of-concept review**

Code as present even if they are not mentioned if it is not feasible that the intervention can be delivered without an informal gatekeeper.

***Secondary recipients*** are individuals who may benefit from intervention 'spillover' effects. For example, an educational campaign to encourage older men to reduce portion sizes of home cooked food may also benefit anyone else living with and sharing home cooked meals with those men. Secondary recipients are rarely made explicit.

| **Application rules within proof-of-concept review** |
| --- |
| Given the difficulty identifying secondary recipients, only code as present if they are explicitly described in the text. |

**Supplementary Appendix 2.1 –** *Application rules provided to participants taking part in the online inter-rater reliability survey (Step 2b)*

1. **Use only information stated or implied in the information provided**

When answering the survey, please only use information that is stated or implied in the intervention description provided. We acknowledge that in some cases, it may be pertinent to use wider topic knowledge, however to maintain consistency between survey participants, please use only information stated or implied in the description.

1. **Apply the framework consistently between different examples**

We acknowledge that some of the categorisations require you to apply binary categories to constructs that inherently lie on a continuum. Please decide how you wish to apply the framework and ensure you apply this consistently for the remainder of the intervention examples.

2. **Assume all interventions seek to change total diet or physical activity**

All interventions have been selected as their overall aim is to change total diet or physical activity. Where an intervention only measures intermediate outcomes, such as change in knowledge or beliefs it should be assumed, and the framework should be applied according to the interventions aims to change total diet or physical activity.

3. **If unsure, please tell us why and what further information you would have required**

Throughout the survey, if you are unable to select a category, please select the appropriate alternative option. The first is that there is insufficient information in the intervention description to allow you to answer and the second is that there is sufficient information to allow you to answer, but you are unsure which answer to code. For each question where you use these responses, please complete the ‘justification’ box and tell us why it was not possible to code and what further information you would have required, either from the framework instructions or definitions or from the intervention description.

#### **Supplementary Appendix 2.2 –** *Worked example of applying the DePtH framework*

The intervention example is a single component intervention in a workplace canteen setting. The workplace canteen implement menu labelling on all dishes. The intervention is implemented in the canteen of a single workplace.

**Application Stage 1: Identify all relevant intervention component-recipient combinations**

***Application Stage 1.1 - Identify all relevant intervention components***

This is a single component intervention and therefore the only component identified in this intervention is adding labels to menus within workplace canteen (Menu labels).

***Application Stage 1.2 - Identify recipient group(s)***

The intended recipient group of this intervention are employees who are regular users of the workplace canteen to purchase lunch (Existing users).

***Application Stage 1.3 - Cross tabulate intervention components and recipient groups***

*Component-recipient combination 1:* Menu labels-Existing users

**Application Stage 2: Identify agentic demand of each component-recipient combination on intervention recipients**

*Component-recipient combination 1:* Menu labels-Existing users

***Application Stage 2.1 - Categorise exposure of each component-recipient combination***

The employees targeted by this intervention are existing users of the workplace canteen and therefore are not required to make a change from this activity in order to come into contact with the menu labelling intervention component. Therefore the exposure of the existing users is categorised as passive. This exposure category is the same for all mechanisms of action identified in the next step.

***Application Stage 2.2 - Identify the mechanism(s) of action for each component-recipient combination***

Menu labelling operates through two mechanisms of action which include;

1) informing intervention recipients of the calorie content of food that they intend to purchase to guide their choice towards lower calorie options. This mechanism aims to change the knowledge of existing canteen users about the calorie content of their goods.
2) trigger the food manufacturers to reformulate their products resulting in lower calorie options being served in the canteen. This mechanism of action aims to change the availability of lower calorie food options.

***Application Stage 2.3 - Categorise the mechanism(s) of action for each component-recipient combination***

1) The mechanism that informs recipients of the calories content is categorised as cognitive because it aims to change their knowledge of the calorie content of food.
2) The food reformulation mechanism is categorised as physical environmental because it aims to change the availability of lower calorie food options in the canteen.

***Application Stage 2.4 - Categorise engagement with each mechanism of action***

This stage must be completed for the two mechanisms of action identified in Application Stage 2.2.

1) In order to benefit from the intervention via the cognitive mechanism, recipients must be aware of the menu label, take notice of it and purposively interact with the label to order a lower calorie dish in order to benefit from the intervention as intended. Therefore, the engagement with the intervention mechanism is categorised as active.
2) In order to benefit from the reformulation of food products, recipients are not required to be aware that the products have been reformulated. They can continue with their existing behaviour without purposeful change and benefit from the reduced calorie content. Therefore the engagement with the intervention mechanism is categorised as passive.

| *2.1 Exposure of component-recipient combination* | *2.2 Identify mechanisms of action* | *2.3 Categorise the mechanisms of action* | *2.4 Categorise engagement with the mechanism of action* |
|---|---|---|---|
| Passive | Inform canteen users of calorie content of food | Cognitive | Active |
| Passive | Reformulation of canteen food in response to labelling requirement | Physical environmental | Passive |

**Step 3: Identify other actors involved in each component-recipient combination**

| | | |
|---|---|---|
| Macro-environmental actors | No | This intervention is applied to a single workplace and therefore there are no macro-environmental actors involved. |
| Micro-environmental actors | Yes | The workplace canteen is a place where people gather for the specific purpose of eating their lunch during working hours and is a geographically distinct setting. |
| Informal gatekeepers | No | There are no informal gatekeepers involved in this intervention. |
| Secondary recipients | No | There are no secondary recipients involved in this intervention as the intervention influences the food purchasing and consumption within the workplace canteen. |
