## Supplementary material 3 for "Development and application of the DePtH framework for categorising the agentic demands of population health interventions"

### Supplementary material 3 – Details of interventions included in testing the DePtH framework

**Table 3.1 -** *Table of study characteristics*

| Study design, aim | Participants/ population | Behavioural target | Intervention summary | Intervention components-recipient combinations (n) | Effectiveness measure and category | SEP Measure and category | Quality rating [a] |
|---|---|---|---|---|---|---|---|
| **Bere-2005 (Norway)(1)** | | | | | | | |
| Design: Repeat cross-sectional with unexposed group<br>Aim: To investigate the effect of free fruit provision at school on the consumption of fruit and vegetables and 'unhealthy' snacks | 7th grade pupils from 29 schools in Hedmark and Telemark counties in Norway (n=638) | Diet | Pupils receive a piece of fruit or carrot each school day in connection with their lunch meal, provided for free. | Recipients: School children<br>Components (n=1)<br>1.Free school fruit | Fruit and vegetable intake at school<br><br>Results favour intervention | Parent education level<br><br>No preferential impact | Moderate |
| **Bere-2010 (Norway)(2)** | | | | | | | |
| Design: Repeat cross-sectional with unexposed group<br>Aim: Evaluate nationwide implementation of free school fruit on adolescent fruit and vegetable intake | 6th and 7th grade pupils from 17 schools in Hedmark and Telemark counties | Diet | Official free school fruit programme where children are entitled to a free piece of fruit or vegetable every school day | Recipients: School children<br>Components (n=1)<br>1.Free school fruit | Fruit and vegetable intake at school<br><br>Results favour intervention | Parental education level<br><br>No preferential impact | Weak |
| **Brownson-1996 (USA)(3)** | | | | | | | |
| Design: Repeat cross-sectional<br>Aim: Evaluate impact of the Bootheel Heart Health Project on cardiovascular disease risk factors | Representative sample of adults in six counties in southeastern Missouri. Region characterised by high rates of poverty and low education levels | Diet and PA | Six community coalitions receive $5000 per year to select, prioritise and implement community based interventions. | Recipients: Adults in Botheel region<br>Components (n=11)<br>1. Walking clubs<br>2. Aerobic exercise classes<br>3. Community blood pressure, diabetes and cholesterol screening<br>4. Cardiovascular disease education programs<br>5. Heart Healthy Cooking Demonstrations | Prevalence of 5+ serving fruit and vegetable daily<br><br>Diet: No difference<br><br>PA: Results favour intervention | Education level<br><br>Diet: Likely to reduce inequalities | 3 |

| | | | | | | | |
|---|---|---|---|---|---|---|---|
| | | | | 6. Exercise demonstrations<br>7. Heart disease education in sermons<br>8. Heart Healthy Dinners served in church<br>9. Poster contest<br>10. Weekly newspaper column<br>11. Environmental changes, eg, walking path | | PA: No preferential impact | |
| **Campbell-1999 (USA)(4)** | | | | | | | |
| Design: RCT<br>Aim: To assess effectiveness of the intervention to increase fruit and vegetable intake among rural African American Church Members | African American attendees of churches in 10 rural counties in eastern North Carolina (n=2,519) | Diet | Multi-component intervention targeting pre-disposing factors, enabling factors and reinforcing factors | Recipients: Members of church<br>Components (n=18)<br>1. Tailored bulletins<br>2. Printed materials – brochures<br>3. Printed materials – posters<br>4. Printed materials – banners<br>5. Printed materials – bulletin board materials<br>6. Printed materials – idea sheets<br>7. Printed materials – church bulletin inserts<br>8. Gardening<br>9. Educational sessions on cooking<br>10. Cookbook and recipe tasting<br>11. Serving more fruit and vegetables at church functions<br>12. Lay health advisors<br>13. Community coalitions to plan community events<br>14. Pastor support<br>15. Grocer-vendor involvement – farmers market posters<br>16. Grocer-vendor involvement – coupon<br>17. Grocer-vendor involvement – recipe cards<br>18. Church initiated events | Fruit and vegetable consumption<br><br>Results favour intervention | Household income<br><br>No preferential impact | 5 |

| | | | | | | | |
|---|---|---|---|---|---|---|---|
| **Coronini Cronberg-2012 (UK)(5)** | | | | | | | |
| Design: Repeat cross-sectional<br>Aim: To assess the impact of a free older persons' bus pass on active travel and regular walking in England | Adults aged over 60 in England (n=16,911) | PA | Free bus pass for older adults | Recipients: Adults aged 60+<br>Components (n=1)<br>1. Free bus pass | Use of active transport, buses and walking 3 or more times/week<br><br>Results favour intervention | Housing tenure<br><br>No preferential impact | Moderate |
| **Cullen-2009 & Mendoza-2010 (USA)(6-8)** | | | | | | | |
| Design: Repeat cross-sectional<br>Aim: Compare impact of Texas Public School Nutrition Policy on lunch consumption of low and middle income students | Two schools in southeast Texas | Diet | An unfunded mandate to promote healthy school environments restricting portion sizes of snack and high fat foods, sales of sweetened beverages and the fat content of all foods. | Recipients: School children<br>Components (n=5)<br>1.Restrict portion sizes of sugar-sweetened beverages in schools<br>2. Limit frequency and serving size of high fat vegetables<br>3. Restrict portion sizes of snacks and high fat foods<br>4. Restricts sales of sweetened beverages<br>5. Restricted fat content of all foods | Nutrient, food group and energy density consumption<br><br>Results favour intervention | School level free school meals<br><br>No preferential impact | Weak |
| **Fogarty-2007 (UK)(9)** | | | | | | | |
| Design: Repeat cross-sectional study<br>Aim: Determine effect of school fruit provision of fruit intake in young children and any effect after intervention ceased | 4-6 year old children attending 113 local authority maintained schools in East Midlands, England | Diet | National Schools Fruit Scheme provided a daily piece of fruit distributed at school | Recipients: School children<br>Components (n=1)<br>1. Provision of free fruit at school | Fruit consumption (eating every day, pieces p/week)<br><br>Results favour intervention | Townsend Deprivation Score<br><br>No preferential impact | Weak |
| **Friel-1999 (Ireland)(10)** | | | | | | | |
| Design: quasi-RCT<br>Aim: To assess change in children's behaviour resulting from the pilot intervention | Irish schoolchildren aged 8-10 years from 8 schools (n=812) | Diet | Education programme comprising 20 sessions over 10 weeks | Recipients: School children<br>Components (n=5)<br>1. Lesson plans for teachers<br>2. Activity worksheets for pupils<br>3. Home team pack<br>4. Teacher training | Food pairing questionnaire score<br><br>Results favour intervention | Area level deprivation<br><br>Likely to widen inequalities | 2 |

| | | | | | | | |
|---|---|---|---|---|---|---|---|
| | | | | 5. Aerobic exercise regime | | | |
| **Haerens-2007 (Belgium)(11)** | | | | | | | |
| Design: Cluster-RCT<br>Aim: To investigate the short term impact of a computer tailored dietary fat intake intervention in adolescents | 7th grade pupils in 10 schools from two cities in Belgium (n=304) | Diet | Theory based computer tailored dietary fat intake intervention | Recipients: School children<br>Components (n=1)<br>1. Tailored CD-ROM containing computer tailored dietary fat intervention | Dietary fat intake<br><br>No difference | Education level<br><br>No preferential impact | 4 |
| **Havas-1998 (USA)(12)** | | | | | | | |
| Design: RCT<br>Aim: To increase fruit and vegetable consumption among women served by WIC programmes | Women attending Supplemental Nutrition Program for Women, Infants and Children, 16 sites in Baltimore City and 6 in Maryland (n=3122) | Diet | The intervention consisted of nutrition sessions conducted by peer educators; printed materials and visual reminders and direct mail. All designed to help participants progress through the stages of change. | Recipients: Low income mothers<br>Components (n=12)<br>*Nutrition education*<br>1. Brief messages at time of enrolment<br>2. Group discussion session<br>*Direct mail*<br>3. Tip sheets<br>4. Clue cards<br>5. Tailored letters<br>*Printed materials*<br>6. Photonovella<br>7. Booklet of recipes<br>8. Children's activities<br>9. Videotape for children<br>10. Refrigerator magnet<br>11. Calendar sheets and stickers<br>12. Fruit and vegetable posters | Fruit and vegetable consumption<br><br>Results favour intervention | Education level<br><br>Likely to widen inequalities | 5 |
| **Havas-2003 (USA)(13)** | | | | | | | |
| Design: RCT<br>Aim: To decrease percent of calories derived from fat and to increase fruit, vegetable and fibre intake among low-income women served by the WIC programme in Maryland | Women attending Supplemental Nutrition Program for Women, Infants and Children, 10 sites in Baltimore City and Maryland (n=2066) | Diet | Intervention participants were invited to five interactive nutrition sessions and sent written materials | Recipients: Low income mothers<br>Components (n=18)<br>1. 5 minute video with enthusiastic previous participants<br>2. Food for Life brochure<br>3. Individualised feedback on baseline data<br>4. Kick off fair<br>5. 4x45 minute workshops | % calories from fat, fruit and vegetable servings, fibre intake<br><br>Results favour intervention | Education level<br><br>Likely to widen inequalities | 5 |

| | | | | | | | |
|---|---|---|---|---|---|---|---|
| | | | | 6. Newsletters<br>7. Mail packets<br>8. Personalised invitations<br>9. Behaviour re-enforcing incentives<br>10. Phone calls<br>11. Group discussion on favourite foods and methods to cook<br>12. Product comparisons<br>13. Food demonstration at workshop<br>14. Healthy Meal<br>15. Tip sheets<br>16. Brief individualised counselling<br>17. Interactive cooking demonstration at fair<br>18. Free bag of food | | | |
| **Hobin-2014 (Canada)(14)** | | | | | | | |
| Design: Longitudinal study<br>Aim: To examine longitudinal changes in and factors associated with the PA trajectories of adolescents in Manitoba in secondary school | 31 schools in Manitoba Province, Canada (n=447) | PA | Province wide PE policy extending secondary school graduation requirements. | Recipients: School children<br>Components (n=3)<br>1. ”Core component” – In class study of health<br>2. “Flexible component” – exploring areas of interest/specialisation<br>3. “PA practicum” – PA participation in and out of class | Minutes MVPA per day<br><br>Results favour control | School neighbourhood disadvantage<br><br>Likely to reduce inequalities | Moderate |
| **Jeffery-1998 (USA)(15)** | | | | | | | |
| Design: RCT<br>Aim: To examine whether weight gain with age can be prevented using a low intensity intervention | 20-45 year old women in 4 local health departments in Minneapolis/St Paul, Minnesota metropolitan area (n=593) | Diet and PA | Monthly newsletter and face to face health education plus lottery to encourage participation | Recipients: Adults aged 20-45<br>Components (n=4)<br>1.Monthly newsletter<br>2. Postcard to encourage reflection on healthy habits<br>3. Face to face education programme<br>4. Lottery to incentivise participation | % energy from fat; Fruit servings p/day; Vegetable servings p/day<br>No difference | Income<br><br>Diet: No preferential impact | 3 |

| | | | | | | | |
|---|---|---|---|---|---|---|---|
| | | | | | PA score; walks p/week<br>PA: No difference | PA: No preferential impact | |
| **Lowe-2004 (UK)(16)** | | | | | | | |
| Design: Before-after study<br>Aim: To assess how a peer modelling and rewards based intervention changes children's consumption of fruit and vegetables | 4 – 11 year old children from 1 school in Wales and 2 in England (n=402) | Diet | Video adventures that include the Food Dudes and small rewards for consuming fruit and vegetables | Recipients: School children<br>Components (n=5)<br>1. Peer modelling videos<br>2. Rewards for healthy eating<br>3. Letters from cartoon characters to children<br>4. Homepacks to encourage healthy eating at home<br>5. Home sticker cards | Snack time fruit consumption; snack time vegetable consumption<br><br>Results favour intervention | % Free School Meals<br><br>Likely to widen inequalities | 3 |
| **Millett-2011 (UK)(17)** | | | | | | | |
| Design: Repeat cross-sectional<br>Aim: To evaluate the impact of the national salt strategy in England | Representative sample of adults living in England (n=1668) | Diet | National salt reduction strategy | Recipients: Adults in UK<br>Components (n=3)<br>Voluntary agreements with food industry to…<br>1. reduce salt content in processed foods<br>2. improve food labelling<br>3. implement public awareness campaigns to change individual behaviour | Daily salt intake<br><br>Results favour intervention | Social class<br><br>No preferential impact | 3 |
| **Moore-2014 (UK)(18)** | | | | | | | |
| Design: Cluster RCT<br>Aim: To examine universal provision of breakfast on socioeconomic inequalities in children's dietary behaviour | Children aged 9-11 from 111 schools in Wales (Baseline n=4350; Follow up n=4472) | Diet | Primary school free breakfast initiative | Recipients: School children<br>Components (n=1)<br>1. Provision of breakfast at school | Healthy items at breakfast; Unhealthy items at home<br><br>No difference | Free School Meal<br><br>No preferential impact | Moderate |
| **Reynolds-2000 (USA)(19)** | | | | | | | |
| Design: Cluster RCT<br>Aim: To evaluate the effects of a school based dietary | Fourth grade children from 28 elementary | Diet | Three component intervention consisting of classroom component, | Recipients: School children<br>Components (n=9)<br>*Classroom component* | Fruit consumption; | Household income | 3 |

| | | | | | | | |
|---|---|---|---|---|---|---|---|
| intervention to increase fruit and vegetable intake | schools (n=1698 families) | | parent component and food service component | 1. Lessons in school<br>*Parent component*<br>2. Homework assignments<br>3. Brochures<br>4. Information evening<br>5. Recipes<br>6. Items to trigger behaviour<br>*Food service component*<br>7. Half day training<br>8. Cafeteria tasks and ratings<br>9. Visits from nutritionists | Vegetable consumption<br><br>Results favour intervention | No preferential impact | |
| **Robroek-2012 (Netherlands)(20)** | | | | | | | |
| Design: Cluster-RCT<br>Aim: To evaluate cost-effectiveness of a long term workplace health promotion programme on PA and diet | 76 departments within 6 workplaces (healthcare organisations, commercial services and executive branch of government) (n=924) | Diet and PA | Additional website functionalities includes action-oriented feedback, self-monitoring, possibility to ask questions and monthly email messages | Recipients: Workplace volunteers<br>Components (n=5)<br>1.Computer tailored advice on self-report PA and diet<br>2. Online self-monitors on fruit and veg intake, PA and weight to monitor progress<br>3. Food frequency questionnaire assessing saturated fat intake<br>4. Possibility to submit questions to health professionals<br>5. Monthly email messages | Fruit intake; vegetable intake<br><br>Diet: No difference<br><br>PA: No difference | Education level<br><br>Diet: No preferential impact<br><br>PA: No preferential impact | Moderate to strong |
| **Rosenkranz-2010 (USA)(21)** | | | | | | | |
| Design: Cluster RCT<br>Aim: To evaluate an intervention designed to prevent obesity by modifying Girl Scout troop meeting environments | 7 Girl Scout Troops consisting of girls in 4th and 5th grades (n=76) | Diet and PA | Environmental intervention in girl scout troops based on three components of social cognitive theory: Role modelling; skill building; enhancement of self-efficacy and proxy efficacy; and | Recipients: Girl Scouts<br>Components (n=11)<br>*Interactive educational curriculum*<br>1. Discussion of target behaviours<br>2. Worksheet for goal setting and monitoring<br>3. Physically active recreation session<br>4. Fruit and Veg snack recipe preparation<br>5. Family meal role-playing | FV daily consumption; SSB daily consumption<br><br>Diet: Results favour intervention | Socioeconomic status<br><br>PA: No preferential impact | Strong |

| | | | | | | | |
|---|---|---|---|---|---|---|---|
| | | | reinforcement of behaviour | *Troop meeting policies implemented by troop leaders*<br>6. Providing 15 minutes per meeting for physically active recreation<br>7. Troop leaders participation in PA with girls<br>8. Provision of a fruit and veg snack prepared by girls<br>9. Troop leaders eating fruit and veg snack with girls<br>10. Troop leaders verbally promoting PA, Fruit and veg in meetings and home<br>11. Prohibition of SSB, candy and TV watching during meetings | PA: Results favour intervention | | |
| **Simon-2008 (France)(22)** | | | | | | | |
| Design: Cluster RCT<br>Aim: To assess a PA intervention in adolescents | 8 middle schools in Eastern France (n=954) | PA | The intervention was designed to promote PA by changing attitudes through debates and attractive activities and by providing social support and environmental changes encouraging PA | Recipients: School children<br>Components (n=7)<br>1.Educational component focusing on PA and sedentary behaviour<br>2. PA opportunities at lunchtime<br>3. PA opportunities during breaks<br>4. PA opportunities after school<br>5. Sporting events<br>6. Cycle to school days<br>7. Regular meetings with parents and educators | Supervised leisure PA; Active home-school commuting<br><br>No difference | Occupation category<br><br>No preferential impact | Strong |
| **Smith-1997 (Australia)(23)** | | | | | | | |
| Design: RCT<br>Aim: To estimate population uptake and effectiveness of dietary behaviour change counselling in higher and lower socio-economic status groups | Randomly selected population sample in Adelaide (n=479) | Diet | The intervention was based on behaviour change techniques and social learning theories and included a one time counselling session with follow up after 3 months | Recipients: Adults<br>Components (n=3)<br>1.Dietary counselling session<br>2. Booklet containing cholesterol test result<br>3. Self-monitoring charts | Food group and nutrient consumption<br><br>No difference | Occupational prestige<br><br>No preferential impact | 4 |

| | | | | | | | |
|---|---|---|---|---|---|---|---|
| | | | including individualised feedback and advice | | | | |
| **Stables-2002 (USA)(24)** | | | | | | | |
| Design: Repeat cross sectional<br>Aim: To assess population-based changes in vegetable and fruit consumption and psychosocial correlates | Representative sample of US adults (Baseline – n=2755; Follow up – n=2544) | Diet | The 5 a day for better health program is a nutrition education campaign designed to increase awareness of the need to consume more FV | Recipients: Adults<br>Components (n=1)<br>1. 5-a-day for better health program (nutrition education) | No difference | Poverty Index Ratio<br><br>No preferential impact | 2 |
| **Sturm-2010 (USA)(25)** | | | | | | | |
| Design: Cross-sectional with unexposed group<br>Aim: To examine whether small taxes are likley to change consumption and weight gain or whether larger taxes would be needed | Nationally representative US kindergartners (n=7403) | Diet | State level soda taxes | Recipients: School children<br>Components (n=1)<br>1. Taxation of soft drinks | SSB consumption<br><br>No difference | Family income<br><br>Likely to reduce inequalities | Moderate |
| **Taber-2013 (USA)(26)** | | | | | | | |
| Design: Cross-sectional with unexposed group<br>Aim: To determine if state laws with stricter school meal nutrition standards are inversely associated with adolescent weight status | 8th grade students in 40 US states (n=4870) | Diet | State laws governing school meal nutrition standards | Recipients: School children<br>Components (n=1)<br>1. State laws governing school meal nutrition standards | Fast food and confectionary intake<br><br>No difference | Free school meal provision<br><br>No preferential impact | Moderate |
| **Tak-2008 & 2010 (Netherlands) (27, 28)** | | | | | | | |
| Design: Longitudinal study<br>Aim: To evaluate long term effects of a Dutch primary school based intervention providing free fruit and vegetables | 9-10 year old children in 44 Dutch schools in 5 cities (n=771) | Diet | Free fruit and veg in schools improving the availability, accessibility and exposure to fruit and veg in schools | Recipients: School children<br>Components (n=4)<br>1. Provision of a portion of free fruit or veg twice a week<br>2. Optional school curriculum to increase knowledge and skills<br>3. Provision of a lunchbox | Results favour intervention | Parental education<br><br>No preferential impact | Strong |

| | | | | | | | |
|---|---|---|---|---|---|---|---|
| | | | | 4. Provision of a calendar for teachers indicating when FV would be provided | | | |
| **Toftager-2014 (Denmark)(29)** | | | | | | | |
| Design: Cluster RCT<br>Aim: To assess the effectiveness of a multi-component environmental school-based intervention to reduce the age-related decline in PA among adolescents | 14 schools in Southern Denmark (n=1348) | PA | Intervention changing the physical and organisational environment of the schools | Recipients: School children<br>Components (n=14)<br>1. Upgrading existing outdoor areas at the school for PA<br>2. Building playgrounds designed for adolescents<br>3. Improving safety for active transport to and from school<br>4. Establishing an afterschool fitness program<br>5. Implementing school PA policy<br>6. Educating teachers as 'kick starters' who facilitate PA during breaks<br>7. School play patrol<br>8. Mandatory outdoor recess and/or free access to gym/sports hall<br>9. School traffic patrol<br>10. Safe cycling training for students<br>11. School project/theme week once a year | FV bought from home; Unhealthy food bought from home<br><br>No difference | Household income<br><br>No preferential impact | Moderate |
| [a] Quality assessment as assigned in the parent systematic reviews<br>MVPA = moderate to vigorous physical activity; FV = Fruit and vegetable; SSB = Sugar Sweetened Beverage | | | | | | | |
